# COVID-19 vaccination rate and protection attitudes can determine the best prioritisation strategy to reduce fatalities

**DOI:** 10.1101/2020.10.12.20211094

**Authors:** Jorge Rodríguez, Mauricio Patón, Juan M Acuña

## Abstract

**Background:** The unprecedented rapid development of vaccines against the SARS-CoV-2 virus creates in itself a new challenge for governments and health authorities: the effective vaccination of large numbers of people in a short time and, possibly, with shortage of vaccine doses. To whom vaccinate first and in what sequence, if any at all, to avoid the most fatalities remains an open question.

**Methods:** A compartmental model considering age-related groups was developed to evaluate and compare vaccine distribution strategies in terms of the total avoidable fatalities. Population groups are established based on relevant differences in mortality (due to e.g. their age) and risk-related traits (such as their behaviour and number of daily person-to-person interactions). Vaccination distribution strategies were evaluated for different vaccine effectiveness levels, population coverage and vaccination rate using data mainly from Spain.

**Findings:** Our results show that, if children could also be included in the vaccination, a rollout by priority to groups with the highest number of daily person-to-person interactions can achieve large reductions in total fatalities. This is due to the importance of the avoided subsequent infections inflicted on the rest of the population by highly interactive individuals. If children are excluded from the vaccination, the differences between priority strategies become smaller and appear highly depending on rollout rate, coverage and the levels of self-protection and awareness exercised by the population.

**Interpretation:** These results are in possible contradiction with several published plans for COVID-19 vaccination and highlight the importance of conducting an open comprehensive and thorough analysis of this problem leaving behind possible preconceptions.

## Introduction

COVID-19 has inflicted great stress worldwide with more than 95 million cases and over two million deaths globally by January, 2021. (1,2). The pandemic is not near its end and the disease will probably not go away, becoming endemic in many regions throughout the world. In this scenario, the development of adequate vaccines is an alternative for primary prevention of the disease, so far implemented through hygiene, massive testing and isolation, social distancing, and lockdowns.

Vaccines, of different types, are products of biotechnological processes which serve two functions: to protect the individual from contracting the disease and to stop the transmission of the disease (3). The development and approval of vaccines has been historically long, taking on average several years (4). This was the rule until now: an unprecedented global effort from pharma and academia, supported by government and private organizations, has made possible that one or more COVID-19 vaccines becoming available by 2021 (5).

With several vaccines already authorised and hundreds of candidates and trials now underway (35 in phase I; 26 in phase II; and 20 in phase III), the next problems to sort out will probably be mass production of the vaccine and how to allocate the available vaccines to potential users (6,7).

The WHO issued a draft statement on September 9^th^, 2020, addressing issues related to the fair allocation of vaccines for countries around the world. Available doses of COVID-19 vaccines worldwide are proposed to be managed centrally and equitably by the COVAX Access Mechanism, a coalition of member countries together with the ACT-Accelerator Collaboration (WHO; the Bill & Melinda Gates Foundation; Gavi, the Vaccine Alliance; the Global Fund; the Coalition for Epidemic Preparedness Innovations -CEPI; and Welcome Trust) (8) to allow fair allocation of available vaccines. The COVAX initiative (8) proposes vaccine doses for each country to cover 3% to 20% of the population proportionally to each country. There are however some small, high income countries that may be capable of securing vaccine doses for their entire population at early stages too.

Despite the estimated and unprecedented fast production of vaccines, the number of vaccines initially available will be insufficient, due to the fact that the vast majority of the population worldwide is still susceptible to the infection. There is no doubt that vaccine uptake will be an important driver (9) however, intra-country capacity for adequate vaccine distribution will be essential and will most likely impact the final amount of deaths related to COVID-19 (10). Some of the important drivers for vaccine allocation seem to rise from ethical principles (11,12). For instance, these ethical principles may point towards first, reducing premature deaths; then, reducing economic and social serious deprivations and last, focus on return to functional populations and societies (13). However, as disputed the order and scope of these principles may be, the immediate need of decreasing COVID-19 deaths (direct and indirect) that may be avoidable, should be the first priority.

Decisions based on empirical observations are a luxury not always possible during a pandemic and mathematical modelling provides a tool to guide immediate decisions. Although several epidemic models for the COVID-19 outbreak have been developed to forecast the extent of the pandemic or the impact of interventions for different regions (14–21), any predictive mathematical model has limitations, which are inherent to the model itself but also to the quality of the input data. Models can lead to great predictive failures (22) however they can also serve as valuable tools to better understand non-trivial underlying interactions in complex processes and to comparatively evaluate public health strategies.

Best vaccine distribution strategies help to decrease the number of fatalities by protecting the groups at risk directly (via vaccination of the individuals that belong to those groups of risk) and indirectly (via vaccination of individuals that are most in contact with those at risk) (3). Vaccination itself changes the characteristics of the population in real-time, changing the probability of death of different groups as time elapses throughout the vaccination campaigns. Considering this, a fixed approach (e.g. vaccinating a particular population group) during a prolonged amount of time as opposed to a dynamic changing adaptive approach, might not produce the best possible results in averting the highest potential number of preventable deaths by vaccination. Dynamic models can be used to inform which group should be vaccinated and at which moment of the vaccination campaign while accounting for the changing characteristics of the population due to the vaccination itself.

If we consider that vaccination has the indirect benefit of decreasing transmission, thereby reducing the infection risk also to those who have not been vaccinated, optimum strategies for vaccine allocation can become counterintuitive and complex. Existing past studies for influenza have already shown how targeting the vaccination of lower-risk and high-transmission groups first may achieve superior results (23). Recent studies have focused on the distribution strategies of vaccines for SARS-CoV-2 (24–31). All these works concluded that prioritisation of certain groups for the distribution of vaccines could substantially reduce fatalities. Some of these works recommended to prioritise vaccination of elders (24,28,32,33) while other works recommended alternative prioritisation strategies in certain conditions (25,30).

In this work, a new COVID-19 SEIRD-type model, segregated by population age groups, based on that by (19), is applied to evaluate and compare vaccine distribution strategies in terms of what group prioritisation should be followed. Although slightly different optimisation goals are possible, the minimum total final fatalities after the vaccination campaign and outbreak is the goal pursued. Several example scenarios using demographic and epidemiological data from Spain are evaluated.

## Methodology

### Dynamic COVID-19 model with vaccination

Based on our previous work (19) a mathematical model was developed specifically for the optimum vaccine allocation problem at hand. A complete description of the model is provided in the Supplementary Information, Section I.

The model allocates individuals of the population in compartments segregated by age groups and by infection or disease severity stage (Figure 1). The population groups are defined based on meaningful differences both epidemiological (in severity and mortality) and of behaviour or activity traits that affect their risk of infection. Individuals in the population belong in only one group which they never abandon.

**Figure 1.**
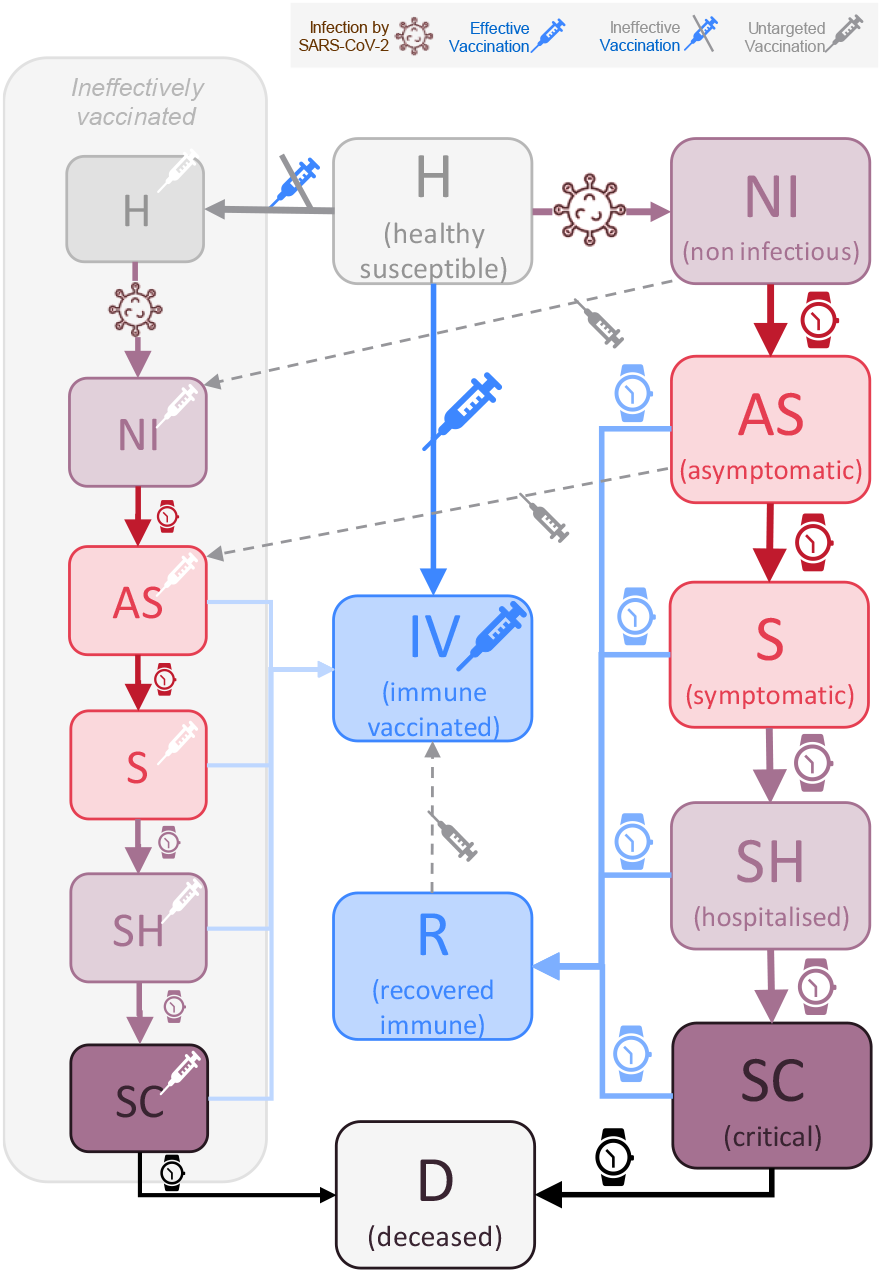
Schematic representation of the model compartments in terms of infection and disease severity stages. Infection can only occur by contact of healthy susceptible individuals with infectious either asymptomatic or symptomatic individuals (red boxes). Individuals spend in each stage an average amount of time depending on their transition path towards recovery or increased severity. Vaccination, if effective, avoids infection and transmission and places individuals at immune vaccinated stage. Ineffective vaccination maintains individuals in their disease stage although accounted separately as already vaccinated.

In addition, the population groups are segregated by age groups (see Supplementary Information Table S3.1). This segregation can be related to their age but also to any other arbitrary classification based on these epidemiological and behavioural differences. For example, groups such as front-line workers, individuals with co-morbidities with a very high risk of mortality could be grouped separately. The most suitable groups definition may differ and be country or region specific however group-specific reliable data will always be required and that poses limitations to the number and types of groups.

Although the model individuals never leave the (age-related) population group to which they belong and, at a given time, they sit on and may transition through the different infection and disease severity stages. These stages are defined in terms of infectiousness and severity of symptoms analogously as in (19), namely: *healthy susceptible* (H); *infected non-symptomatic non-infectious* (NI); *asymptomatic infectious* (AS); *symptomatic infectious* (S); *in need of hospitalisation* (SH); *in need of critical care* (SC); *recovered immune* (R) and *deceased* (D), in addition, those effectively vaccinated become *vaccinated immune* (IV). Individuals ineffectively vaccinated (i.e., the vaccine does not immunise them against infection, severity or transmission) maintain their current stage and they are simply accounted separately as already vaccinated (see Figure 1).

Figure 1 provides an overview of these different stages and their transitions. The disease severity progression and vaccination govern the transitions of individuals between disease stages. Vaccination causes individuals to transition either to vaccinated immune or to ineffectively vaccinated non-immune stages. Most parameters involved on the disease transitions (i.e., times from asymptomatic to develop symptoms) are either already available from recent clinical and/or epidemiological information (see Supplementary Information section III).

To calculate the rates of infection (Figure 2), a number of daily contacts (ni) that individuals in each age group have with the other age groups is required. To define these, a contact matrix for Italy was used (30). We considered this source as being the most relevant and similar to the contact pattern expected for Spain. Other contact matrices (36) can and have been used in other models (37). The likelihood of infection per contact is calculated based on the level of protection and awareness for each age group. These type of parameters (e.g., level of protection awareness or number of interactions) are also mechanistic and can be interpreted. These behaviour related parameters can be surveyed for among the target populations or communities. A complete reproducible description of the equations, variables and parameters of the model is provided in the Supplementary Information sections I-V.

**Figure 2.**
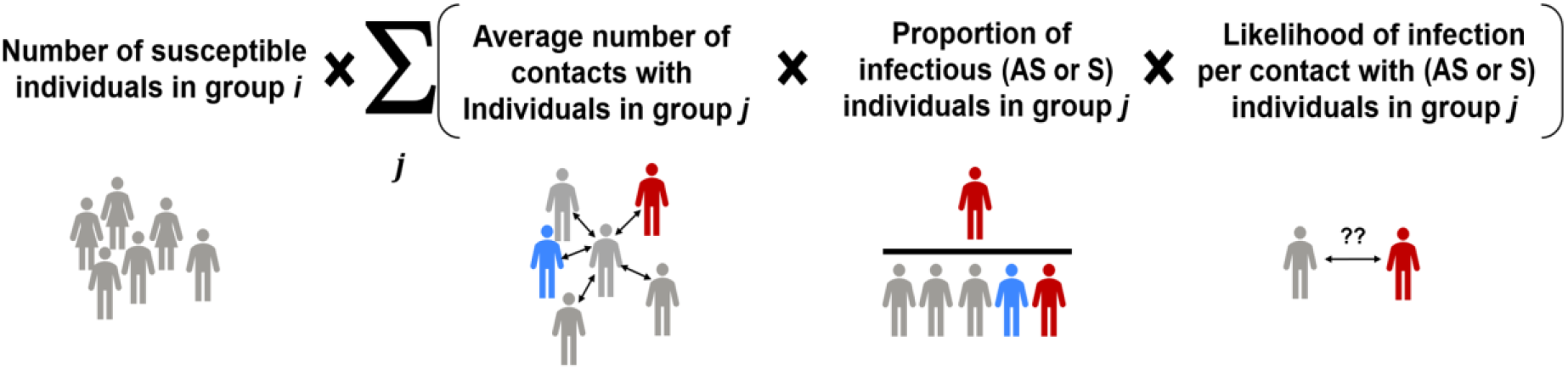
The rate of infection of heathy susceptible individuals from a group *i* (see also Eq. S1.4) is the sum of the infections by asymptomatic and symptomatic individuals from all other *j* groups.

### Model limitations and assumptions

Although the model was specifically developed for the optimum vaccination problem at hand, it shares many of the fundamental characteristics of compartment SEIRD-type models as well as their limitations. As for the case of our earlier model (19), this type of models consider all individuals located in a common single domain or closed community. Since no geographical clustering or separation, neither any form of migration in or out of the community are captured, large cities with ample use of public transportation remain the best described by this and other SEIRD-type disease propagation models.

In these models, variables and parameters refer to representative averages for each compartment stage and population group. These characteristics may limit the model representation of non-linear relevant phenomena that may occur in the real-world that could be better described by agent-based models. Examples of these include phenomena such as the so-called super spread events or other location specific phenomena that cannot be described by SEIRD-type models. Any quantitative application of this model and any other model for prediction purposes in public health must always be accompanied by a critical discussion against these limitations (38).

In addition to the above, other specific assumptions in this model, that are of special relevance to the vaccination problem at hand, include:

- All ever-infected individuals that recover become fully immune, irrespective of their severity path, and cannot be infected or infectious again. Reinfections are not modelled in this work.
- The vaccine effectiveness was assumed to be protective against both infection and transmission. This can be revised in future model versions as specific information on effectiveness against transmission becomes available for the vaccines.
- No differences in immunity (or any other epidemiological aspect) were considered between individuals ineffectively vaccinated and those never vaccinated.
- Eligible individuals to be vaccinated are those who belong to one of the following groups: Healthy (H), non-infectious (NI), asymptomatic (AS) and recovered (R).
- If multiple groups are called for vaccination simultaneously, their rates of vaccination are proportional to the groups relative sizes in terms of their eligible unvaccinated individuals. The total vaccination rate will match the logistic rate limit set by the capacity of the health system or by the availability of vaccine doses.

### Evaluation of vaccine distribution strategies

Although the model can be applied to evaluating complex scenarios simultaneously combining non pharmaceutical interventions (NPIs) (such as the isolation of specific groups) and vaccine distribution strategies, only the later are evaluated here.

A plethora of optimisation methods can be attempted to find the best possible vaccination strategies on a given model and are available in the literature. This work focused on simple strategies such that they can be easily applied by a policy maker when distributing the vaccines to the population. Five different strategies for vaccine distribution in terms of population groups were compared, namely:

i. *No group prioritisation*: all population groups are called for vaccination in equal terms.
ii. *Priority by mortality*: Age groups are prioritised for vaccination according to their mortality per infection (from highest to lowest). Groups are called one by one and only once (single call) until coverage for that group is reached.
iii. *Priority by interactivity*: Population age groups are prioritised for vaccination according to their number of interactions (daily contacts) (from highest to lowest). Groups are called one by one and only once (single call) until coverage is reached.
iv. *R-based Projected Avoidable Deaths (RbPAD) criteria*. A computationally inexpensive, model-independent optimisation method was specifically developed for the vaccine distribution problem as part of this work. The method (fully described in the Supplementary Information section VI) is based on the estimation of the projected avoidable deaths (PAD) per vaccination in each group at each moment in time. To that end the dynamic R_t_ number and the group’s own mortality are used. The method allocates priority to the group *i* with the highest PAD number per vaccination. The PAD are calculated as function of three elements, namely (i) the risk of infection in that group (as the ratio of the present rate of infections and the number of individuals in the group); (ii) the mortality per infection in the group (f_d_ni_^i^) (from epidemiological data) and (iii) the mortalities of the projected secondary and subsequent infections avoided that a vaccinated group member would have inflicted on the entire population (obtained via estimated R matrix). This strategy is evaluated for weekly cycles and allows for multiple partial calls to any specific group.
v. *The best of all the possible sequences of groups priority*. All possible permutations of the age groups as defined were evaluated assuming they are called one by one and only once. This approach selects the permutation with lowest number of deaths as the best sequence. This approach required intensive computation as all single call strategies must be evaluated (9! = 362,880 simulations). This approach is very sensitive to model accuracy and of difficult practical implementation however it is presented to provide an estimation of how close the different strategies are from optimality.

## Results and discussion

The model was applied for the population demographic distribution of Spain also applicable to other European and developed countries. Population groups were defined by (age-related) activity as namely: *preschool children* (ages 0-4); *school children* (ages 5-14); *higher school and university young* (ages 15-24), *young workers* (ages 25-49); *mature workers* (ages 50-59); *senior workers* (ages 60-64); *early retired* (ages 65-69); *retired* (ages 70-79); *elderly* (ages 80+). This definition of groups is partly arbitrary, but it is based on the criteria of meaningful differences in mortality and behaviour that impact the problem at hand.

The example scenario was selected with the following initial conditions and assumptions:

- A total population of 47,026,208 as for Spain in 2019 was considered.
- No initially vaccinated individuals nor fatalities.
- Initial number of recovered individuals (R) considered to be of approximately 10% of the population, based on the results of the fourth seroprevalence study of Spain in December (39).
- Initial case incidence was set as of 200 active cases per 100,000 population. These number of cases were distributed proportionally based on their transition times between non-infectious (NI), asymptomatic (PS) and symptomatic (S).
- All other individuals initially considered as healthy susceptible (H).
- Different vaccine effectiveness and maximum population coverage values were evaluated. Vaccines effectiveness is defined as the proportion of vaccinated individuals who become **immune and non-transmitters**. Population coverage is defined as the maximum percentage of the population that can be reached and accepts the vaccine.
- Different daily constant vaccination rates ranging from 0.25-1.5% of the population per day until the campaign ends were evaluated. The rates covered are based on initial values observed in January 2021 (40).

For the example scenario, two types of vaccine distribution scenarios are presented, namely (i) distribution of the vaccine to the whole population and (ii) distribution of the vaccine to the whole population except children. The parameters shown in Table S3.1 and Tables S5.1-2 were used.

### Scenario 1: Vaccine can be distributed to all age groups

The first scenario evaluates for a vaccine that is available to all population groups (including children) and under sufficient doses available at different rates of vaccination rollout. A summary of the results obtained for the five vaccine distribution strategies described above is presented in Table 1. In addition, the scenario of no vaccination applied is presented for comparison purposes. A summary of results for different values of vaccine effectiveness, daily vaccination rate and maximum population coverage are presented in Table 1. The full set of combinations evaluated are shown in the Supplementary Information (Section XI).

**Table 1.** Performance of the different vaccine distribution strategies (children included) for different vaccine effectiveness, daily vaccination rate and population coverage.

| Input parameters |  |  | Total Fatalities per vaccination priority sequence after 360 days |  |  |  |  |  |
| --- | --- | --- | --- | --- | --- | --- | --- | --- |
| Vaccine Effectiveness | Daily Vaccination Rate | Population Vaccination Coverage | No Prioritisation by Groups | by Mortality (highest-to-lowest) | by Interactions (highest-to-lowest) | RbPAD method | Best of all possible (single call) | No Vaccination |
| 75% | 0.25% | 25% | 38,288 | 41,011 | 33,836 | 34,819 | 33,290 | 70,020 |
| 75% | 0.25% | 50% | 36,755 | 38,932 | 27,882 | 33,000 | 27,147 | 70,020 |
| 75% | 0.25% | 80% | 36,659 | 39,558 | 30,359 | 32,807 | 29,127 | 70,020 |
| 75% | 0.75% | 25% | 29,994 | 30,574 | 28,688 | 28,652 | 28,513 | 70,020 |
| 75% | 0.75% | 50% | 11,407 | 17,137 | 7,802 | 9,085 | 7,760 | 70,020 |
| 75% | 0.75% | 80% | 9,022 | 15,013 | 5,377 | 6,589 | 5,213 | 70,020 |
| 75% | 1.25% | 25% | 29,489 | 29,687 | 28,409 | 28,249 | 28,249 | 70,020 |
| 75% | 1.25% | 50% | 7,400 | 9,540 | 5,927 | 6,715 | 5,908 | 70,020 |
| 75% | 1.25% | 80% | 4,129 | 7,319 | 3,263 | 3,703 | 3,108 | 70,020 |
| 88% | 0.25% | 25% | 34,425 | 37,890 | 29,082 | 30,324 | 28,623 | 70,020 |
| 88% | 0.25% | 50% | 32,827 | 35,503 | 22,699 | 28,122 | 22,319 | 70,020 |
| 88% | 0.25% | 80% | 32,727 | 35,400 | 26,476 | 26,440 | 25,371 | 70,020 |
| 88% | 0.75% | 25% | 24,939 | 25,672 | 23,642 | 23,647 | 23,506 | 70,020 |
| 88% | 0.75% | 50% | 8,043 | 13,621 | 5,128 | 6,215 | 5,090 | 70,020 |
| 88% | 0.75% | 80% | 6,814 | 11,527 | 4,628 | 5,563 | 4,400 | 70,020 |
| 88% | 1.25% | 25% | 24,322 | 24,553 | 23,313 | 23,247 | 23,191 | 70,020 |
| 88% | 1.25% | 50% | 4,662 | 6,610 | 3,591 | 4,253 | 3,564 | 70,020 |
| 88% | 1.25% | 80% | 3,369 | 5,430 | 2,961 | 3,243 | 2,751 | 70,020 |

The detailed simulation results for the five strategies evaluated with a vaccine effectiveness of 87.5 %, a population coverage of 80% and a daily vaccination rate of 0.75% of the population are presented in Figure 3. The detailed results of all the cases in Table 1 are shown in the Supplementary Information Section VII.A.

**Figure 3.**
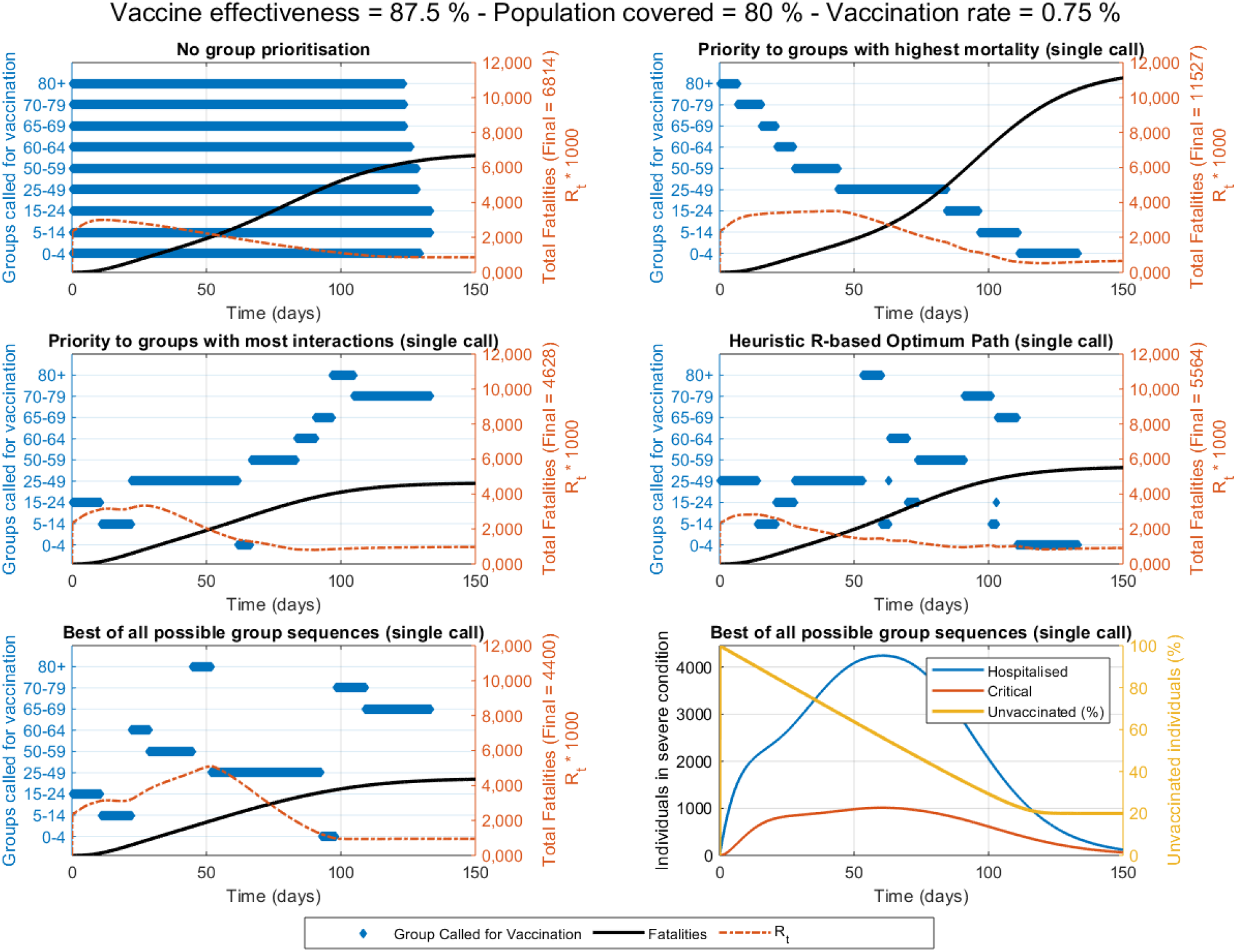
Details of the vaccination strategies with all population (including children) eligible for vaccination. Active groups called for vaccination, fatalities and R_t_ value over time are shown for each strategy. Case for a daily vaccination rollout rate of 0.75% of the total population, a vaccine effectiveness of 87.5% and a population coverage of 80%. The best result was obtained after (computationally intensive) model evaluation of all possible group sequences. The computationally inexpensive RbPAD strategy is evaluated for weekly cycles and allows for partial calls to specific groups.

The different strategies evaluated lead to significantly different outcomes in terms of fatalities respect to the base case of no group prioritisation. Under the scenario with all population (including children) eligible to be vaccinated, prioritisation by mortality results in a number of deaths higher than if no prioritisation is followed. In some cases, the relative difference between strategies becomes substantial, particularly in the cases in which coverage is over 50% (e.g. ∼70% worse when vaccination rate is 0.75% and coverage is 50 or 80%). On the contrary, a prioritisation by interactions (from highest to lowest), results in substantial reductions in fatalities compared to no prioritisation (∼40% lower at vaccine effectiveness of 75%, vaccination rate of 0.75% and population coverage of 80%) (Table 1). Intensive computational evaluation of all possible permutations of the nine groups (for single group calls) provided an additional reduction in fatalities compared to the strategy that follows a prioritisation by interactions (∼3-5% % lower number of fatalities). The main conclusion from Table 1 is that, contrary to existing public health thought, a vaccine effective both against infection and transmission would have a greater impact by following a prioritisation based on interactions than following a prioritisation based on mortality, especially at a population coverage of 50% and above. This falls in line with previous reported findings (25), in which over 50% coverage of the youngest should be vaccinated first. These results differ with the conclusions in other publications (or pre-prints) that conclude that elders should be prioritised (24,28,32,33). The difference in outcomes is attributed to the self-protection levels allocated to the elders due to the characteristics of the disease (see *Impact of behavioural parameters* section).

Figure 3 shows the details of each vaccination strategy. The prioritisation by mortality is almost inverse to that by interactions, due to the higher levels of interactivity in younger groups while the RbPAD optimisation method and the best of all sequences provide mixed orders of the groups for vaccination. A side-by-side comparison between prioritisation strategies by mortality or by interactions is shown in Figure 4 both against a no prioritisation strategy.

**Figure 4.**
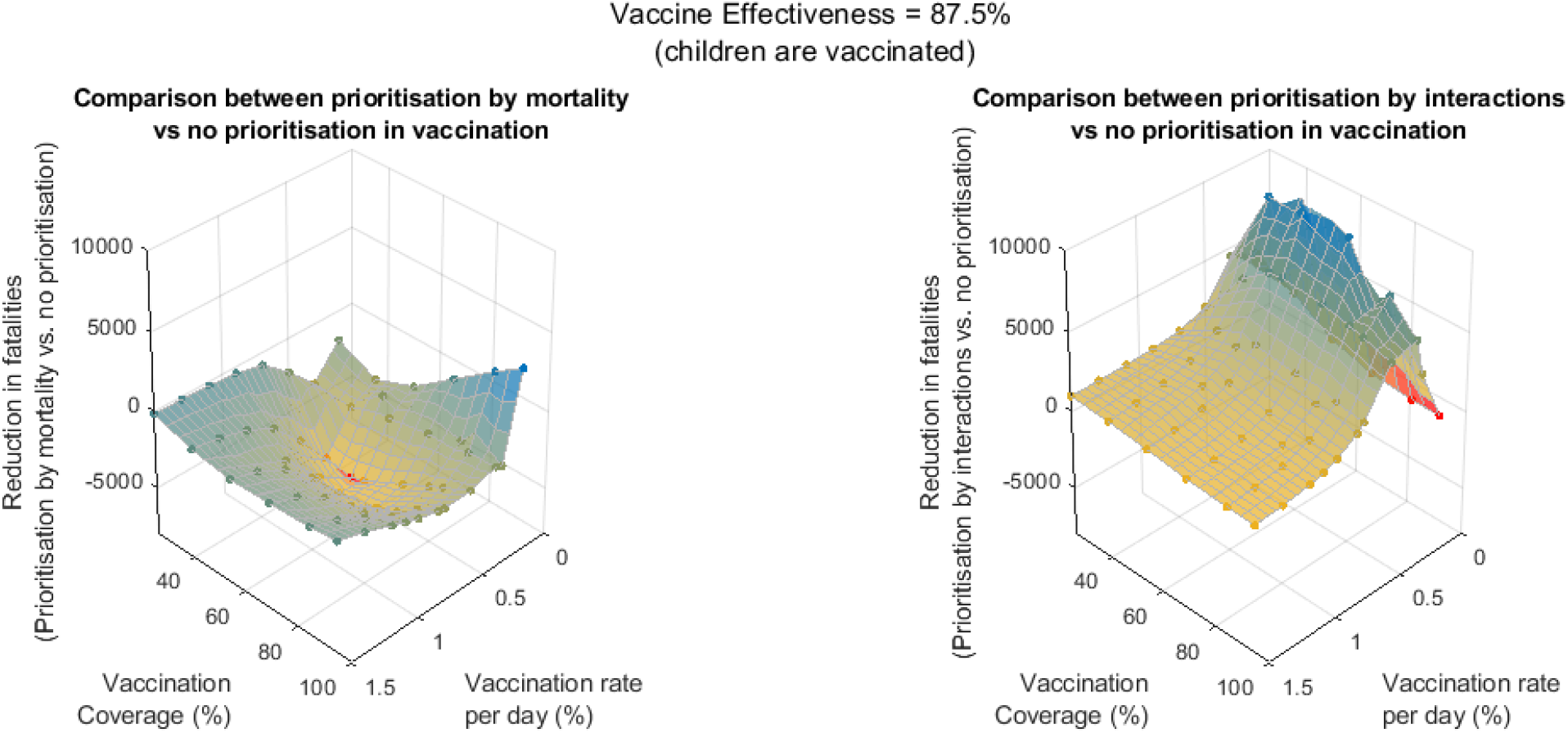
Reduction in fatalities if prioritisation by mortality (left) or by interactions (right) is used instead of a no prioritisation strategy, for different rates of vaccination rollout and coverage. Case for a vaccine effectiveness of 87.5% with children included in the vaccination and under the behavioural and epidemiological parameters from Tables S3.1, S5.1 and S5.2.

Figure 4 (left) for a vaccine effectiveness of 87.5% suggests that prioritising by mortality appears to be worse than no prioritisation in practically all cases except marginally the opposite in extremes of very low daily vaccination rollouts (0.1 %) and for very high coverage (95 %). On the other hand, Figure 4 (right) suggests prioritising by interactions appears to be practically equal to no prioritisation at high vaccination rates but substantially superior strategy (a reduction of over 8000 fatalities) at low daily vaccination rollouts. These results (Figure 4) include children in the vaccination campaign.

### Scenario 2: Vaccination is not distributed to children

Due to the absence of finalised trials, no vaccine for COVID-19 has been yet approved to be used in children. Therefore, a second scenario is presented that evaluates for a vaccine that is available to all population groups excluding children, also under sufficient doses available at different rates of vaccination rollout. A summary of the results obtained for the five vaccine distribution strategies described above is presented in Table 2. Results for the same values of vaccine effectiveness, daily vaccination rate and maximum population coverage are presented as well as the scenario of no vaccination for comparison purposes. The full set of combinations evaluated are shown in the Supplementary Information (Section XI).

**Table 2.** Performance of the different vaccine distribution strategies (children not vaccinated) for different vaccine effectiveness, daily vaccination rate and population coverage

| Input parameters |  |  | Total Fatalities per vaccination priority sequence after 360 days |  |  |  |  |  |
| --- | --- | --- | --- | --- | --- | --- | --- | --- |
| Vaccine Effectiveness | Daily Vaccination Rate | Population Vaccination Coverage | No Prioritisation by Groups | by Mortality (highest-to-lowest) | by Interactions (highest-to-lowest) | RbPAD method | Best of all possible (single call) | No Vaccination |
| 75% | 0.25% | 25% | 39,596 | 41,120 | 37,798 | 38,080 | 37,146 | 70,020 |
| 75% | 0.25% | 50% | 37,759 | 38,932 | 34,201 | 35,039 | 30,953 | 70,020 |
| 75% | 0.25% | 80% | 37,667 | 39,558 | 33,672 | 34,623 | 30,098 | 70,020 |
| 75% | 0.75% | 25% | 34,236 | 34,561 | 34,410 | 34,402 | 34,388 | 70,020 |
| 75% | 0.75% | 50% | 17,974 | 19,712 | 17,291 | 17,416 | 17,028 | 70,020 |
| 75% | 0.75% | 80% | 12,973 | 15,150 | 11,696 | 11,683 | 10,020 | 70,020 |
| 75% | 1.25% | 25% | 34,046 | 34,241 | 34,255 | 34,279 | 34,248 | 70,020 |
| 75% | 1.25% | 50% | 16,282 | 16,729 | 16,226 | 16,273 | 16,157 | 70,020 |
| 75% | 1.25% | 80% | 8,389 | 9,409 | 8,135 | 8,173 | 7,709 | 70,020 |
| 88% | 0.25% | 25% | 36,169 | 38,015 | 34,138 | 34,257 | 33,449 | 70,020 |
| 88% | 0.25% | 50% | 34,248 | 35,504 | 30,414 | 31,336 | 26,924 | 70,020 |
| 88% | 0.25% | 80% | 34,154 | 35,400 | 30,182 | 30,512 | 26,385 | 70,020 |
| 88% | 0.75% | 25% | 30,419 | 30,704 | 30,478 | 30,510 | 30,459 | 70,020 |
| 88% | 0.75% | 50% | 14,443 | 16,068 | 13,760 | 13,838 | 13,450 | 70,020 |
| 88% | 0.75% | 80% | 10,231 | 11,634 | 9,624 | 9,415 | 7,744 | 70,020 |
| 88% | 1.25% | 25% | 30,258 | 30,313 | 30,305 | 30,328 | 30,301 | 70,020 |
| 88% | 1.25% | 50% | 12,747 | 13,235 | 12,695 | 12,715 | 12,604 | 70,020 |
| 88% | 1.25% | 80% | 6,254 | 6,943 | 6,237 | 6,235 | 5,746 | 70,020 |

The detailed simulation results for the five strategies evaluated with a vaccine effectiveness of 87.5%, a population coverage of 80% and a daily vaccination rate of 0.75% are presented in Figure 5. The complete results are shown in the Supplementary Information Section VII-B.

**Figure 5.**
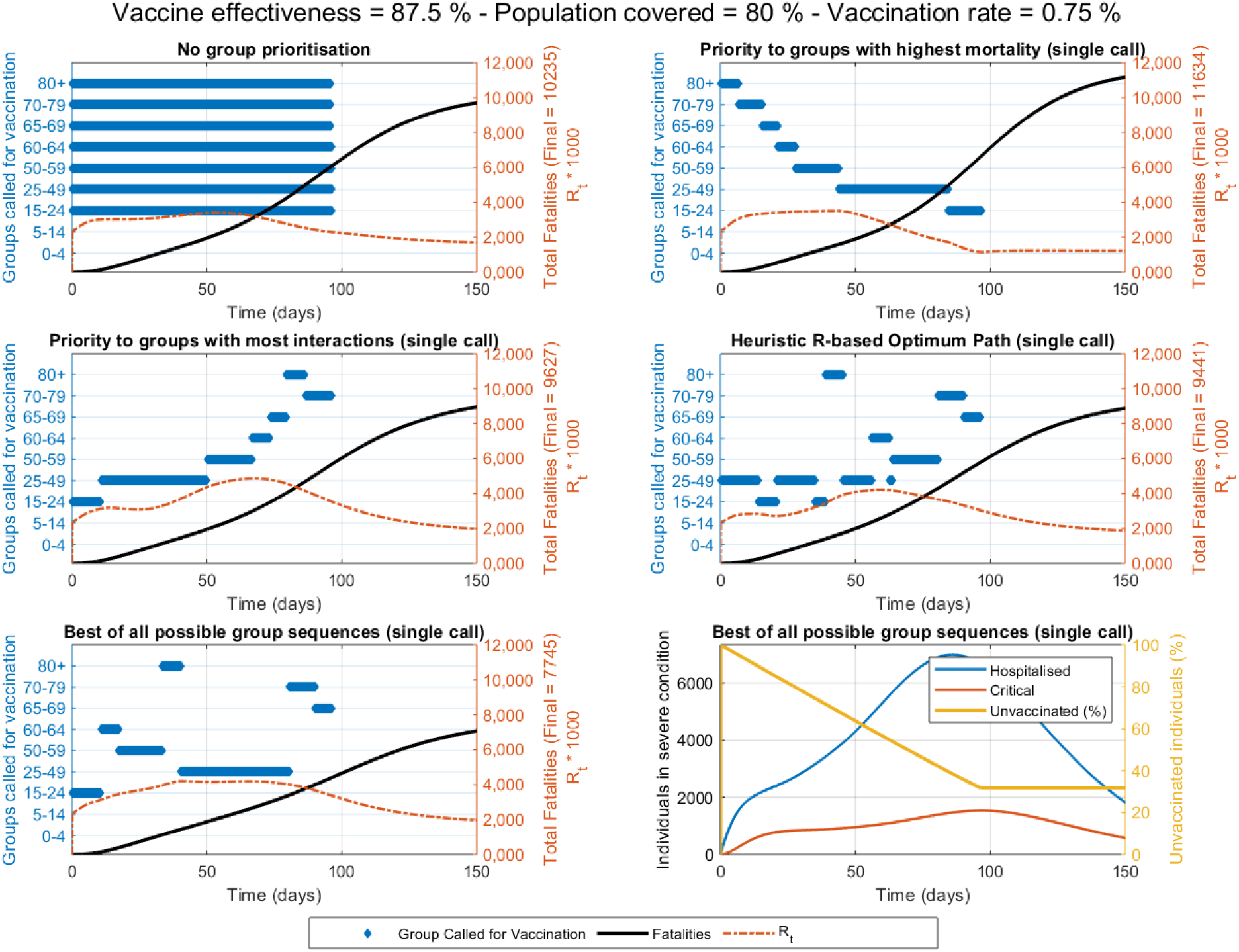
Details of the vaccination strategies of the population with children excluded from vaccination. Active groups called for vaccination, fatalities and R_t_ value over time are shown for each strategy. Case for a daily vaccination rollout rate of 0.75% of the total population, a vaccine effectiveness of 87.5% and a population coverage of 80%. The best result was obtained after (computationally intensive) model evaluation of all possible group sequences. The computationally inexpensive RbPAD strategy is evaluated for weekly cycles and allows for partial calls to specific groups.

Due to the lower number of individuals that can be reached when children are not vaccinated, the total fatalities predicted are higher than for the scenario that includes children in vaccination. In addition, the differences in fatalities between strategies become much smaller. For this scenario, the relative difference between prioritisation by mortality respect to no prioritisation is just 10-17% and it appears that no prioritisation is equal or better than prioritisation by mortality.

The prioritisation by interactions appears still superior than no prioritisation. The relative differences between strategies, however, are much smaller than if children were vaccinated. The largest relative differences (approximately an improvement of 9-12%) appear to occur when the daily vaccination rate is low (0.25%).

In this scenario with children not vaccinated, the final number of fatalities is substantially larger and the best of all sequences appears far from the other strategies suggesting an important suboptimality of the latter. In addition, due to the unavailability for vaccination for children, the R_t_ values results higher, leading to a longer time to control the pandemic. It is worth noting that for population group coverages of 80%, the total unvaccinated individuals in the population remain around 33% due to the unavailability of the children groups for vaccination.

A side-by-side comparison between prioritisation strategies by mortality or by interactions is shown in Figure 6, for a vaccine effectiveness of 87.5 % and the parameters from Tables S3.1, S5.1 and S5.2., both respect to a no prioritisation strategy. In this case, the prioritisation by mortality appears to be slightly worse than a no prioritisation strategy, only under low daily vaccination rollout (0.1 %) and extremely high coverage (95 %) some reductions in fatalities appear as feasible (Figure 6, left).

**Figure 6.**
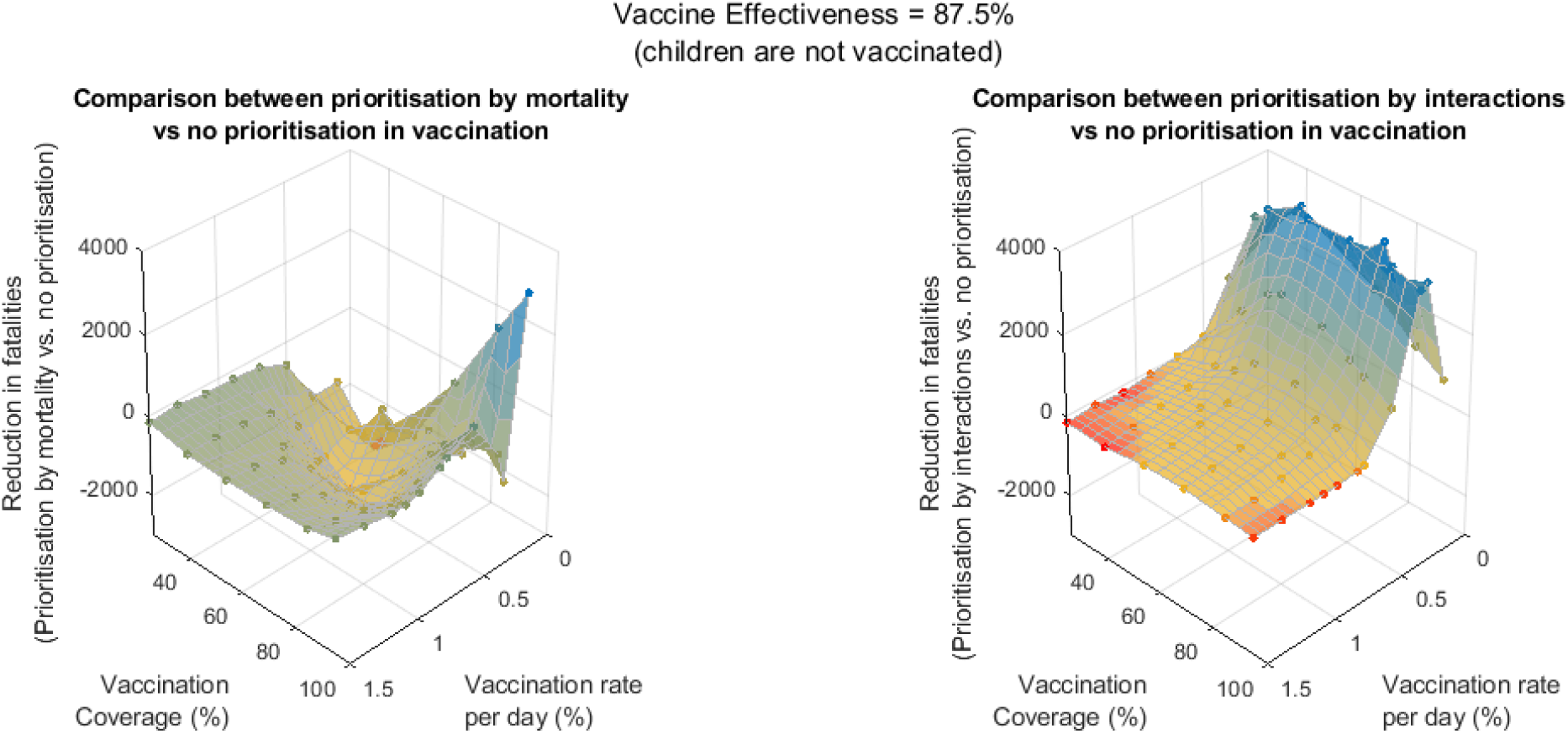
Reduction in fatalities if prioritisation by mortality (left) or by interactions (right) is used instead of a no prioritisation strategy, for different rates of vaccination rollout and coverage. Case for a vaccine effectiveness of 87.5% with children excluded from vaccination and under the behavioural and epidemiological parameters from Tables S3.1, S5.1 and S5.2 and demographics of Spain.

On the other hand, a prioritisation by interactions appears to reduce fatalities but only at daily vaccination rollout rates below circa 0.5% (Figure 6, right). This reduction is however substantially smaller than the potential reductions if children were eligible for vaccination (Figure 4).

### Impact of behavioural parameters

The model requires epidemiological parameters including the fractions of individuals progressing in severity and recovering, as well as the average times in each disease stage. These parameters have been taken from literature good estimations based on latest data that becomes more and more accurate as we learn more about the SARS-CoV-2 virus. Their values can be updated as new information becomes available but their degree of uncertainty is now relatively small.

The model requires also behaviour-related parameters in order to simulate the infection rate. Two types of these parameters have great impact on simulation results. The first is the number of daily contacts (*ni*) that individuals in each age group have with individuals in each of the other age groups. The second important behavioural set of parameters are the levels of self-protection and awareness (*lpa*), used to estimate the likelihood of infection when a contact between a susceptible and an infectious individual takes place.

In this model these parameters are a single-number account for habits and behaviours such as the use of mask, and the general attitude and awareness to protect oneself and others, and they are therefore the parameters with the largest uncertainty. Although the default values used (*lpa* = 0.75 for younger than 65 and *lpa* = 0.95 for 65 and older) have been estimated such that the R values across the simulations correspond to values that have been reported (see Supplementary Information section IX), the uncertainty in these *lpa* parameters remains very high. In order to reduce this uncertainty and its propagation to the vaccination strategy comparison results, an analysis of the impact of different levels of self-protection and awareness (*lpa*) between those under 65 years old (the active population) and over 65 years old (retired individuals) was conducted.

For the case of vaccination including children, the differences in total final fatalities between a vaccination priority by interactions vs. by mortality for different levels of self-protection and awareness are shown in Figure 7.

**Figure 7.**
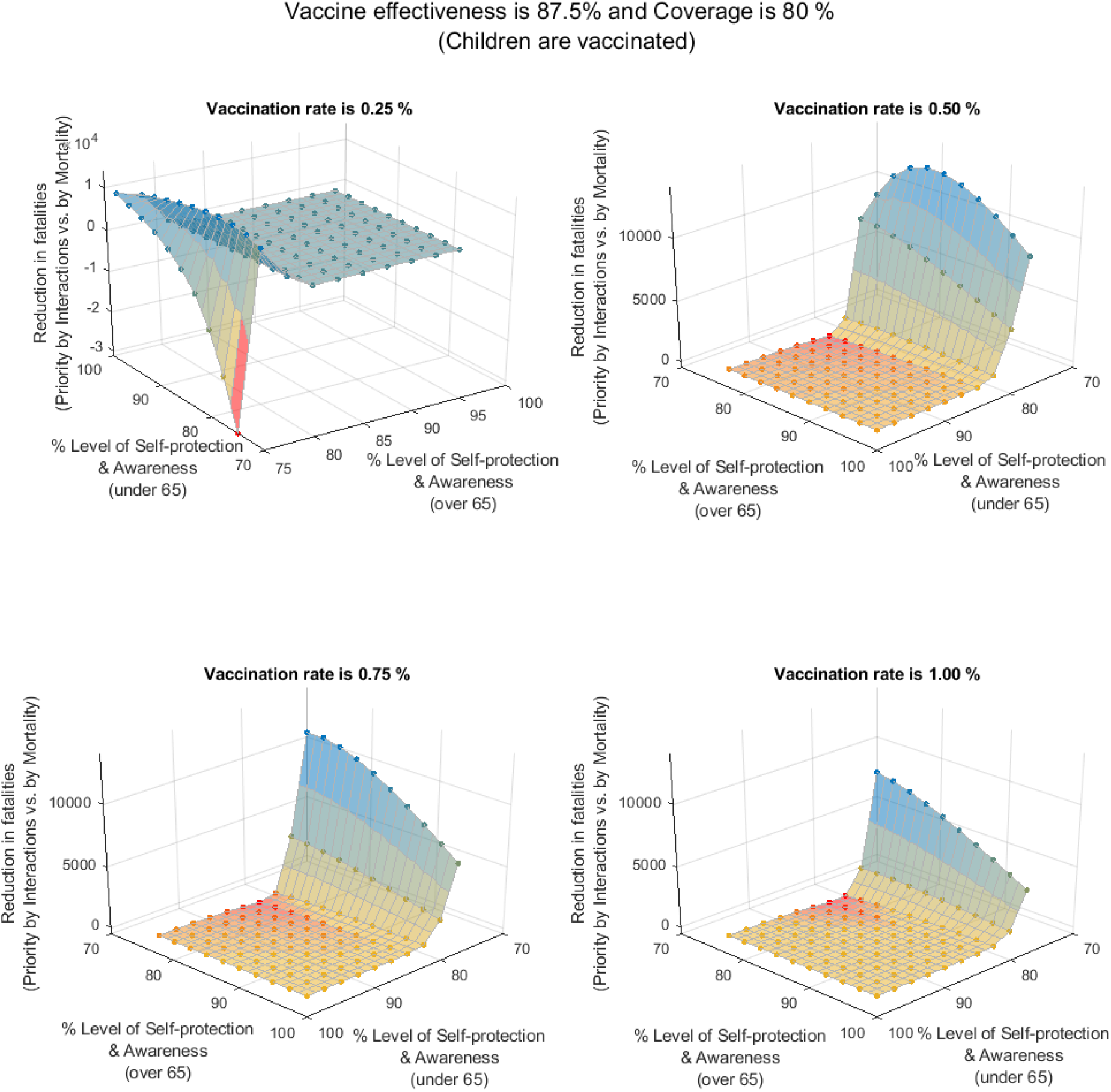
Impact of the different levels of self-protection and awareness (for those under 65 and those over 65 years of age) on the number of possible fatalities that can be avoided if the vaccination is distributed with priority to those with the highest number of daily interactions respect to if it is distributed those with the highest mortality first. Results shown correspond to a vaccination including children, an effectiveness of 87.5%, a population coverage of 80% and demographics for Spain

The results clearly show that a very significant asymmetric potential gain is possible in terms of avoided fatalities if the most interactive groups are vaccinated first when the younger (and more interactive) groups exercise low levels of self-protection and awareness (*lpa*). An opposite result seems to emerge if the daily vaccination rollout is slow (0.25%) and both the younger and the elderly have low levels of self-protection (*lpa*). In this case a priority by mortality seems to achieve a significant reduction in fatalities. The complete results of this analysis respect to the *lpa* parameter for three different vaccine effectiveness, daily vaccination rates and coverage values are shown in Figures S8.1-4 in the Supplementary information.

For the case of vaccination excluding children (since vaccine approvals for children are still pending), the differences in total final fatalities between the two vaccination priority strategies for different *lpa* values are shown in Figure 8.

**Figure 8.**
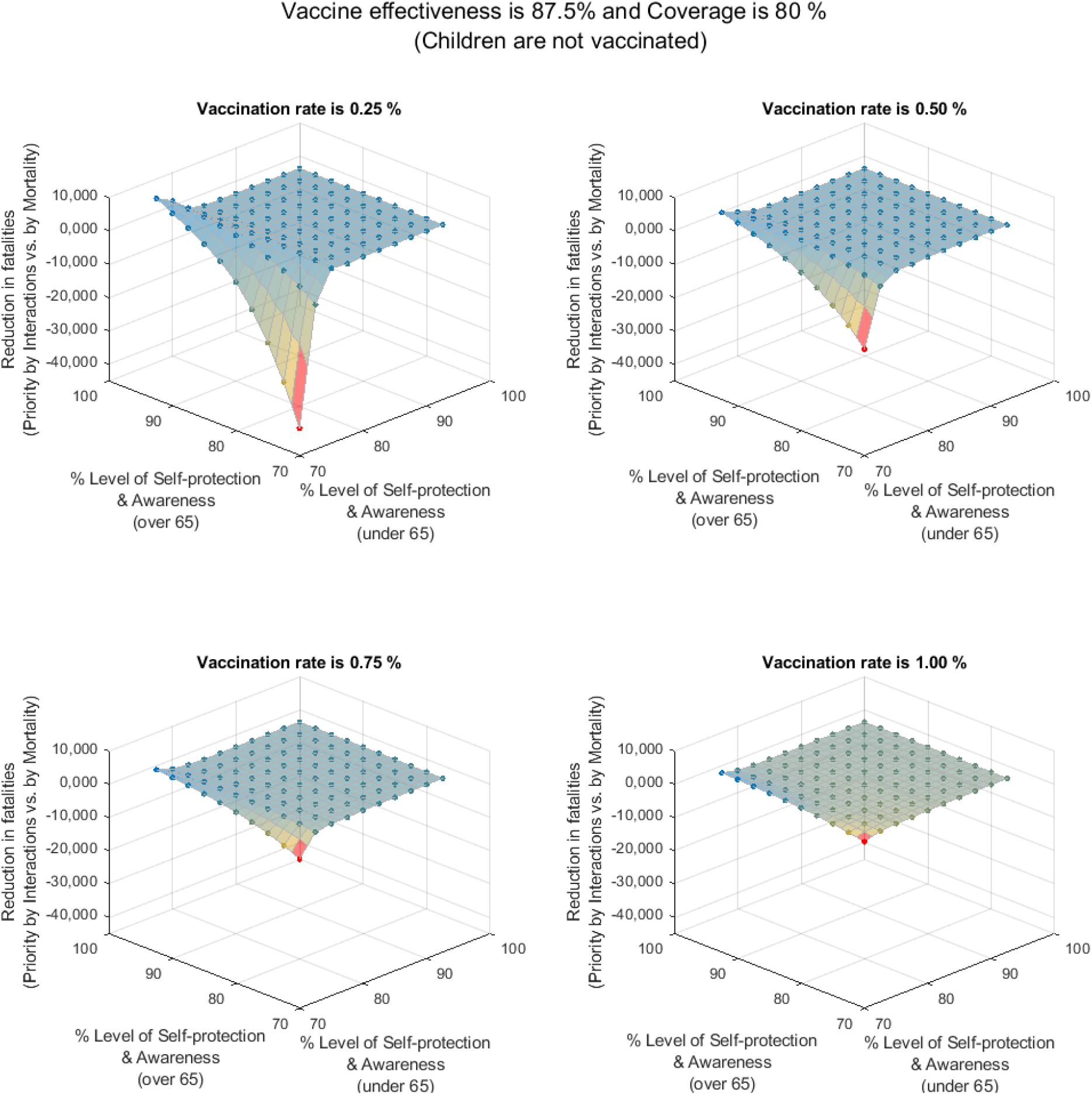
Impact of the different levels of self-protection and awareness (for those under 65 and those over 65 years of age) on the number of possible fatalities that can be avoided if the vaccination is distributed with priority to those with the highest number of daily interactions respect to if it is distributed those with the highest mortality first. Results shown correspond to a vaccination not including children, an effectiveness of 87.5%, a population coverage of 80% and demographics for Spain.

The exclusion of children from the vaccination campaign appears to dissipate the possible benefits of vaccinating with priority by interactions. The advantage of vaccinating the highly interactive groups first is partly lost since children were a key part of that strategy and they are now excluded. Under the scenario in which children are not vaccinated, the differences between strategies become much smaller or almost equivalent if the population groups exercise moderate to high levels of self-protection and awareness (*lpa*). On the other hand, when the levels of self-protection are low, the best strategy seems to favour the vaccination with priority by mortality.

In this scenario with children excluded from vaccination, the levels of self-protection (*lpa*) of the population appear as capable of shifting which one is the most favourable strategy either from priority by interactions or by mortality.

## Conclusions

The impact of different vaccination strategies can be evaluated using a dynamic deterministic model, describing individuals in (age-related) population groups and disease stages. The mechanistic nature of the model allows for the interpretation of the results in order to draw hypotheses and conclusions to inform public health policy.

If everyone, including children, is eligible for vaccination, the model-based comparison of vaccination strategies indicates that a planned vaccine distribution, following prioritised sequences of population groups, can achieve significant reductions in the total final number of fatalities, especially if the younger population is exercising moderate to low levels of self-protection and awareness while the elders exercise a high level of self-protection and awareness. Based on those results (and with the underlying assumption that the vaccine effectiveness applies equally against the transmission of the disease), group prioritisation by those with the highest mortality will not reduce (and may actually increase) the final total fatality count. These results contradict recently announced strategies in several countries and call for a thorough analyses and reconsideration of vaccine distribution strategies against the SARS-CoV-2.

If children remain not eligible for vaccination (due to the lack of approved vaccines against SARS-CoV-2 for children), the comparison of vaccination strategies indicates however that only moderate differences may exist by following one or other prioritisation strategy i.e. by interactions, by mortality or no priority). Differences depend on vaccination rollout rate, coverage and behavioural parameters. The level of self-protection and awareness (*lpa*) exercised by those over 65 year of age during the vaccination campaign appears paramount in order to limit the number of fatalities.

## Contributors

JR conceived, developed the model, designed the experiments and supervised the study. JR and MP analysed the model results, conducted the computational analysis and prepared all figures. JMA, JR and MP collected, processed, analysed and contributed data. JR, MP and JMA drafted the original Article. All authors contributed to the editing of the Article. All authors read and approved the final Article.

## Supporting information

Suplementary Information (Section XI)

## Data Availability

The Matlab source code and the Excel files containing all inputs and parameters used are fully available on demand.

## Acknowledgements

Khalifa University (Grant 8474000317 CRPA-2020-SEHA) and the Government of Abu Dhabi for the funding and support provided.

## Supplementary information

**Section I**. Dynamic COVID-19 model for vaccination (Complete description)

**Section II**. Computation of the dynamic reproduction number (R_t_)

**Section III**. Epidemiological and clinical parameters per age group

**Section IV**. Data sources for the epidemiological and clinical parameters

**Section V**. Behavioural parameters per age group: COVID-19 case study

**Section VI**. R-based projected avoidable deaths (RbPAD) method for vaccination prioritisation

**Section VII**. Compared distribution strategies under limited logistic rate of vaccination (no shortage)

**Section VIII**. Impact of the behavioural parameters of level of self-protection and awareness

**Section IX**. Impact of lpa parameters of R_t_ values.

**Section X**. Sensitivity of the daily number of interactions per group (ni)

**Section XI**. Complete results tables in separate supplementary Excel file.

## Supplementary Section I. Dynamic COVID-19 model for vaccination

The definition of the model dynamic (state) variables is shown in Table S1.1. Each variable corresponds to the number of individuals in that stage in a vector per (age-related) population group (9 activity groups as defined above). Under this structure, each dynamic variable is a vector of dimension 1×9, and the total number of states is a matrix of dimensions 15×9 (15 stages and 9 population groups). Vector variables and parameters are represented in bold font and scalar ones in regular font across all the manuscript.

**Table S1.1.** Model dynamic variables accounting for the number of individuals in each infection stage and group.

| Definition “Number of individuals...” | Vector per group<br>($1 \times 15$ vector) | Total from all groups |
| --- | --- | --- |
| Healthy susceptible to infection | $\mathbf{N}_h$ | $N_{hT}$ |
| Non-infectious asymptomatic | $\mathbf{N}_{ni}$ | $N_{niT}$ |
| Infectious asymptomatic | $\mathbf{N}_{as}$ | $N_{asT}$ |
| Infectious symptomatic | $\mathbf{N}_s$ | $N_{sT}$ |
| Requiring hospitalisation | $\mathbf{N}_{sh}$ | $N_{shT}$ |
| Requiring critical care | $\mathbf{N}_{sc}$ | $N_{scT}$ |
| Recovered & immune | $\mathbf{N}_r$ | $N_{irT}$ |
| Vaccinated still susceptible to infection | $\mathbf{N}_h^v$ | $N_{hvT}$ |
| Vaccinated Non-infectious asymptomatic | $\mathbf{N}_{ni}^v$ | $N_{nivT}$ |
| Vaccinated Infectious asymptomatic | $\mathbf{N}_{as}^v$ | $N_{asvT}$ |
| Vaccinated Infectious symptomatic | $\mathbf{N}_s^v$ | $N_{svT}$ |
| Vaccinated Requiring hospitalisation | $\mathbf{N}_{sh}^v$ | $N_{shvT}$ |
| Vaccinated Requiring critical care | $\mathbf{N}_{sc}^v$ | $N_{scvT}$ |
| Vaccinated Immune | $\mathbf{N}_i^v$ | $N_{ivT}$ |
| Deceased | $\mathbf{N}_d$ | $N_{dT}$ |

### Rates of transition between infection stages

The progression of individuals across stages, as illustrated in Figure 1, includes the impact of vaccination bringing individuals directly into an immune stage or, if ineffectively vaccinated, remaining in their current stage. The transitions between stages are governed by the rates of infection, disease transition and vaccination shown in Table S1.2. All the rates are in vectors with each element corresponding to the rate for each age-related population group.

The average rates of transition between stages are defined as per the latest epidemiological and clinical data and they can be updated as knowledge of the disease increases and treatments improve. These parameters include the proportion of individuals that transition to a more severe stage or recover (Table S1.3) and the average times reported at each stage before transition or recovery (Table S1.4).

**Table S1.2.**
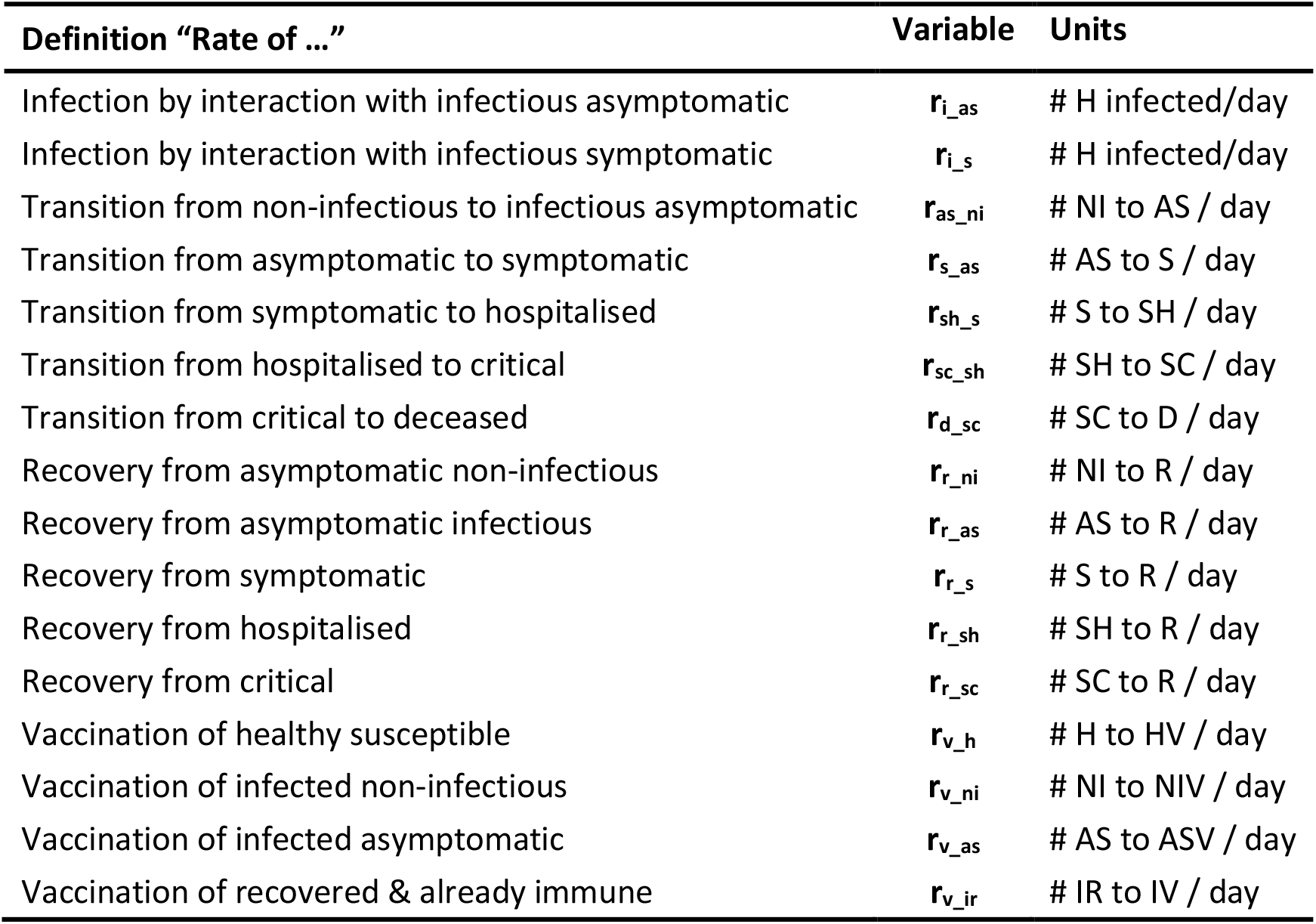
Rates of infection and transition between states in vectors per population group.

| Definition "Rate of ..." | Variable | Units |
| --- | --- | --- |
| Infection by interaction with infectious asymptomatic | $r_{i\_as}$ | # H infected/day |
| Infection by interaction with infectious symptomatic | $r_{i\_s}$ | # H infected/day |
| Transition from non-infectious to infectious asymptomatic | $r_{as\_ni}$ | # NI to AS / day |
| Transition from asymptomatic to symptomatic | $r_{s\_as}$ | # AS to S / day |
| Transition from symptomatic to hospitalised | $r_{sh\_s}$ | # S to SH / day |
| Transition from hospitalised to critical | $r_{sc\_sh}$ | # SH to SC / day |
| Transition from critical to deceased | $r_{d\_sc}$ | # SC to D / day |
| Recovery from asymptomatic non-infectious | $r_{r\_ni}$ | # NI to R / day |
| Recovery from asymptomatic infectious | $r_{r\_as}$ | # AS to R / day |
| Recovery from symptomatic | $r_{r\_s}$ | # S to R / day |
| Recovery from hospitalised | $r_{r\_sh}$ | # SH to R / day |
| Recovery from critical | $r_{r\_sc}$ | # SC to R / day |
| Vaccination of healthy susceptible | $r_{v\_h}$ | # H to HV / day |
| Vaccination of infected non-infectious | $r_{v\_ni}$ | # NI to NIV / day |
| Vaccination of infected asymptomatic | $r_{v\_as}$ | # AS to ASV / day |
| Vaccination of recovered & already immune | $r_{v\_ir}$ | # IR to IV / day |

**Table S1.3.**
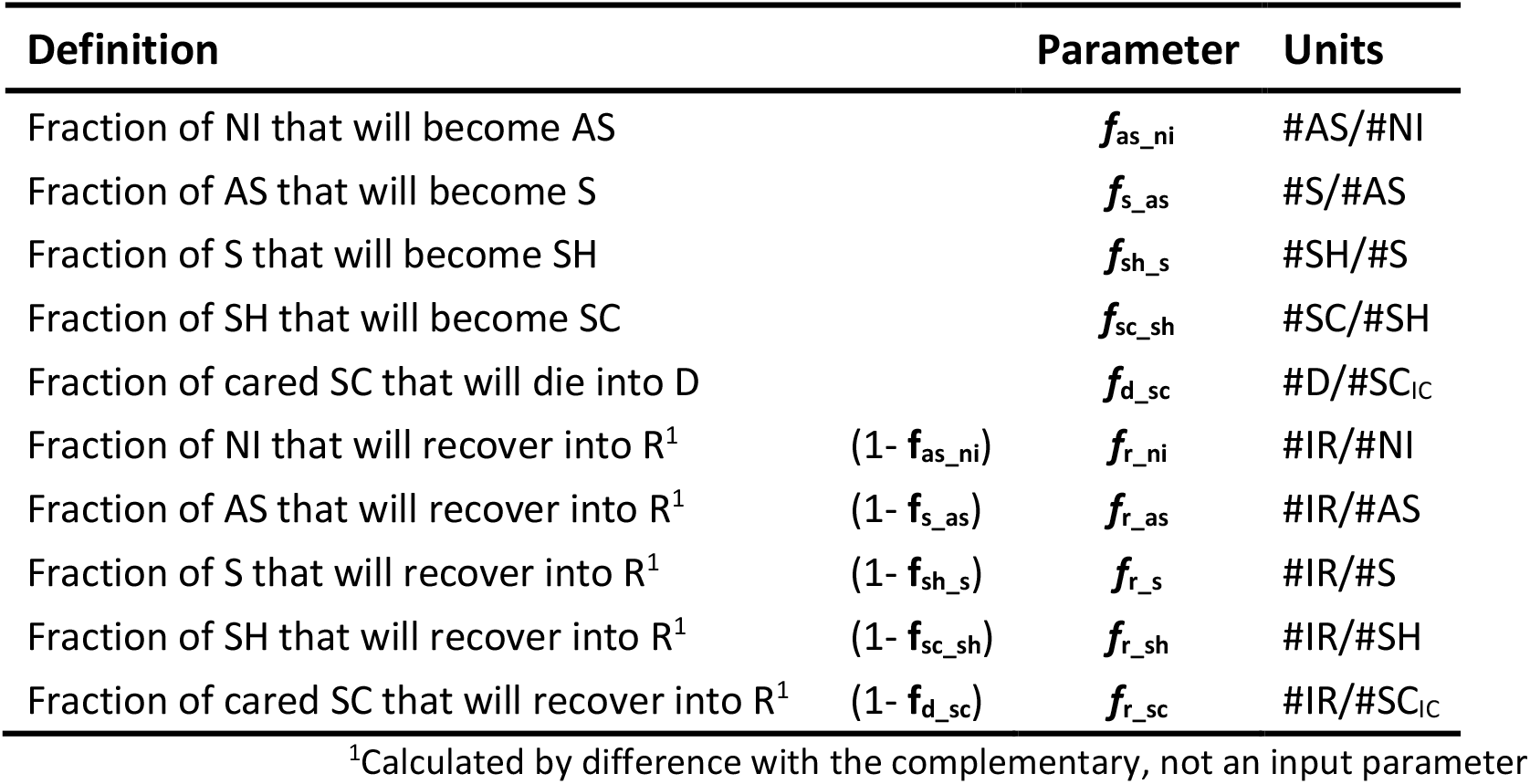
Fractions of individuals progressing through each severity stage per population group.

**Table S1.4.** Clinical average times in each infection stage in vectors per population group.

| Definition | Parameter | Units |
| --- | --- | --- |
| Time to become infectious after infection | $t_{as\_ni}$ | days |
| Time to develop symptoms from becoming infectious | $t_{s\_as}$ | days |
| Time to require hospitalisation from symptoms onset | $t_{sh\_s}$ | days |
| Time to require critical care from hospitalisation | $t_{sc\_sh}$ | days |
| Time to death from critical condition | $t_{d\_sc}$ | days |
| Time to death from critical if no care is available | $t_{d\_nc}$ | days |
| Time to fully recover from asymptomatic non-infectious | $t_{r\_ni}$ | days |
| Time to fully recover from asymptomatic infectious | $t_{r\_as}$ | days |
| Time to fully recover from (non-severe) symptoms | $t_{r\_s}$ | days |
| Time to fully recover from hospitalisation | $t_{r\_sh}$ | days |
| Time to fully recover from critical condition | $t_{r\_sc}$ | days |

The rates of transition between stages (in number of individuals per day) are described in Eqs. A1.a-e. All rates are vectors per age group of dimensions (1×9). Note that the point operators between vectors indicate an element-by-element vector operation.

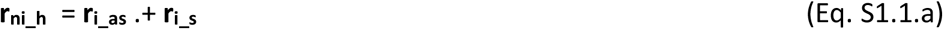

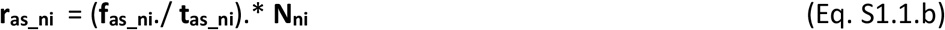

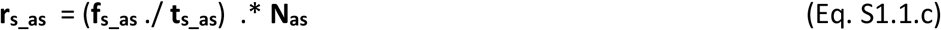

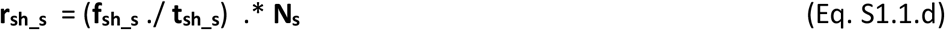

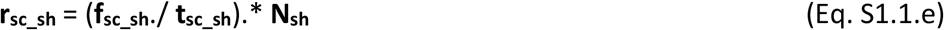

The rates of individuals fully recovering and becoming immune from the different infected stages (in number of individuals per day) are described in Eqs S1.2.a-e. (all rates in vectors per age group).

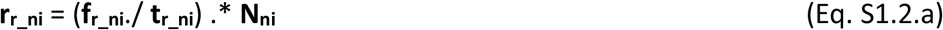

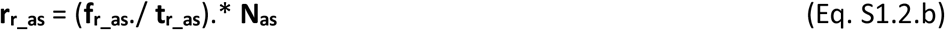

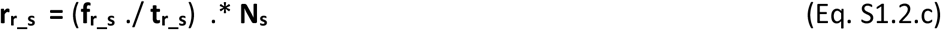

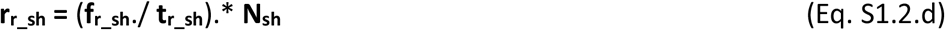

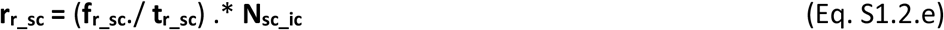

The rate of transition from critical to deceased is the sum of that of those in critical condition receiving intensive care (**r**_**d_scic**_) plus that of those without available care (**r**_**d_scnc**_) as per Eqs. A3.a-c. All critical individuals not receiving intensive care (**N**_**sc_ncc**_) are assumed to become fatalities after a time (**t**_**d_nc**_). A description of the critical care model allocation (in case the ICU capacity limits is reached) is provided below.

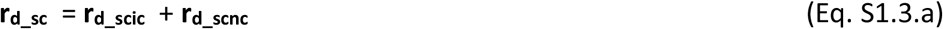

where

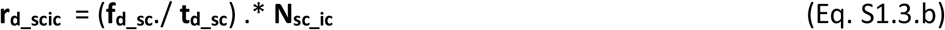

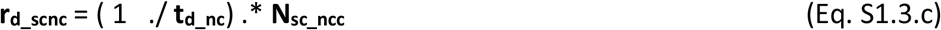

### Allocation of critical care capacity if exceeded

The impact of available critical care capacity is modelled by a specific function to allocate critically ill individuals as per the available ICU. The function allocates critically ill individuals in two possible groups, namely those admitted to ICU (**N**_**sc_ic**_) and those not admitted to ICU due to lack of capacity or for medical or humanitarian reasons (**N**_**sc_ncc**_). At each simulation time point the allocation function is computed for the total **N**_**sc**_ per age group.

The function allocates ICU resources with priority to populations groups with higher ICU survival rate (f_r_sc_) until the maximum number of intensive care units is reached leaving any remaining individuals without care, in this way **N**_**sc_ic**_ and **N**_**sc_ncc**_ are computed.

As the COVID-19 outbreak has progressed, data indicate that not all patients in critical condition have been admitted into intensive care units (ICU). Data show that many individuals with very poor prognosis, particularly those of oldest age may have never been referred to ICU due to capacity limitations or other medical humanitarian reasons. Data from Spain (1) show that for individuals over 70, only a fraction of the reported fatalities previously hospitalised was ever admitted to ICU and this may not be only due to ICU lack of capacity. In order to maintain consistency with the reported data (1) the parameters of f_d_sc_ and f_sc_sh_ have been estimated such that the product of f_d_sc_* f_sc_sh_ (fatality ratios over hospitalised individuals) is consistent with reported numbers for all ages irrespective of reported ICU admissions.

### Rates of infection

The infection of healthy susceptible individuals (H and HV) is modelled as occurring only via their interaction with infectious either asymptomatic (AS) or symptomatic (S) individuals. Hospitalised (SH) and critical (SC) individuals are assumed not available for contacts neither are those deceased (D). Immune individuals after recovery (IR) or effective vaccination (IV) are also not infectious but contribute to the interactive pool of individuals towards herd immunity.

Two rates of infection of healthy susceptible individuals (in number of infections per day) are defined, one from each one of the two possible infecting groups (AS and S). However, in order to serve the goal of determining the optimal sequence of vaccination through population groups, the rates of infection have been expanded into terms for each population group.

The rate of infection of each (age-related) population group i results from the product of the number of healthy susceptible (H) individuals in the group (N_h_^i^) times the sum of the rates of infection from contacts with each one of all the groups. This, for each group, is the product of the average number of daily contacts with individuals of that group (ni_h_^i,j^) times the fraction of those in that group which are infectious (AS or S) times the likelihood of contagion to occur (modelled as function of the use of protection measures e.g. PPE by individuals in *i* and *j* groups) (see Eqs S1.4.a-b).

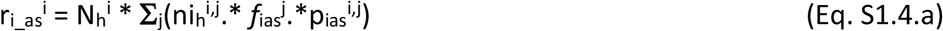

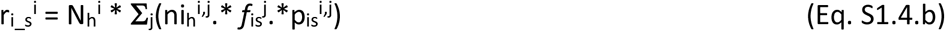

For each (age-related) group, the *average number of daily contacts* an individual in a group *i* has with individuals from each one of the groups *j* (ni_i,j_) are the most important parameters required as an input as they describe the level of social interactivity. These inputs allow for the description of specific key activities in a given group as well as for a complete customisation to the specifics of any community or country. Previous estimations and recent data from contact tracing applications and modelling makes the use of these information possible and reliable in terms of these parameters (2,3). Examples on how these parameters reflect interventions is the opening of schools, which would involve high numbers of daily contacts between children in schools age groups, similarly for secondary school or universities but not necessarily with the other groups. This mapping of contacts is provided via a contacts matrix, in which only the part above the diagonal can be provided as direct input while the part below the diagonal is automatically calculated for consistency between groups based on their relative population sizes. Table S3b shows an example of values arbitrarily assigned and using the demographics of Spain 2019 (4). Interventions such as the *degree of social isolation* as described by (5) remain possible to simulate by modification of these matrix of average daily contacts even at a more detailed level since specific values for group-to-group number of daily contacts can be defined.

The second term impacting the rate of infection of a group *i* is, for each *j* group with which contacts exist, the *proportion of individuals that are infectious* (i.e. AS and S) and still interacting (*f*_ias_^j^ and *f*_is_^j^). This can be directly computed at every time step from the dynamic variables (Eq. S1.5). This computation incorporates the impact of the *awareness of infection* after positive testing (5) via a reduction factor of social interaction for those infected-aware (due to positive testing) or of infected-suspicious individuals. For untested individuals in a group *j* showing symptoms (S), a precautionary self or imposed partial quarantine is captured by the parameter (rfi_s_^j^). For tested individuals, the awareness of infection after a positive result is assumed to lead to a full quarantine and removes those individuals from regular interaction with others. The fractions of infectious AS and S individuals that remain in interaction with others (*f*_ias_ and *f*_is_) are calculated as per Eqs S1.5.a-b. Hospitalised, critical and deceased are considered excluded from the pool of interacting individuals.

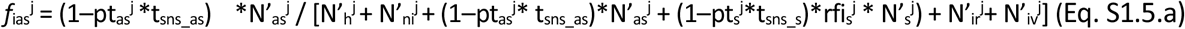

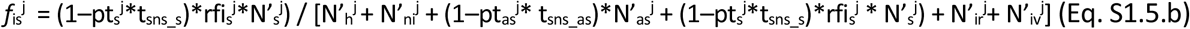

where, N’_x_^j^ means the sum in stage x of both those not vaccinated and those ineffectively vaccinated (N’_x_^j^ = N_x_^j^ + N_xv_^j^) in population group *j*; pt_as_^j^ is the proportion of randomly tested non-symptomatic individuals and pt_s_^j^ is the proportion of symptomatic individuals tested. The parameters t_sns_as_ and t_sns_s_ refer to the sensitivity of the tests for individuals in AS and S stages respectively. The differences are justified since e.g. for non-symptomatic individuals, only RT-qPCR tests are typically assumed adequate while, for symptomatic S individuals, both RT-qPCR and serological tests are used.

The third term impacting the rate of infection within group *i*, is the *likelihood of infection per contact with an infectious* (AS or S) individual of the group *j*. This is an element that could be directly provided in the form of matrix of likelihood of infection between groups (**p**_**i_as**_ and **p**_**i_s**_) without further modelling. If the effect of specific interventions if of interest, the likelihood of infection can be further modelled in more detail as per below.

In this example likelihood of infection between groups is presented modelled as per Eq. S1.6a-b based on the *level of PPE use and awareness* displayed by the two interacting (5) (see Eqs. A6) together with a newly proposed degree of infectiousness for AS and S. The parameters inf_as_ and inf_s_ reflect the degree of infectiousness between 0 and 1. This degree of infectiousness is considered potentially relevant in view not only on possible differences in viral load between AS and S individuals but also of ongoing research regarding specific population groups (e.g. children) which could display different levels of infectiousness to others. The level of protection and awareness of healthy susceptible individuals in the group *i* is described by the parameter lpa_h_^i^ as they interact with infectious AS and S individuals of group *j* with lpa_as_^j^ and lpa_s_^j^ respectively. The values of the parameters can vary between 0 and 1, with 1 corresponding to the use of comprehensive protective measures and zero to the most careless opposite situation. Different values are assigned for the different activity and age-related population groups. (e.g. for primary school children and elderly). In this way, for individuals in group *i*, the likelihood of infection per interaction with individuals of the group *j* (AS and S respectively) is calculated as per Eqs S1.6.a-b.

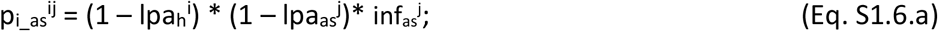

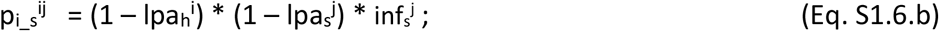

Table S1.5 shows the definitions and units of key variables and parameter used in the calculation of the rate of infection.

**Table S1.5.** Behaviour-related variables and parameters affecting the rate of infection.

| Definition | Parameter | Units |
| --- | --- | --- |
| Average daily contacts per H group $i$ individual with $j$ | $ni_h^{ij}$ | #contacts $j$/ H individual $i$-day |
| Personal protection and awareness by H in group $i^1$ | $lpa_h^i$ | ∅ |
| Personal protection and awareness by AS in group $j^1$ | $lpa_{as}^j$ | ∅ |
| Personal protection and awareness by S in group $j^1$ | $lpa_s^j$ | ∅ |
| Infectiousness level by AS in group $j^1$ | $inf_{as}^j$ | ∅ |
| Infectiousness level by S in group $j^1$ | $inf_s^j$ | ∅ |
| Likelihood of infection by H individual $i$ with AS in $j^2$ | $p_{i\_as}^{ij}$ | infections / $i$ contacting AS in $j$ |
| Likelihood of infection by H individual $i$ with S in $j^2$ | $p_{i\_s}^{ij}$ | infections / $i$ contacting S in $j$ |
| Percentage of tested individuals from AS in group $j^1$ | $pt_{as}^j$ | #random (non S) tested/#(H+NI+AS) |
| Percentage of tested individuals from S in group $j^1$ | $pt_s^j$ | #random S tested/#S |
| Sensitivity of tests used on AS individuals | $t_{sns\_as}$ | #AS detected / #AS tested |
| Sensitivity of tests used on S individuals | $t_{sns\_s}$ | #S detected / #S tested |
| Reduction factor in contacts by S in group $j^1$ | $rfi_s^j$ | ∅ |

<sup>1</sup>Values only within the interval [0,1]; <sup>2</sup>Calculated if modelled or they can be directly provided as input parameters.

### Rate of vaccination

Once an effective vaccination against the SARS-CoV-2 virus becomes available, its mass deployment will take place, at any given moment, at a specific vaccination rate. This rate will be limited either by the availability of vaccine doses or by the system’s capacity to deliver them.

The recipients of the vaccine within the population groups will be all those individuals, either non tested or having tested (truly or falsely) negative or having recovered, that display no symptoms. This will include the vaccinations to the target healthy individuals (H) but also to untargeted individuals such as the non infectious (NI), those unaware (untested or with false negative test) that are asymptomatic infectious (AS) and those that have recovered from the infection (aware or not) (R).

A total capacity vaccination rate is defined (r_v_^T^), in number of individuals that can be vaccinated per day. The vaccination rates of individuals in each of the four possible stages suitable for vaccination, are defined as per Eqs S1.7.a-d in vectors for each (age-related) population group. The rate is proportional to the individuals in each stage and in each group among those suitable and called for vaccination.

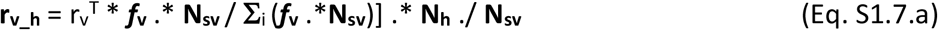

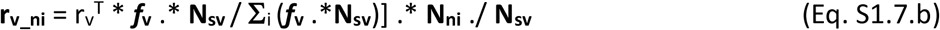

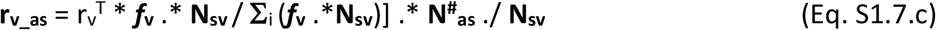

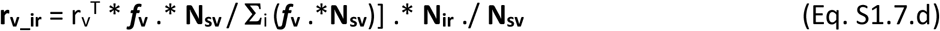

where *f*_v_ is a vector of zeros with one only on the groups currently called for vaccination in a specific moment. **N**^**#**^_**as**_ refers to untested AS individuals therefore unaware of their infection and susceptible of receiving vaccination **N**^**#**^_**as**_ = (1-**pt**_**as**_.**\*** t_sns_as_).\***N**_**as**_.

In a given population group that no distinction is assumed possible between individuals in the vaccination suitable stages without symptoms. Therefore, in a population group *i*, the total number of undistinguishable individuals suitable for vaccination (N_SV_^i^) is given by Eq. S1.7.d.

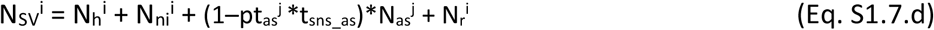

With the above definition, the problem of determining the optimum plan for vaccine application is reduced to the determination, at any given moment in time, of the optimum values of *f*_V_^i^ for each population group, until the vaccination campaign is completed.

### Dynamic transition equations

The dynamic variation on the number of unvaccinated individuals in each stage over time in vectors of (age-related) population groups is governed by the population balance equations Eqs S1.8.a-g.

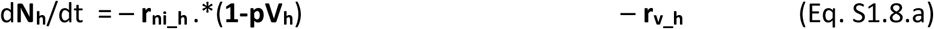

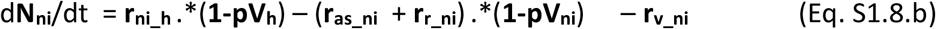

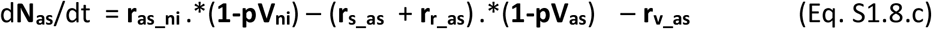

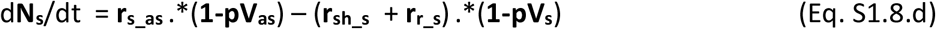

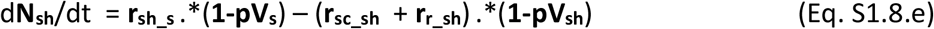

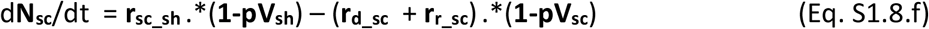

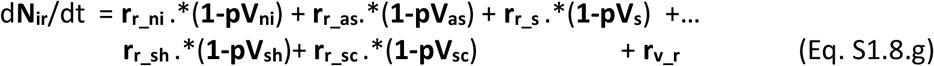

The vaccines once commercialised will likely display a given effectiveness (**eV**) per population group, which will lead to a proportion of ineffective vaccinations. Those ineffectively vaccinated individuals will not acquire immunity and therefore remain susceptible to infection and to all stages of severity. These must however be accounted for separately as they should not be vaccinated again. The population balances for all vaccinated individuals in their possible stages are presented in Eqs S1.8.h-n.

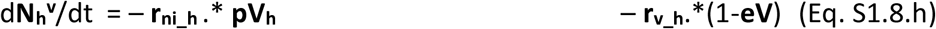

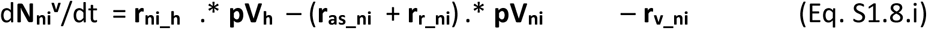

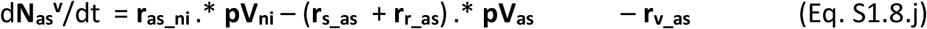

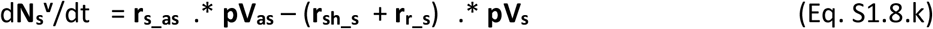

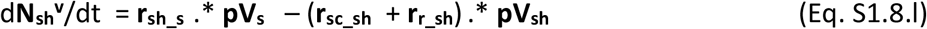

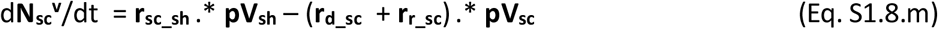

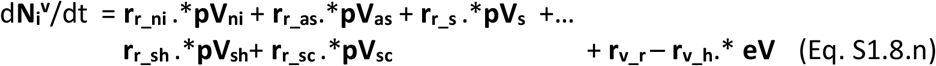

Finally the balance of fatalities is shown in Eq. S1.8.o.

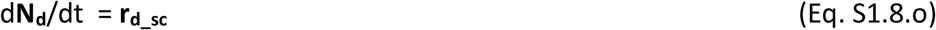

where **pV**_**#**_ is the proportion of individuals in stage # (vector for each population group) that have received a vaccine dose (effective or not).

### Source code

The model source code in MATLAB® and the Excel files containing all inputs and parameters are available on demand.

## Supplementary section II. Computation of the dynamic reproduction number (R_t_

The reproduction number describes the potential infections of susceptible individuals from infected individuals (6). Since the model generates a deterministic set of values for its outputs at any given time, an instantaneous deterministic computation of the reproduction number (R_t_) is possible. Multiple parameters and variables influence the R_t_ such as the duration of the infectious stages; the likelihoods of infection per contact as well as the percentages of individuals transitioning between stages.

The dynamic reproduction number (R_t_) is computed over time according to Eq. S2.1 from the current values of the model state variables. The computation of R_t_ assumes that infectious individuals can only infect others while they are in asymptomatic (AS) and symptomatic (S) stages. Although it has been speculated that post-symptomatic recovered individuals may be infectious for some period of time, this has not been considered at this time given the insufficient data. Hospitalised and critical individuals are assumed not able to infect others in the general population as they are in controlled settings. The provided output of the dynamic reproduction number R_t_ provides additional information that can be used for decision making.

The computation of the R_t_ number is conducted considering that infected individuals can take only three possible infectious paths, namely: (i) Recovery after a period as asymtomatic (AS → R); (ii) Recovery after periods as asymptomatic and symptomatic (AS → S → R) and (iii) Hospitalisation after periods as asymptomatic and symptomatic (AS → S → SH). These paths are made of combinations of four possible infectious stage intervals in which infected individuals spend time and can infect others at their corresponding infection rate (see Table S2.1).

**Table S2.1.** Possible infectious stages intervals for the R_t_ number computation.

| Infectious interval | Fraction of infected passing the interval<br>($\text{ind}_{\text{interv}}/\text{ind}_{\text{inf}}$) | Interval duration<br>(d) | Total infections per stage interval per individual infected<br>($\text{inf}_{\text{interv}}/\text{ind}_{\text{inf}}$) |
| --- | --- | --- | --- |
| $AS \rightarrow R$ | $f_{r\_as}$ | $t_{r\_as}$ | $(r_{i\_asT} / N_{asT}) * t_{r\_as} * f_{r\_as}$ |
| $AS \rightarrow S$ | $(1 - f_{r\_as})$ | $t_{s\_as}$ | $(r_{i\_asT} / N_{asT}) * t_{s\_as} * (1 - f_{r\_as})$ |
| $S \rightarrow R$ | $(1 - f_{r\_as}) * f_{r\_s}$ | $t_{r\_s}$ | $(r_{i\_sT} / N_{sT}) * t_{r\_s} * (1 - f_{r\_as}) * f_{r\_s}$ |
| $S \rightarrow SH$ | $(1 - f_{r\_as}) * (1 - f_{r\_s})$ | $t_{sh\_s}$ | $(r_{i\_sT} / N_{sT}) * t_{sh\_s} * (1 - f_{r\_as}) * (1 - f_{r\_s})$ |

The dynamic computation of R_t_ results from adding the total infection contributions of the four stage intervals as shown in Eq. S2.1.

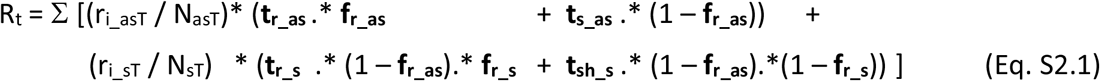

in which, the population group weighted average rates of infection by AS and S are given as per Eqs. S2.2a-b.

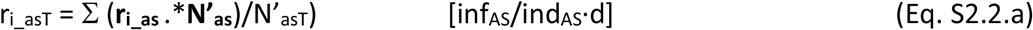

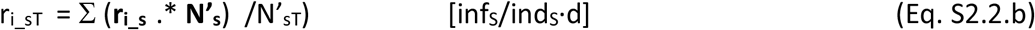

where, N’ _x_^j^ means the sum in stage x of both those unvaccinated and those ineffectively vaccinated (**N’**_**X**_ = **N**_**x**_ + **N**_**xv**_) for all population groups.

## Supplementary section III. Epidemiological and clinical parameters per age group

**Table S3.1.**
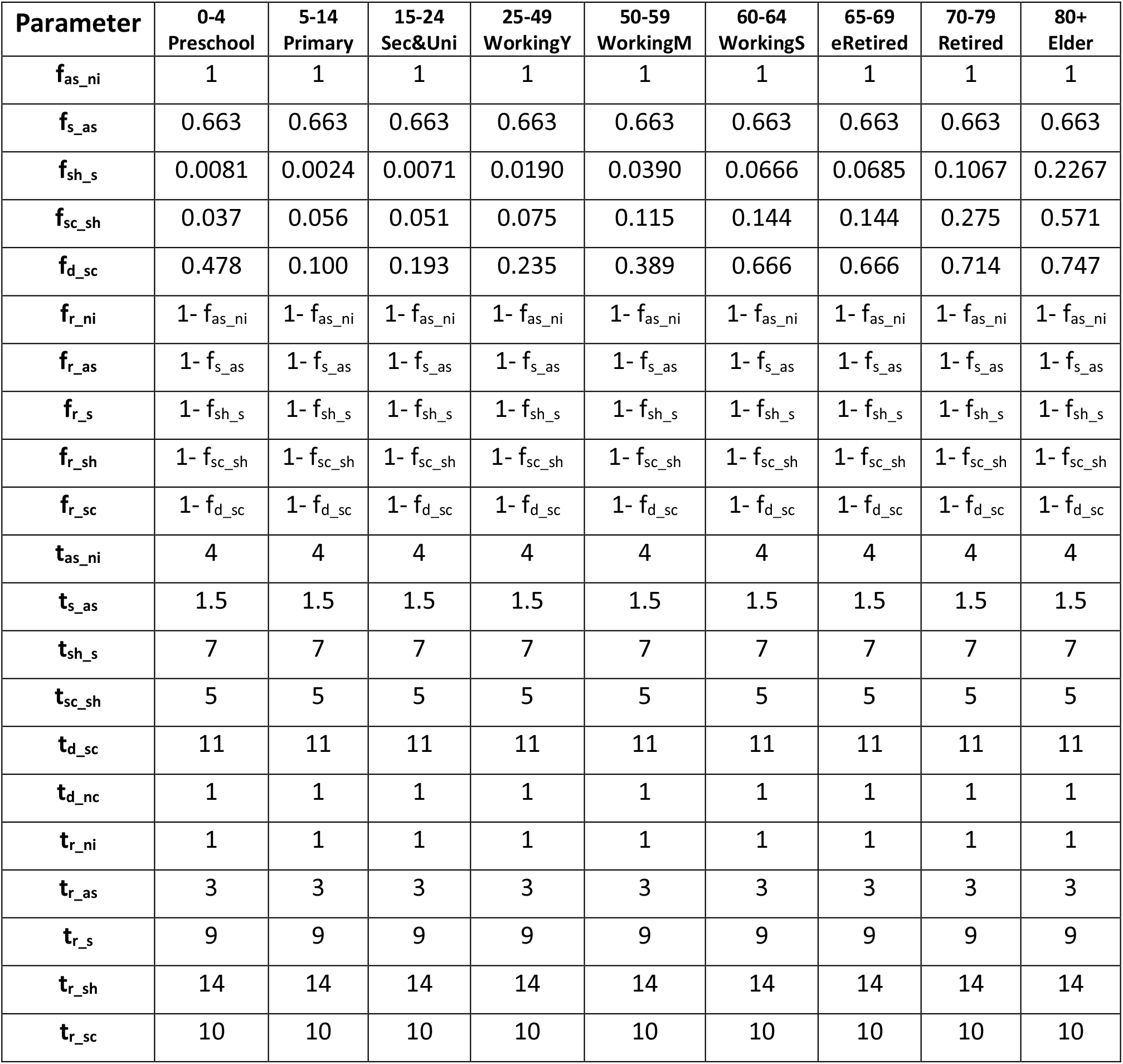
Epidemiological and clinical parameters per (age-related) population group used.

## Supplementary section IV. Data sources for the epidemiological and clinical parameters

**Table S4.1.** Data sources and level of confidence assigned to the epidemiological and clinical parameters from Table S3.1 for the COVID-19 outbreak case study.

| Parameter | Sources and details of estimation | Confidence Level |
| --- | --- | --- |
| $f_{as\_ni}$ | Estimated (100%) | L |
| $f_{s\_as}$ | (7,8) | H |
| $f_{sh\_s}$ | (1,7) | M |
| $f_{sc\_sh}$ | (1) | M |
| $f_{d\_sc}$ | (1) | M |
| $f_{r\_ni}$ | Estimated (0%) | M |
| $f_{r\_as}$ | Calculated ($1 - f_{s\_as}$) | M |
| $f_{r\_s}$ | Calculated ($1 - f_{sh\_s}$) | M |
| $f_{r\_sh}$ | Calculated ($1 - f_{sc\_sh}$) | M |
| $f_{r\_sc}$ | Calculated ($1 - f_{d\_sc}$) | M |
| $t_{as\_ni}$ | Estimated by personal communication | L |
| $t_{s\_as}$ | (9) | L |
| $t_{sh\_s}$ | (10,11) | M |
| $t_{sc\_sh}$ | (11) | M |
| $t_{d\_sc}$ | Personal communications | M |
| $t_{d\_nc}$ | Estimated as one day | M |
| $t_{r\_ni}$ | Estimated by analogy | VL |
| $t_{r\_as}$ | Estimated by analogy | VL |
| $t_{r\_s}$ | Estimated by analogy | L |
| $t_{r\_sh}$ | (11) | M |
| $t_{r\_sc}$ | Personal communications | M |

## Supplementary section V. Behavioural parameters per age group: COVID-19 case study

**Table S5.1.** Estimated average number of daily interpersonal contacts per individual in each population group (row) with individuals from all the groups (columns) used*.

|  | 0-4<br>Preschool | 5-14<br>Primary | 15-24<br>Sec&Uni | 25-49<br>WorkingY | 50-59<br>WorkingM | 60-64<br>WorkingS | 65-69<br>eRetired | 70-79<br>Retired | 80+<br>Elder |
| --- | --- | --- | --- | --- | --- | --- | --- | --- | --- |
| 0-4<br>Preschool | 4.68 | 2.04 | 0.43 | 4.89 | 1.15 | 0.47 | 0.53 | 0.47 | 0.36 |
| 5-14<br>Primary | 0.86* | 13.76 | 1.95 | 1.14 | 1.13 | 0.29 | 0.41 | 0.44 | 0.33 |
| 15-24<br>Sec&Uni | 0.19 | 2.04 | 12.87 | 2.91 | 2.03 | 0.44 | 0.12 | 0.28 | 0.21 |
| 25-49<br>WorkingY | 0.60 | 0.33 | 0.81 | 9.47 | 2.31 | 0.75 | 0.46 | 0.49 | 0.37 |
| 50-59<br>WorkingM | 0.34 | 0.79 | 1.36 | 5.59 | 3.51 | 1.00 | 0.53 | 0.71 | 0.54 |
| 60-64<br>WorkingS | 0.34 | 0.51 | 0.73 | 4.51 | 2.46 | 0.94 | 1.11 | 0.97 | 0.73 |
| 65-69<br>eRetired | 0.45 | 0.83 | 0.24 | 3.24 | 1.54 | 1.30 | 1.21 | 0.79 | 0.60 |
| 70-79<br>Retired | 0.25 | 0.56 | 0.34 | 2.17 | 1.29 | 0.71 | 0.50 | 0.72 | 0.55 |
| 80+<br>Elder | 0.25 | 0.56 | 0.34 | 2.17 | 1.29 | 0.71 | 0.50 | 0.72 | 0.55 |
| Group<br>Size<sup>#</sup><br>(x10<sup>-3</sup>) | 2,039 | 4,847 | 4,628 | 16,752 | 6,905 | 2,799 | 2,399 | 3,794 | 2,865 |

\*Values based on (2); Shaded cells are forced calculated for consistency in total numbers of contacts considering the different numbers of individuals in each group. <sup>#</sup>2019 population demographics of Spain is used (4).

**Table S5.2.** Behavioural and other infectivity relevant parameter as selected for the case study

| Parameter* | 0-4<br>Preschool | 5-14<br>Primary | 15-24<br>Sec&Uni | 25-49<br>WorkingY | 50-59<br>WorkingA | 60-64<br>WorkingM | 65-69<br>eRetired | 70-79<br>Retired | 80+<br>Elder |
| --- | --- | --- | --- | --- | --- | --- | --- | --- | --- |
| $lpa_h$ | 0.75 | 0.75 | 0.75 | 0.75 | 0.75 | 0.75 | 0.95 | 0.95 | 0.95 |
| $lpa_{as}$ | 0.75 | 0.75 | 0.75 | 0.75 | 0.75 | 0.75 | 0.95 | 0.95 | 0.95 |
| $lpa_s$ | 0.85 | 0.85 | 0.95 | 0.95 | 0.95 | 0.95 | 0.95 | 0.95 | 0.95 |
| $inf_{as}$ | 0.75 | 0.75 | 0.75 | 0.75 | 0.75 | 0.75 | 0.75 | 0.75 | 0.75 |
| $inf_s$ | 1 | 1 | 1 | 1 | 1 | 1 | 1 | 1 | 1 |
| $rfi_s$ | 0.1 | 0.1 | 0.25 | 0.25 | 0.25 | 0.25 | 0.1 | 0.1 | 0.1 |
| $pt_{as}$ | 0 | 0 | 0 | 0 | 0 | 0 | 0 | 0 | 0 |
| $pt_s$ | 0 | 0 | 0 | 0 | 0 | 0 | 0 | 0 | 0 |
| $t_{sns\_as}$ | 0.8 | 0.8 | 0.8 | 0.8 | 0.8 | 0.8 | 0.8 | 0.8 | 0.8 |
| $t_{sns\_s}$ | 0.8 | 0.8 | 0.8 | 0.8 | 0.8 | 0.8 | 0.8 | 0.8 | 0.8 |

\*Rationale: The default level of personal protection and awareness ( $lpa$ ) in those older than 65 is taken as higher than the younger population; Adult symptomatic individuals are expected to take higher level of personal protection and awareness ( $lpa_s$ ) on suspicion or confirmation of COVID infection. No reduction factor of their social interactivity respect to healthy ones is applied for asymptomatic infected individuals as they are ignorant of their condition; Symptomatic infected individuals are expected to reduce their social interactivity respect to healthy ones as they feel sick ( $rfi_s < 1$ ); to facilitate the interpretations of results the vaccine optimisation example was in absence of testing; the sensitivities of tests, when active, can be made different for AS and for S if information is available.

## Supplementary section VI. R-based projected avoidable deaths (RbPAD) method for vaccination prioritisation

A strategy based on the estimation the projected avoidable deaths (PAD) by vaccination in each group was developed. The strategy does not require a model to be applied and it establishes the vaccination priority at any given time on the group with the highest PAD by vaccination. The avoided deaths are accounted from two sources and not from the group’s own mortality only, but also from that of those the projected secondary and subsequent infections on the entire population avoided by the vaccination in the group. These are estimated and accounted for using the dynamic reproduction number (R_t_) which can be estimated real time by health system data.

Under the RbPAD method, priority is directed at any given moment to the vaccination of the group *i* with the highest number of projected avoidable deaths per vaccination (see Eq. 1). This is calculated as a function of three elements, namely (i) the risk of infection in that group (as the ratio of the present rate of infections and the number of individuals in the group); (ii) the mortality per infection in the group (f_d_ni_^i^) (from epidemiological data) and (iii) the mortalities of the projected secondary and subsequent infections avoided that a vaccinated group member would have inflicted on the entire population.

If the R_t_ number is used as the estimator of infection propagation at a given moment, the last term of projected secondary infections can be estimated if group-specific detailed information about the ongoing R_t_ number is available such that a matrix of R numbers between groups (R_M_) can be built. This matrix is then projected to any number of infection cycles (n) by powering the matrix to n.

All three terms can in principle be measured or estimated from actual data directly collected by the health systems in a given community. Although the RbPAD strategy is demonstrated below on the dynamic model, its practical implementation does not require or rely on any model, partly decoupling the outcome sequence of priority groups from possible model shortcomings and uncertainties.

The calculation of the projected avoidable deaths for all population group*s* (**PAD**_**v**_), in terms of the model variables, is presented in Eq. 1 (see also Appendix). Eq. 1 was used to evaluate the RbPAD method against the alternatives for the case study example for Spain. A value of three infection cycles for projected propagation (n=3) was used.

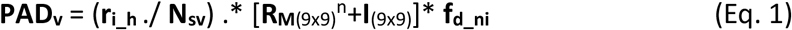

where **I**_(9×9)_ is the identity matrix of the indicated size to account for each group’s own mortalities.

The RbPAD methods was applied in conjunction the dynamic model and it is computationally inexpensive as it is evaluated in one single model simulation that follows the defined optimum path. There is no requirement for repeated model simulations to evaluate the objective function for changes in vaccination sequence. The computational low cost allows for its use in conjunction with sensitivity and Monte Carlo simulations to address uncertainty in model parameters. This also allows for the RbPAD method to be deployed together with the model on low-cost web-based platforms, making a vaccination optimisation method available globally at no cost.

## Supplementary section VII. Vaccine distribution strategies

Compared vaccination strategies showing active groups called for vaccination, fatalities and R_t_ value over time for the case study with data and demographics from Spain. Different daily vaccination rates (0.25 and 1.25 of the total population per day), vaccine effectiveness (75 and 95%) and population coverage (25 and 80%) are shown. The best result was obtained after (computationally intensive) evaluations of all possible group sequences (assuming only one single call per group). The computationally inexpensive R-based projected avoidable deaths method is evaluated for weekly cycles and allows for multiple partial calls to any specific group. The results are shown for scenarios in which children are vaccinated (Figures S7.1-8) and for scenarios in which children are not vaccinated (Figures S7.9-16). Analogously to Figure 3, these figures complement the results shown in the Results and discussion section of the main manuscript in Table 1 (Section VII.A) and Table 2 (Section VII.B).

### A. Comparative evaluation of vaccine distribution strategies when all population can be vaccinated

**Figure S7.1.**
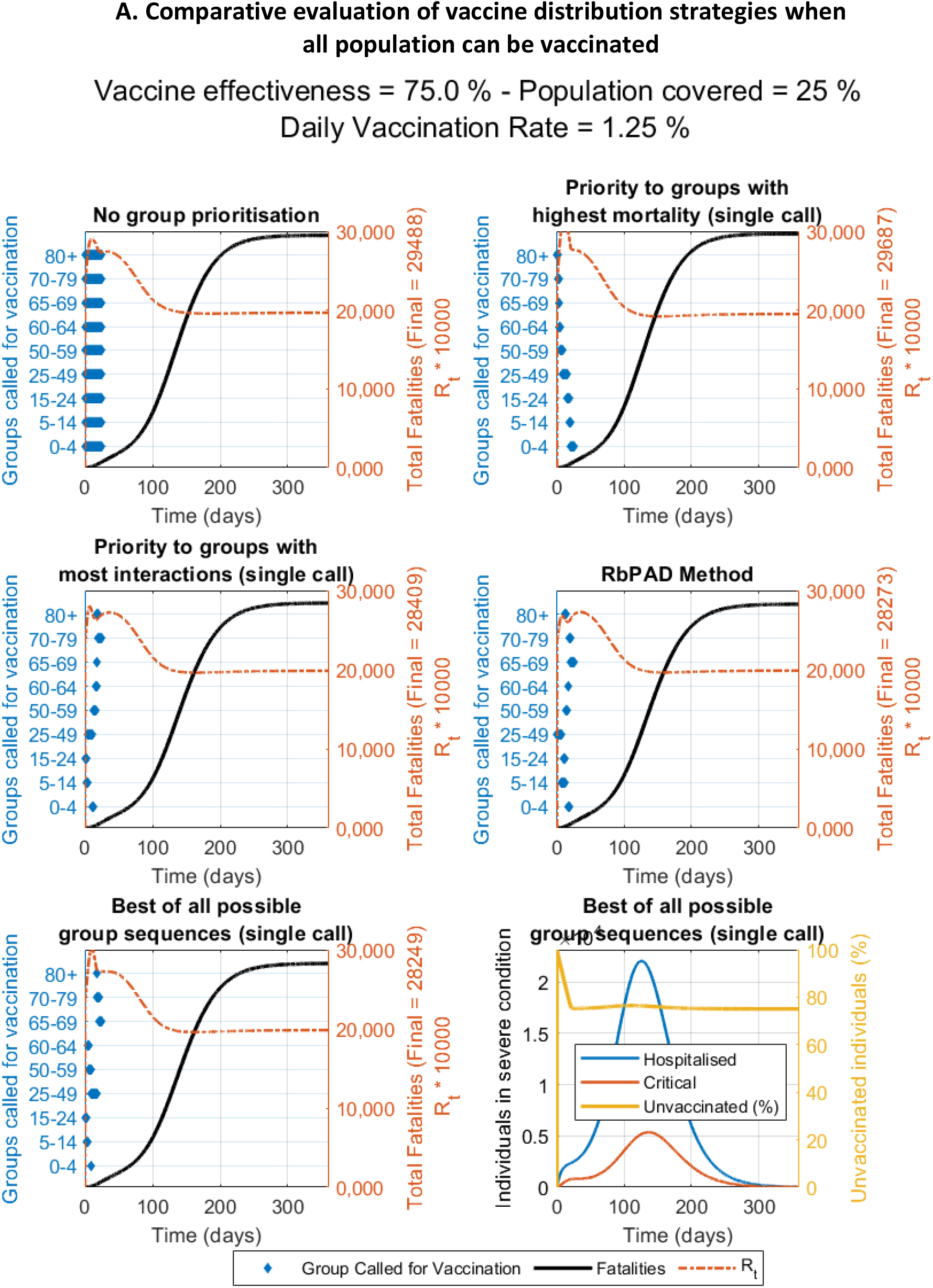
Compared vaccination strategies showing active groups called for vaccination (in blue bars), fatalities (thick black line) and R_t_ value over time (dotted orange line) for the case study with data and demographics from Spain. Constant vaccination rate is set at 1.25% of the total population per day, vaccine effectiveness of 75% and population coverage of 25% as shown.

**Figure S7.2.**
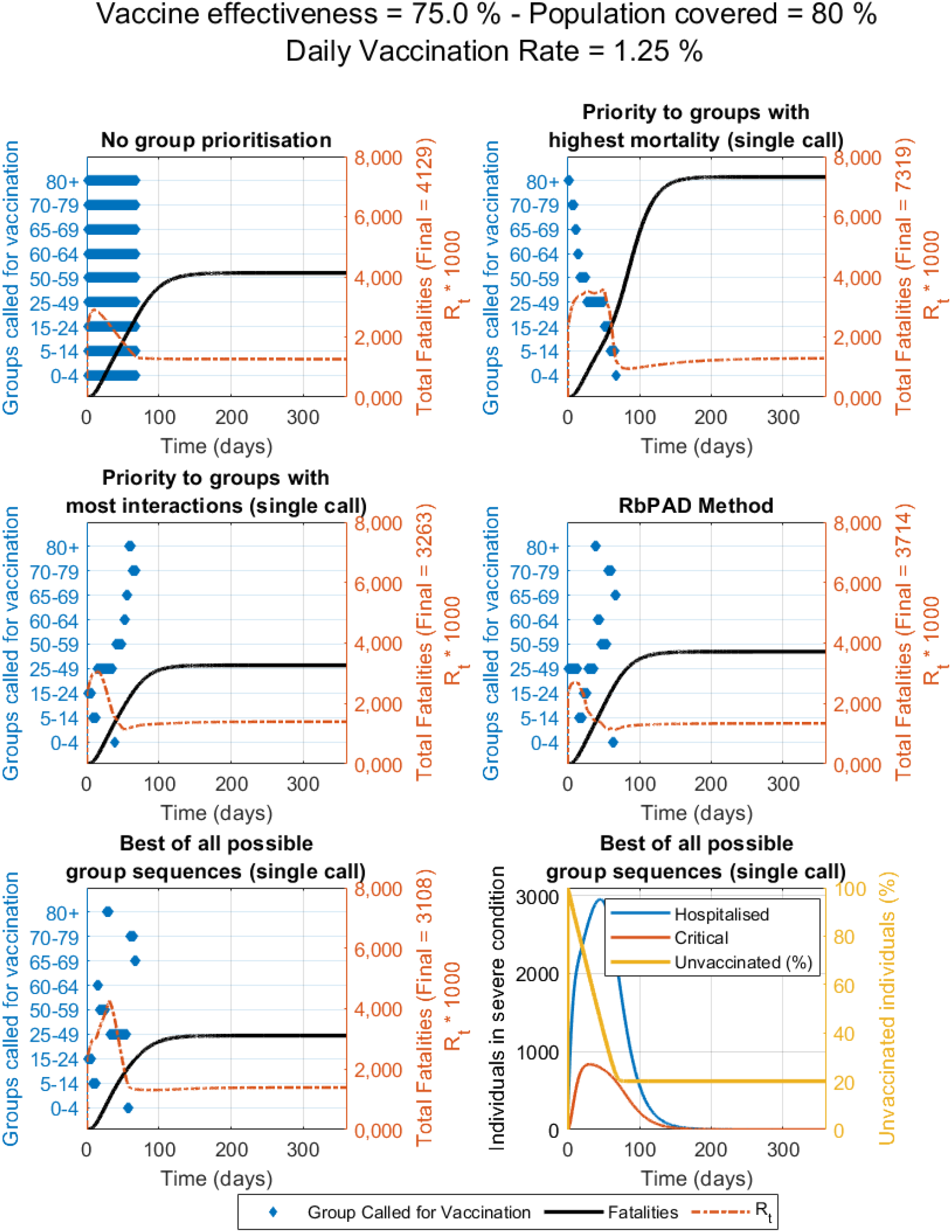
Compared vaccination strategies showing active groups called for vaccination (in blue bars), fatalities (thick black line) and R_t_ value over time (dotted orange line) for the case study with data and demographics from Spain. Constant vaccination rate is set at 1.25% of the total population per day, vaccine effectiveness of 87.5% and population coverage of 80% as shown.

**Figure S7.3.**
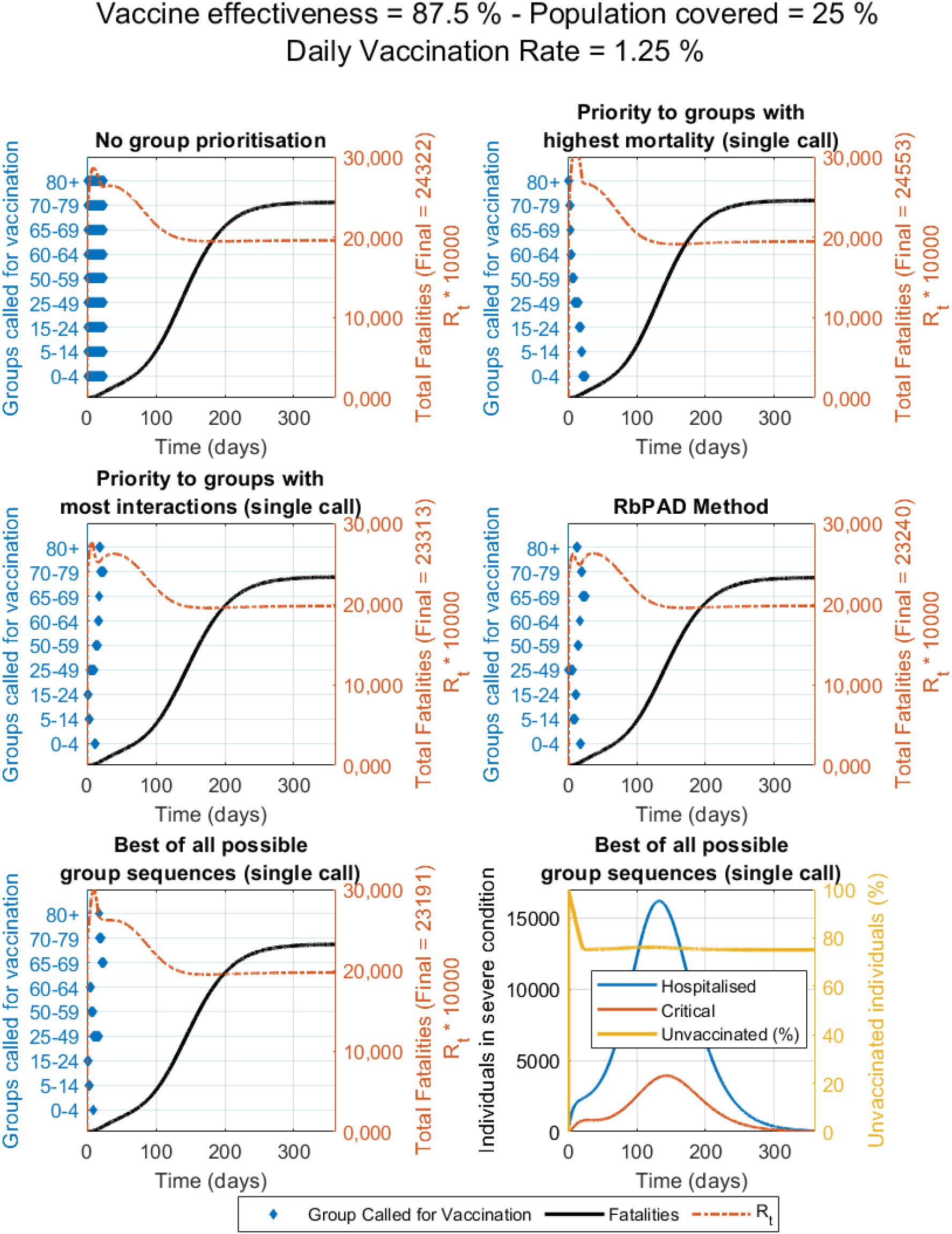
Compared vaccination strategies showing active groups called for vaccination (in blue bars), fatalities (thick black line) and R_t_ value over time (dotted orange line) for the case study with data and demographics from Spain. Constant vaccination rate is set at 1.25% of the total population per day, vaccine effectiveness of 87.5% and population coverage of 25% as shown.

**Figure S7.4.**
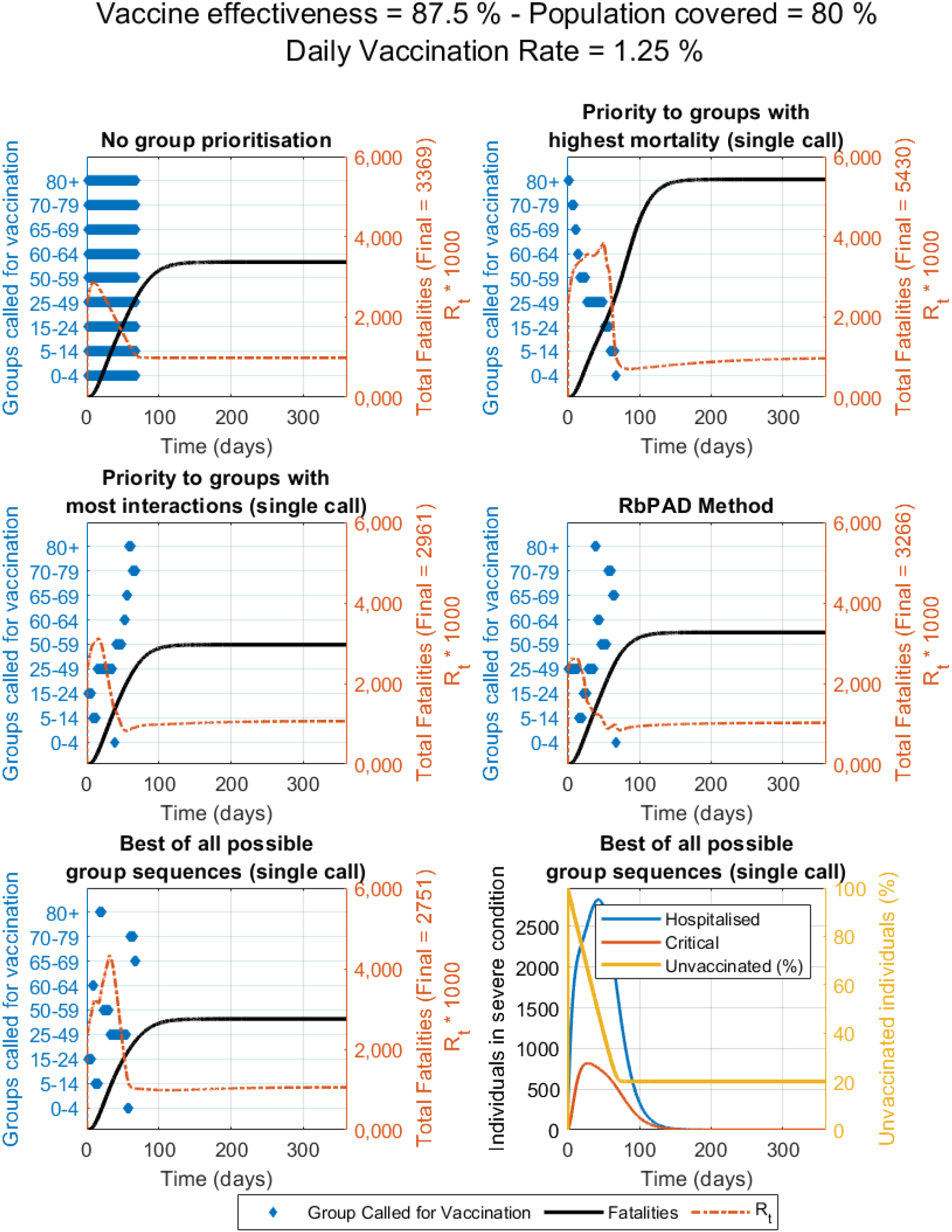
Compared vaccination strategies showing active groups called for vaccination (in blue bars), fatalities (thick black line) and R_t_ value over time (dotted orange line) for the case study with data and demographics from Spain. Constant vaccination rate is set at 1.25% of the total population per day, vaccine effectiveness of 87.5% and population coverage of 80% as shown.

**Figure S7.5.**
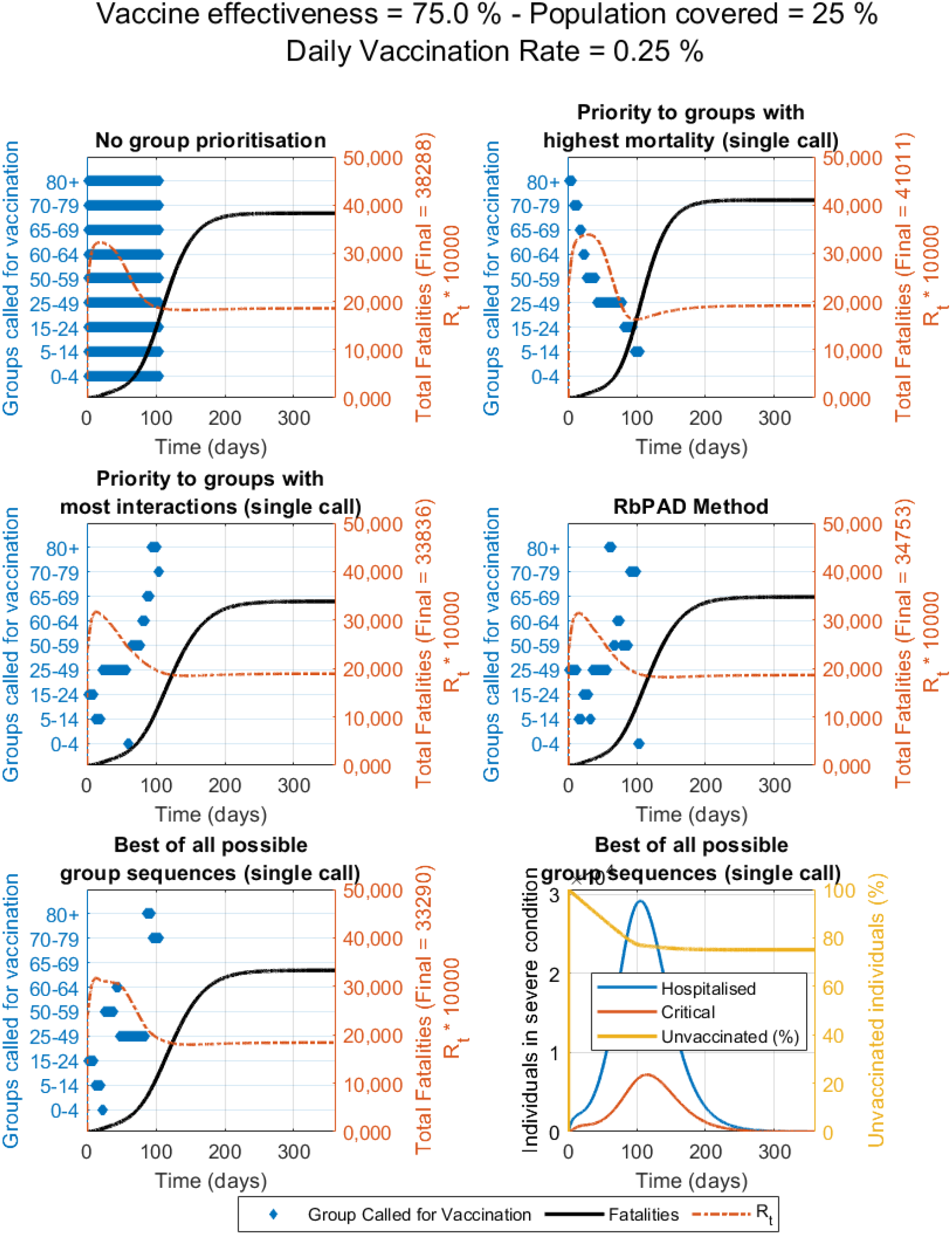
Compared vaccination strategies showing active groups called for vaccination (in blue bars), fatalities (thick black line) and R_t_ value over time (dotted orange line) for the case study with data and demographics from Spain. Constant vaccination rate is set at 0.25% of the total population per day, vaccine effectiveness of 75% and population coverage of 25% as shown.

**Figure S7.6.**
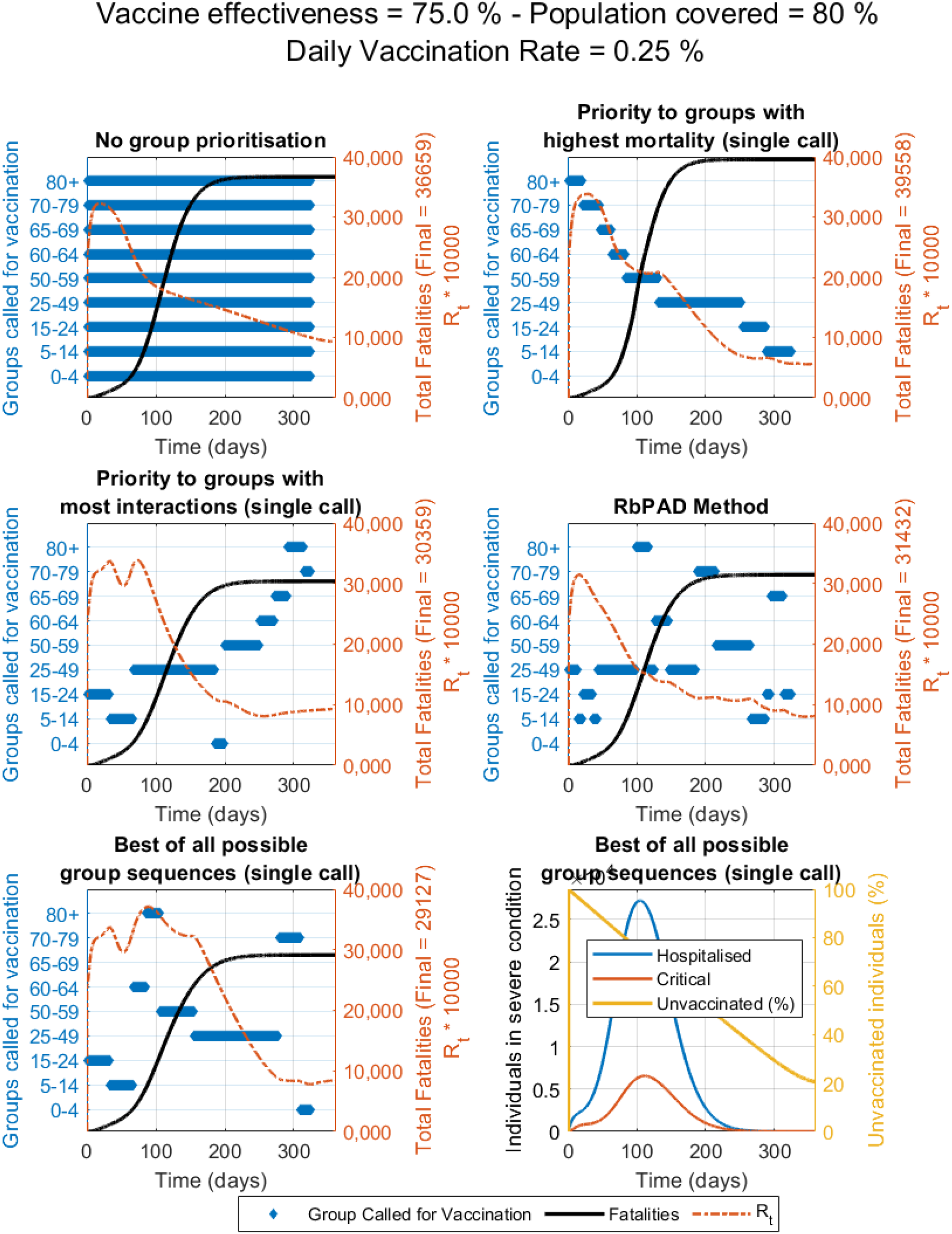
Compared vaccination strategies showing active groups called for vaccination (in blue bars), fatalities (thick black line) and R_t_ value over time (dotted orange line) for the case study with data and demographics from Spain. Constant vaccination rate is set at 0.25% of the total population per day, vaccine effectiveness of 75% and population coverage of 80% as shown.

**Figure S7.7.**
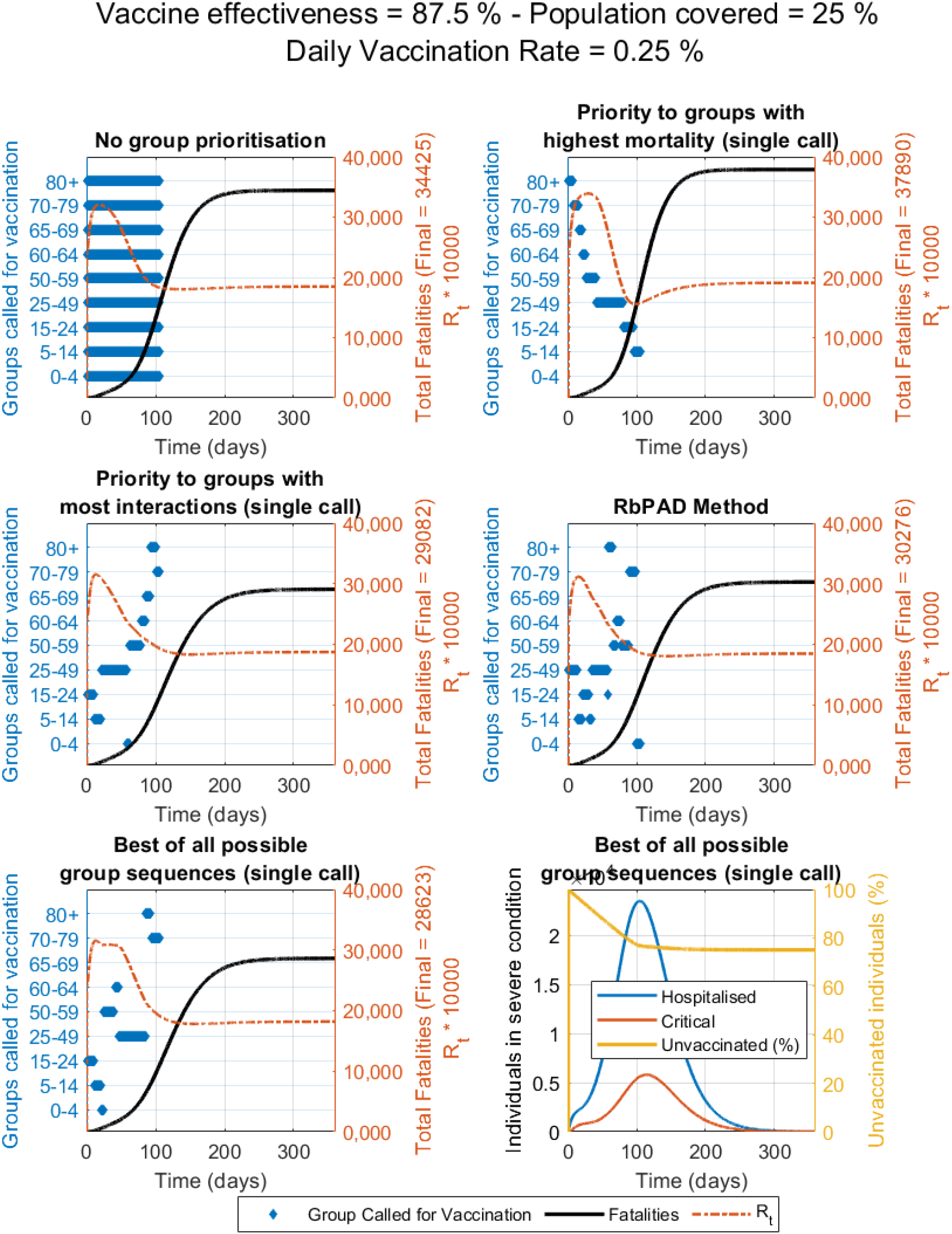
Compared vaccination strategies showing active groups called for vaccination (in blue bars), fatalities (thick black line) and R_t_ value over time (dotted orange line) for the case study with data and demographics from Spain. Constant vaccination rate is set at 0.25% of the total population per day, vaccine effectiveness of 87.5% and population coverage of 25% as shown.

**Figure S7.8.**
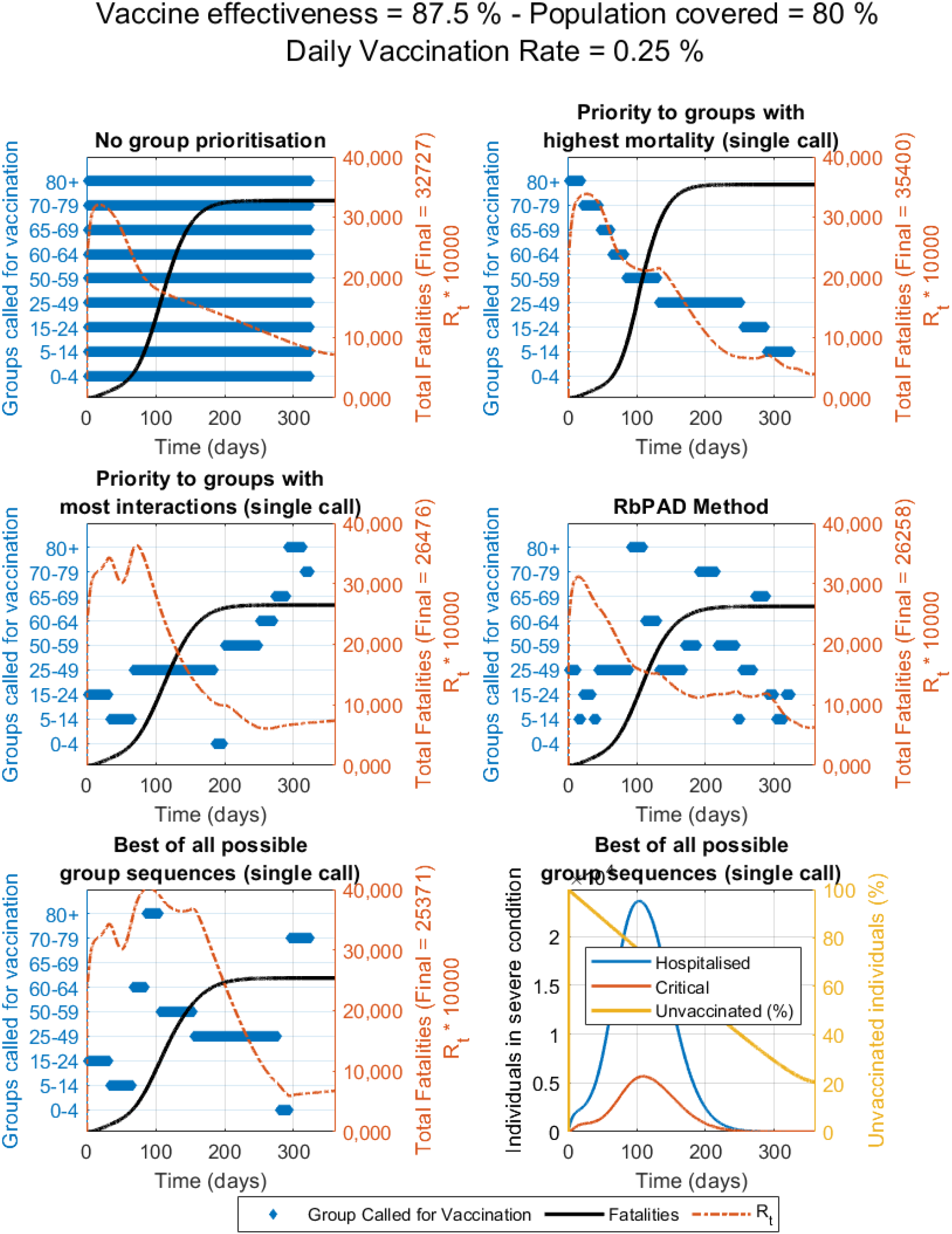
Compared vaccination strategies showing active groups called for vaccination (in blue bars), fatalities (thick black line) and R_t_ value over time (dotted orange line) for the case study with data and demographics from Spain. Constant vaccination rate is set at 0.25% of the total population per day, vaccine effectiveness of 87.5% and population coverage of 80% as shown.

### B. Comparative evaluation of vaccine distribution strategies when children under 16 are not vaccinated

**Figure S7.9.**
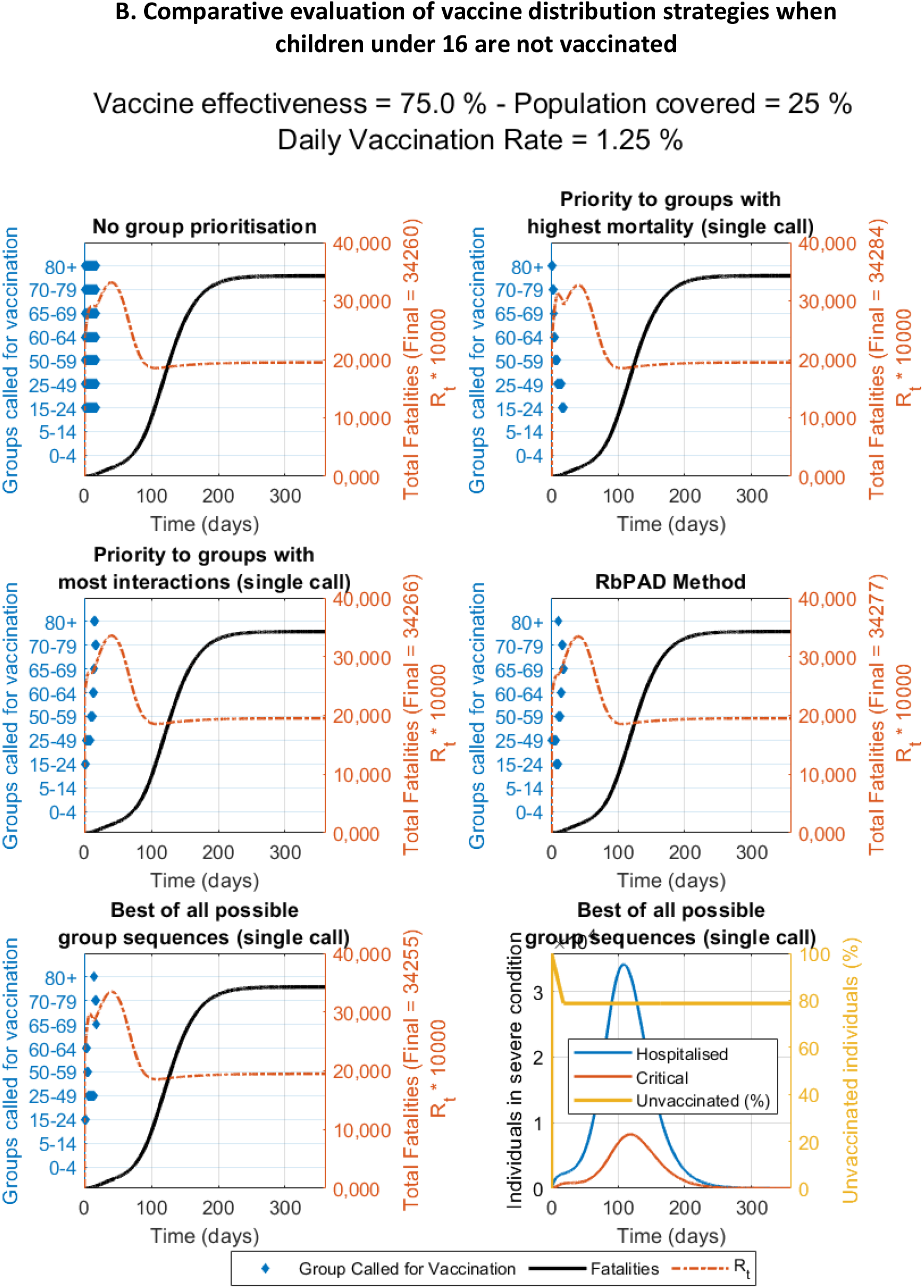
Compared vaccination strategies showing active groups called for vaccination (in blue bars), fatalities (thick black line) and R_t_ value over time (dotted orange line) for the case study with data and demographics from Spain. Constant vaccination rate is set at 1.25% of the total population per day, vaccine effectiveness of 75% and population coverage of 25% as shown.

**Figure S7.10.**
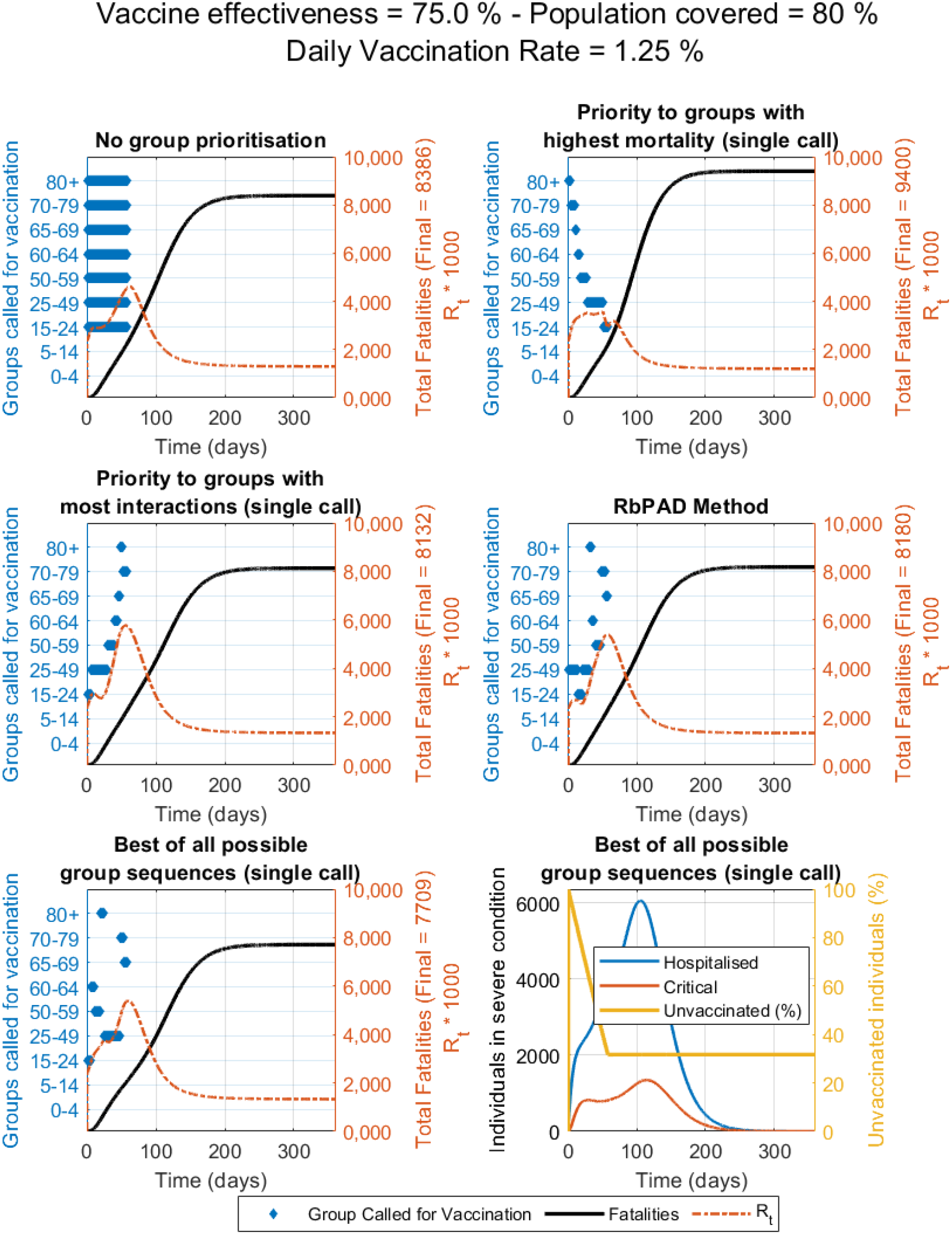
Compared vaccination strategies showing active groups called for vaccination (in blue bars), fatalities (thick black line) and R_t_ value over time (dotted orange line) for the case study with data and demographics from Spain. Constant vaccination rate is set at 1.25% of the total population per day, vaccine effectiveness of 75% and population coverage of 80% as shown.

**Figure S7.11.**
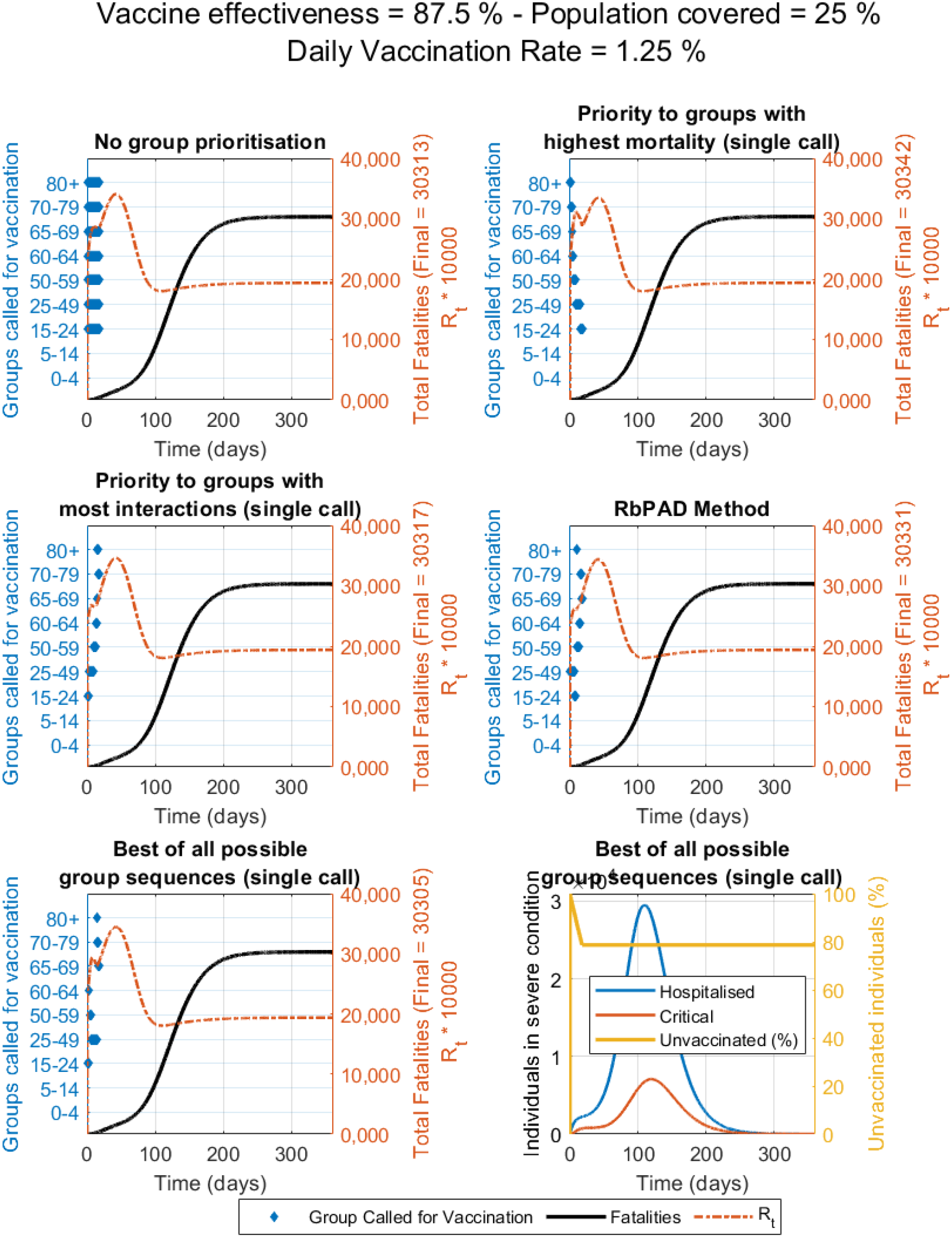
Compared vaccination strategies showing active groups called for vaccination (in blue bars), fatalities (thick black line) and R_t_ value over time (dotted orange line) for the case study with data and demographics from Spain. Constant vaccination rate is set at 1.25% of the total population per day, vaccine effectiveness of 87.5% and population coverage of 25% as shown.

**Figure S7.12.**
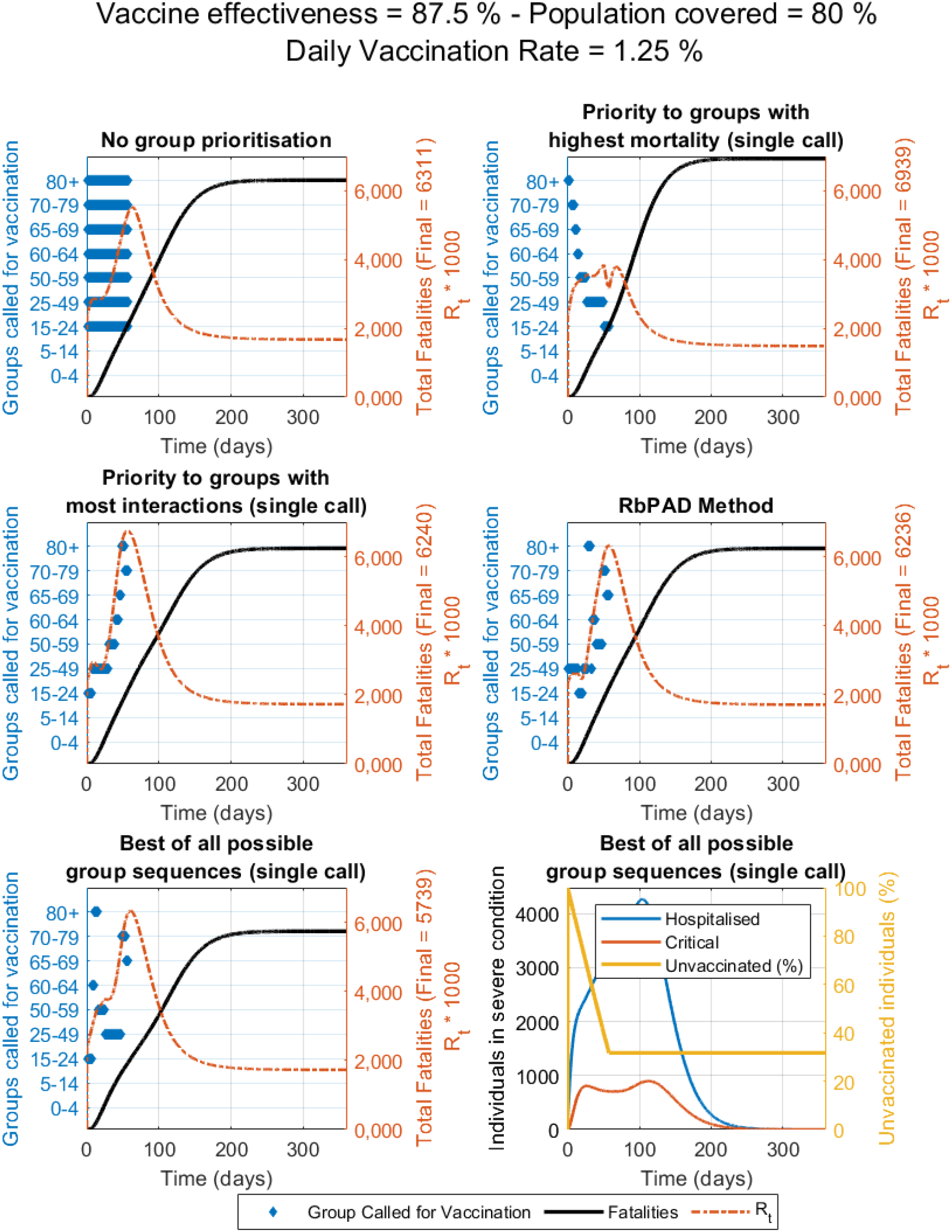
Compared vaccination strategies showing active groups called for vaccination (in blue bars), fatalities (thick black line) and R_t_ value over time (dotted orange line) for the case study with data and demographics from Spain. Constant vaccination rate is set at 1.25% of the total population per day, vaccine effectiveness of 87.5% and population coverage of 80% as shown.

**Figure S7.13.**
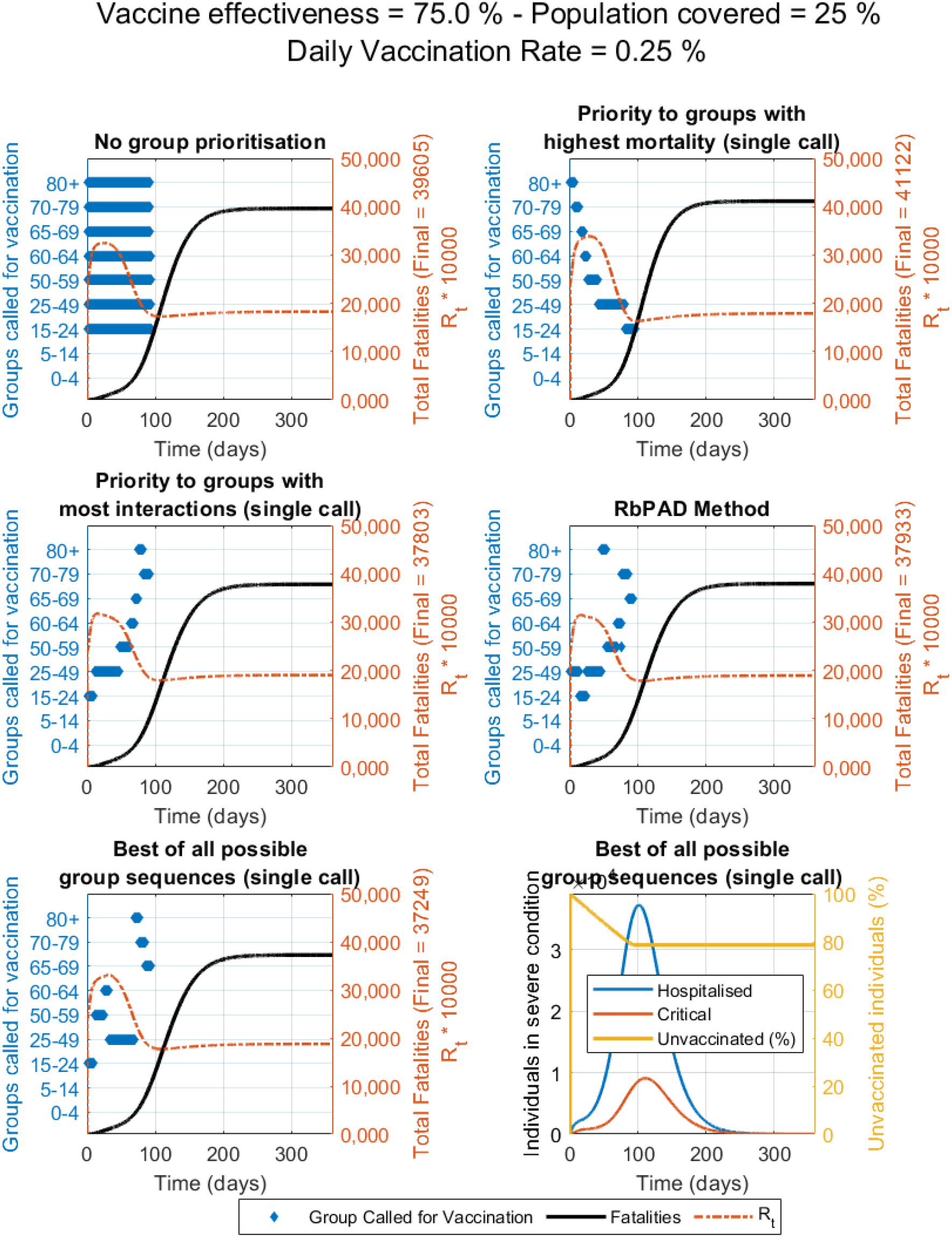
Compared vaccination strategies showing active groups called for vaccination (in blue bars), fatalities (thick black line) and R_t_ value over time (dotted orange line) for the case study with data and demographics from Spain. Constant vaccination rate is set at 0.25% of the total population per day, vaccine effectiveness of 75% and population coverage of 25% as shown.

**Figure S7.14.**
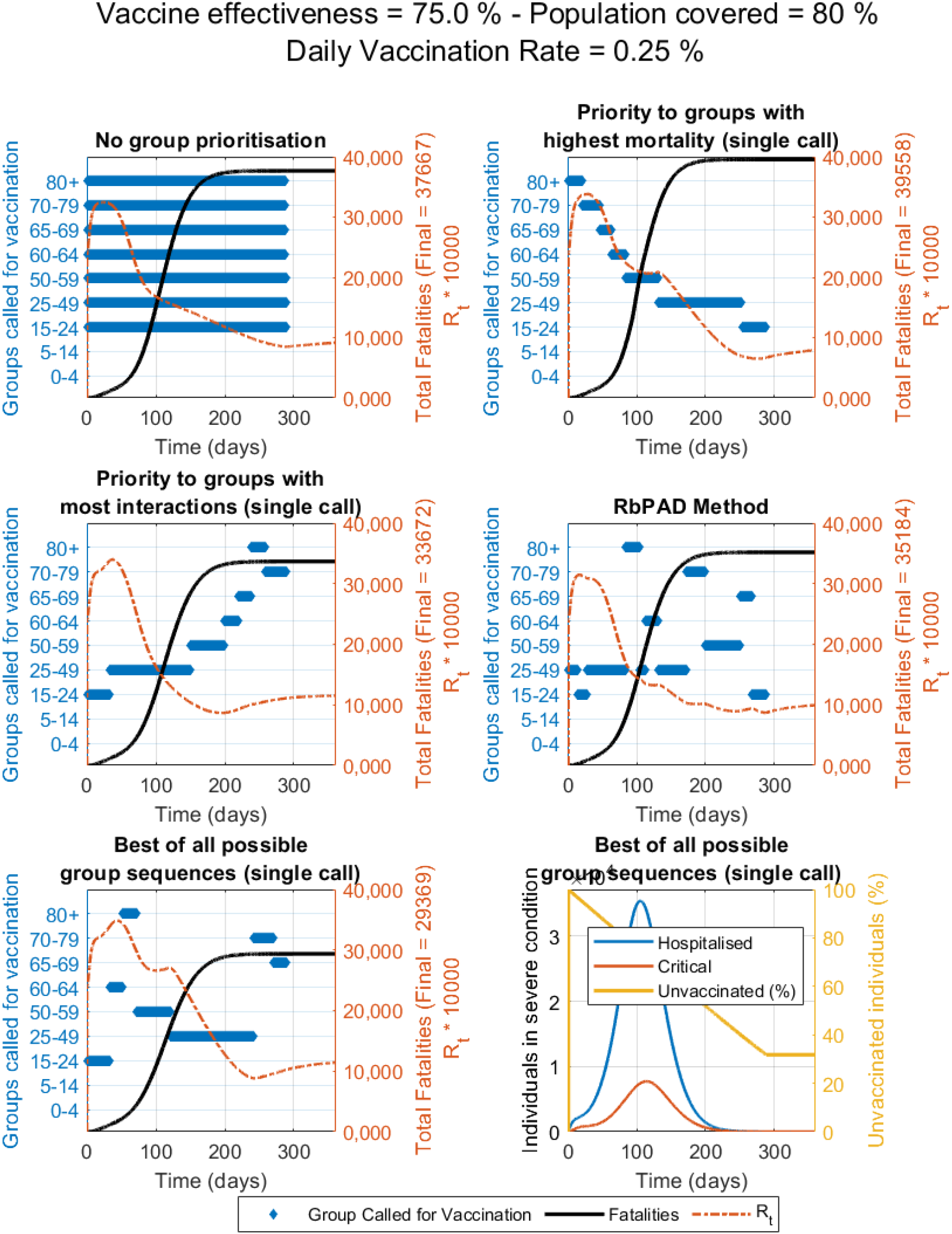
Compared vaccination strategies showing active groups called for vaccination (in blue bars), fatalities (thick black line) and R_t_ value over time (dotted orange line) for the case study with data and demographics from Spain. Constant vaccination rate is set at 0.25% of the total population per day, vaccine effectiveness of 75% and population coverage of 80% as shown.

**Figure S7.15.**
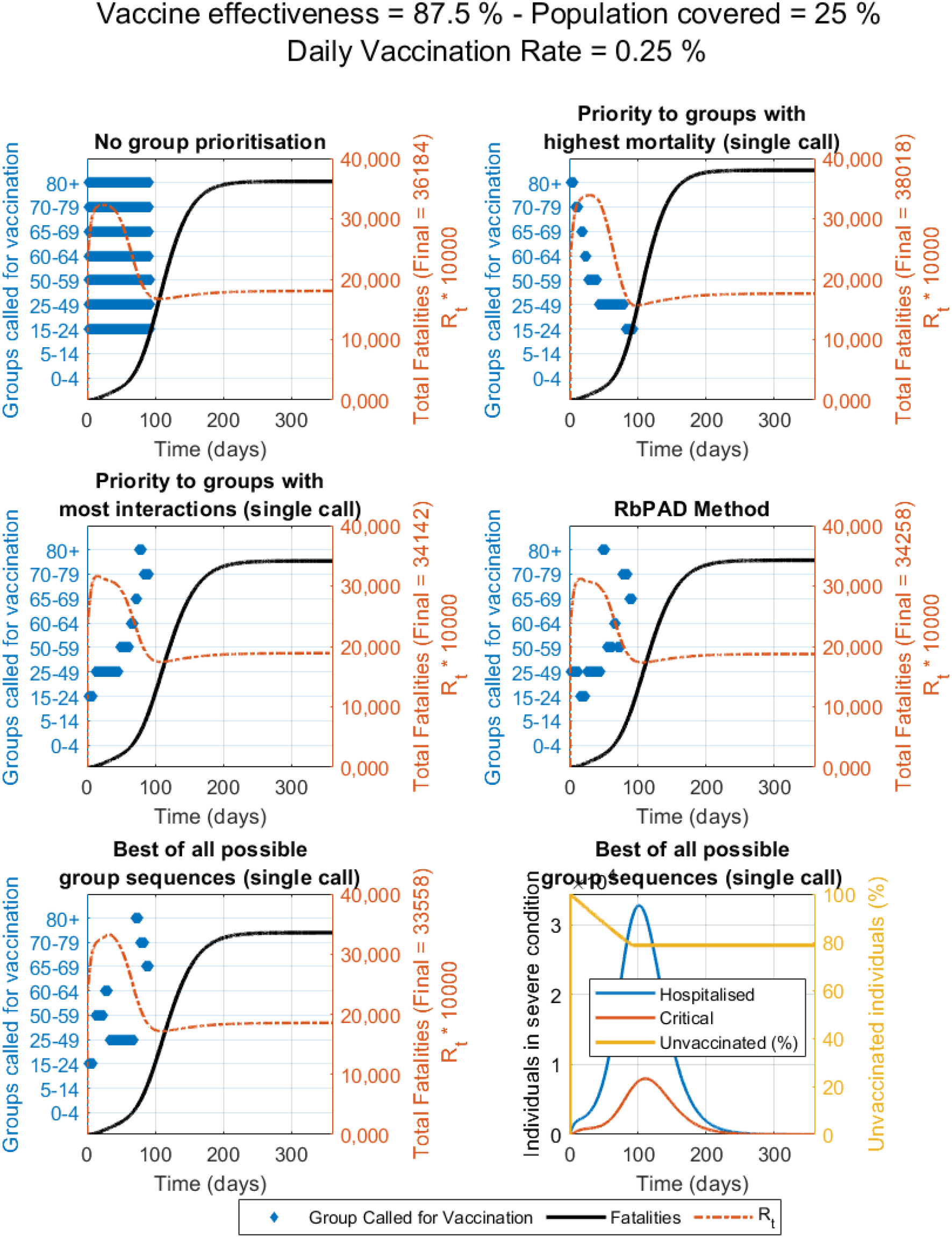
Compared vaccination strategies showing active groups called for vaccination (in blue bars), fatalities (thick black line) and R_t_ value over time (dotted orange line) for the case study with data and demographics from Spain. Constant vaccination rate is set at 0.25% of the total population per day, vaccine effectiveness of 87.5% and population coverage of 25% as shown.

**Figure S7.16.**
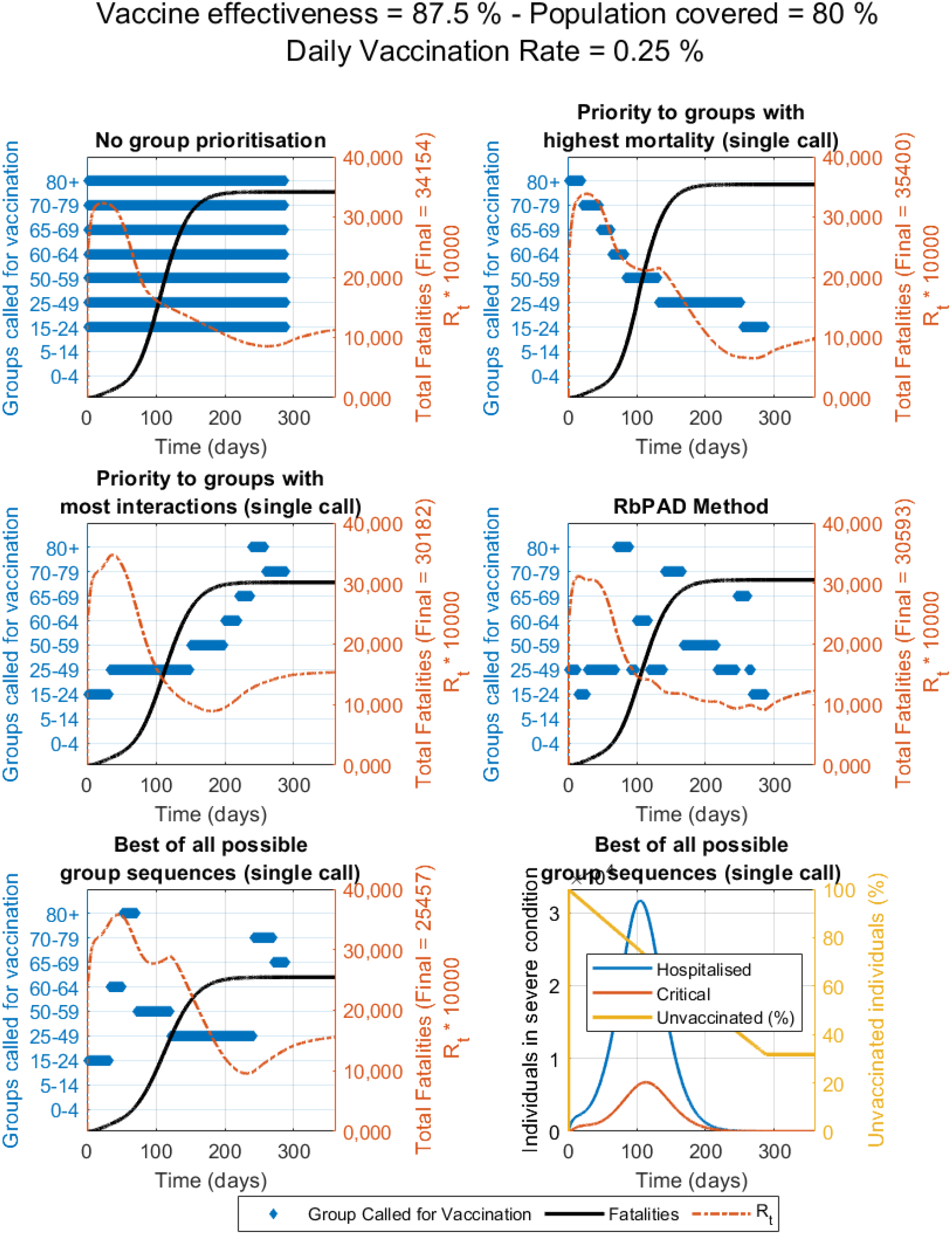
Compared vaccination strategies showing active groups called for vaccination (in blue bars), fatalities (thick black line) and R_t_ value over time (dotted orange line) for the case study with data and demographics from Spain. Constant vaccination rate is set at 0.25% of the total population per day, vaccine effectiveness of 87.5% and population coverage of 80% as shown.

## Supplementary section VIII. Impact of level of self-protection and awareness

As discussed in the manuscript, the level of protection and awareness (lpa) is a single-number description that incorporates habits such as the use of mask, and the general attitude and awareness to protect oneself and others. Since they are parameters with the largest uncertainty, a sensitivity analysis that evaluates the impact of the values selected is required. Ranges of values for younger than 65 years old and for older than 65 years old were evaluated.

The impact of the lpa values for the different age groups at the vaccine effectiveness and population coverage in the manuscript. Different daily vaccination rates ranging from 0.25 to 1 were also evaluated. Section VIII-A shows the results for the scenario in which children are also vaccinated while section VIII-B shows the results for the scenario in which children are not vaccinated.

The results in Section VIII-A show that, at daily vaccination rates higher than 0.50%, the strategy of prioritising the vaccination of people based on the number of interactions appears to be substantially better than a vaccination that prioritises by mortality. Only at lower daily vaccination rates (0.25%) it appears that the criteria of prioritising by mortality could be better in the case that people under 65 and over 65 do not use protection.

On the other hand, the results in Section VIII-B (when children are not vaccinated) show that the criteria of vaccinating by interactions would only obtain gains if the population coverage is 50%. The number of people that can be saved however is much lower than the number of fatalities avoided if all people was available for vaccination.

### A. Impact of the behavioural parameters of level of self-protection and awareness when children are also vaccinated

**Figure S8.1.**
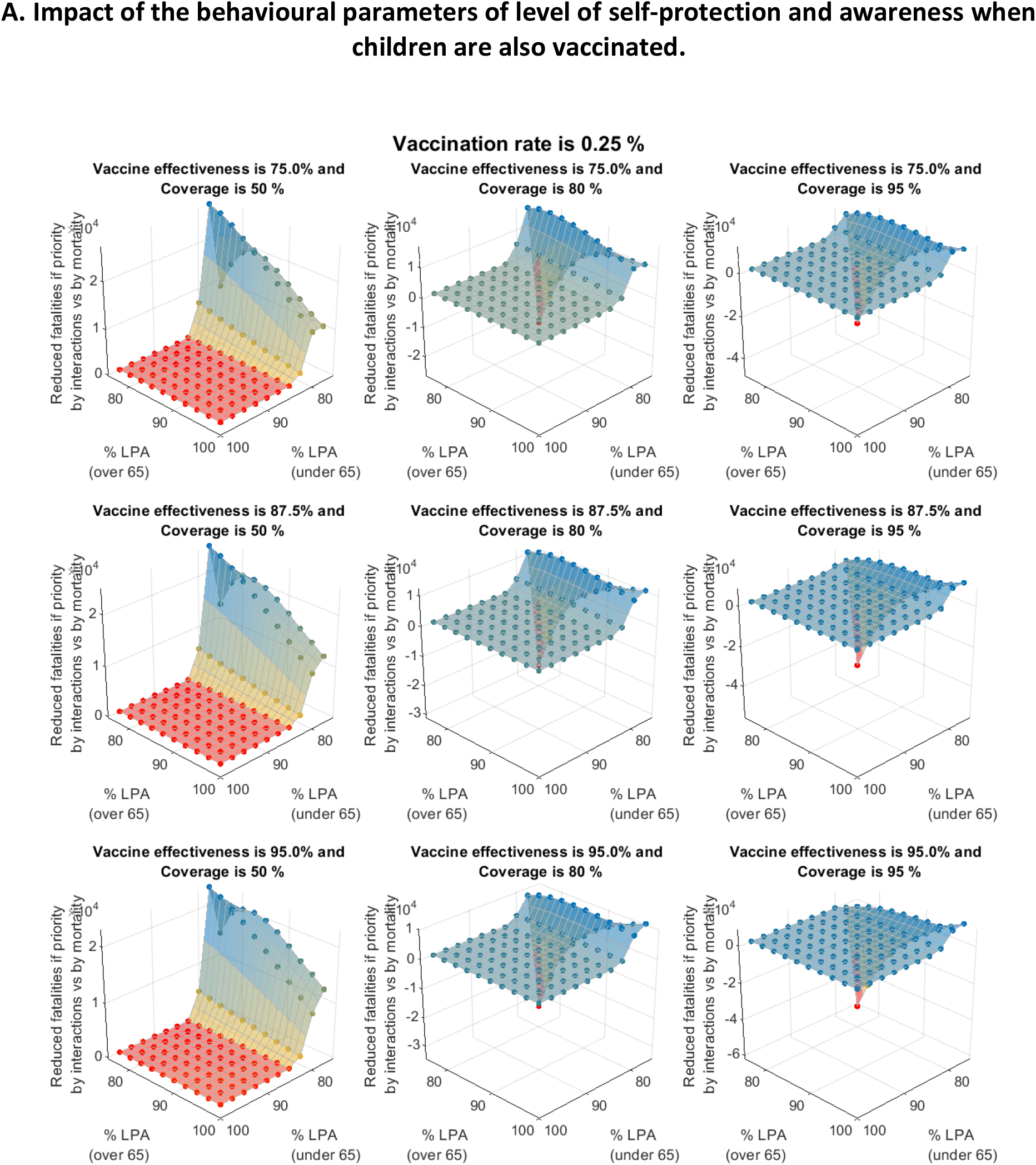
Impact of the different levels of self-protection and awareness (for those under 65 and for those over 65 years of age) on the number of fatalities that can be avoided if the vaccination is distributed with priority to groups from the highest to the lowest daily interactions vs. from the highest to the lowest mortality. The daily vaccination rate considered is 0.25% per day.

**Figure S8.2.**
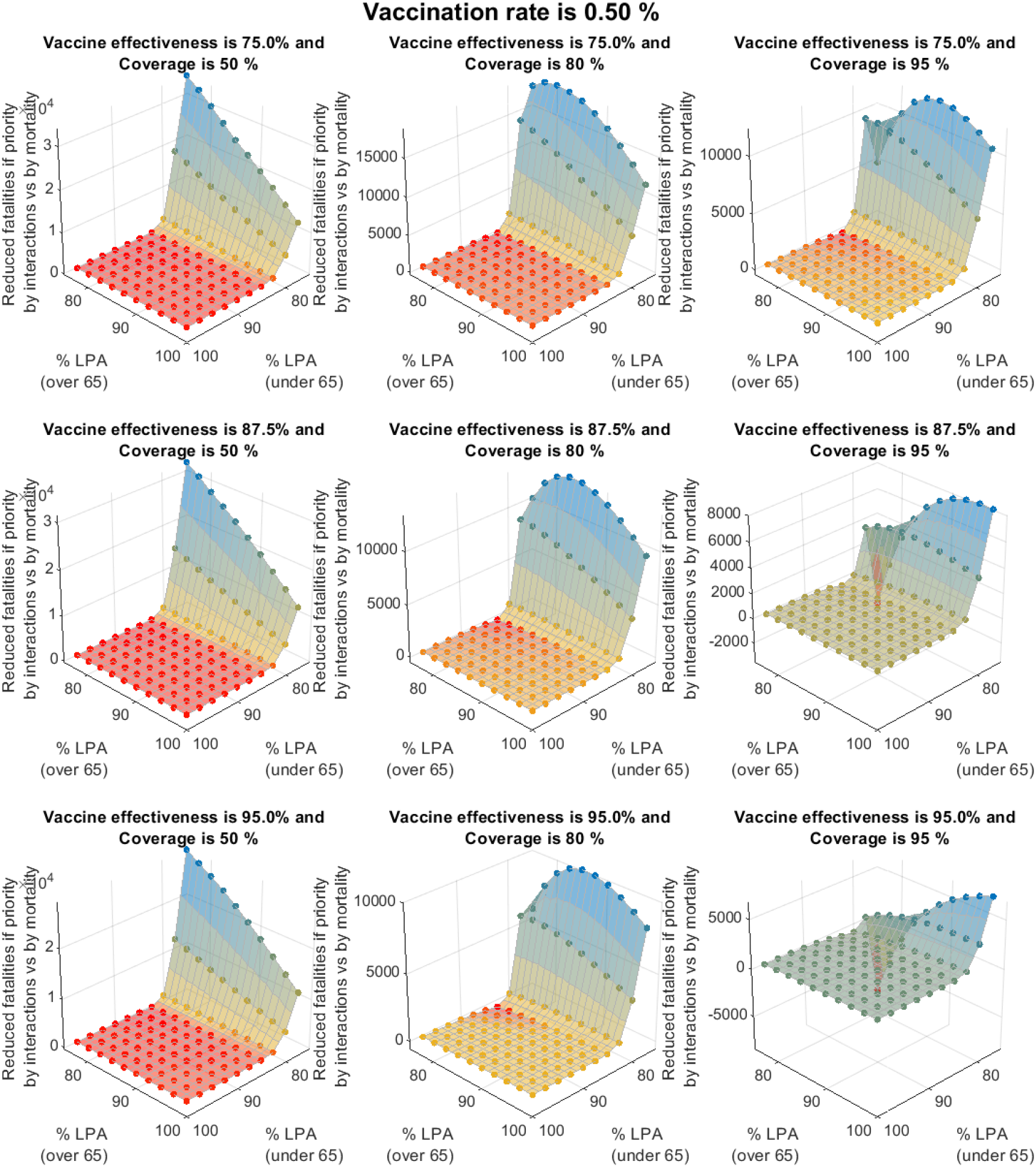
Impact of the different levels of self-protection and awareness (for those under 65 and for those over 65 years of age) on the number of fatalities that can be avoided if the vaccination is distributed with priority to groups from the highest to the lowest daily interactions vs. from the highest to the lowest mortality. The vaccination rate considered is 0.5% per day.

**Figure S8.3.**
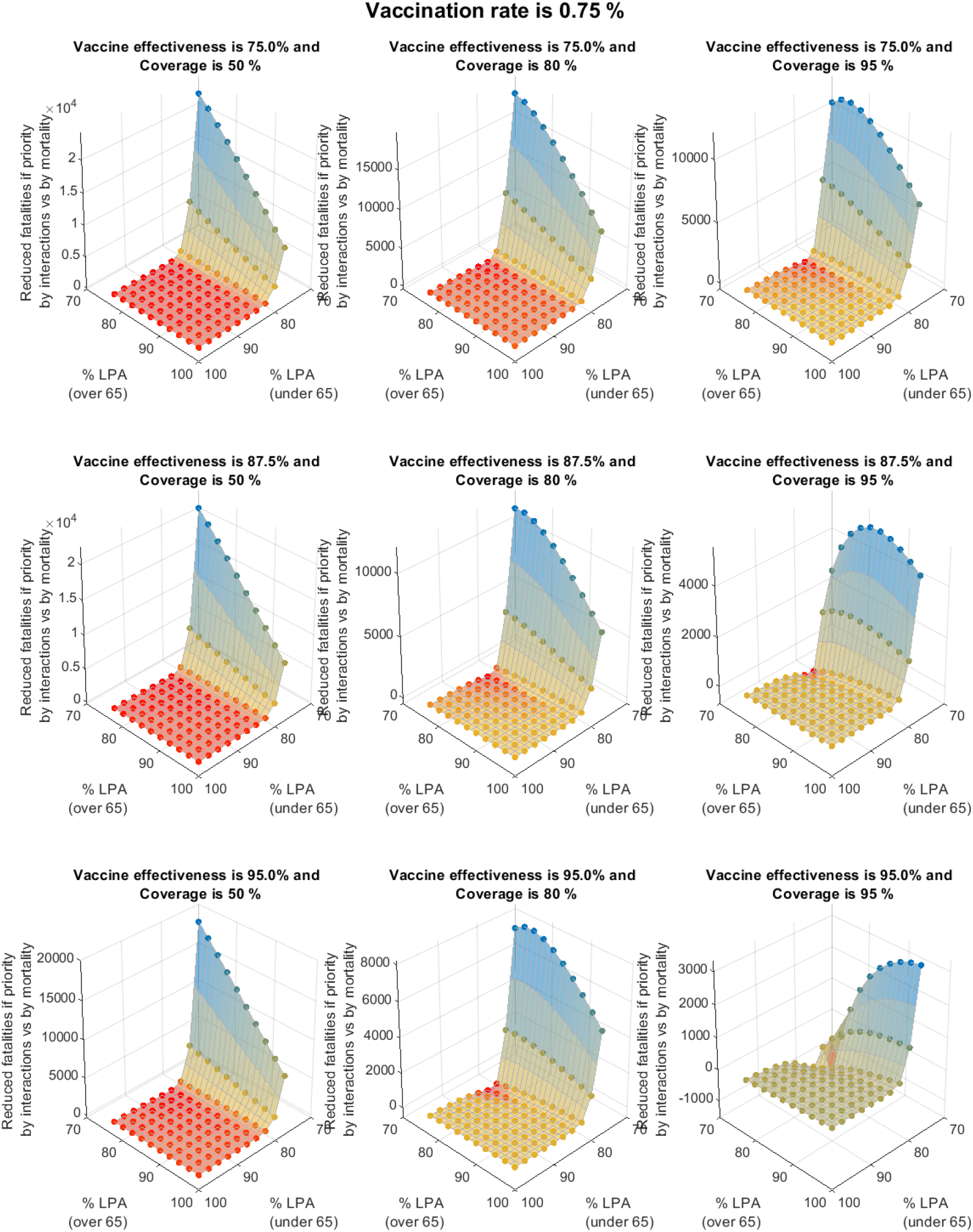
Impact of the different levels of self-protection and awareness (for those under 65 and for those over 65 years of age) on the number of fatalities that can be avoided if the vaccination is distributed with priority to groups from the highest to the lowest daily interactions vs. from the highest to the lowest mortality. The vaccination rate considered is 0.75% per day.

**Figure S8.4.**
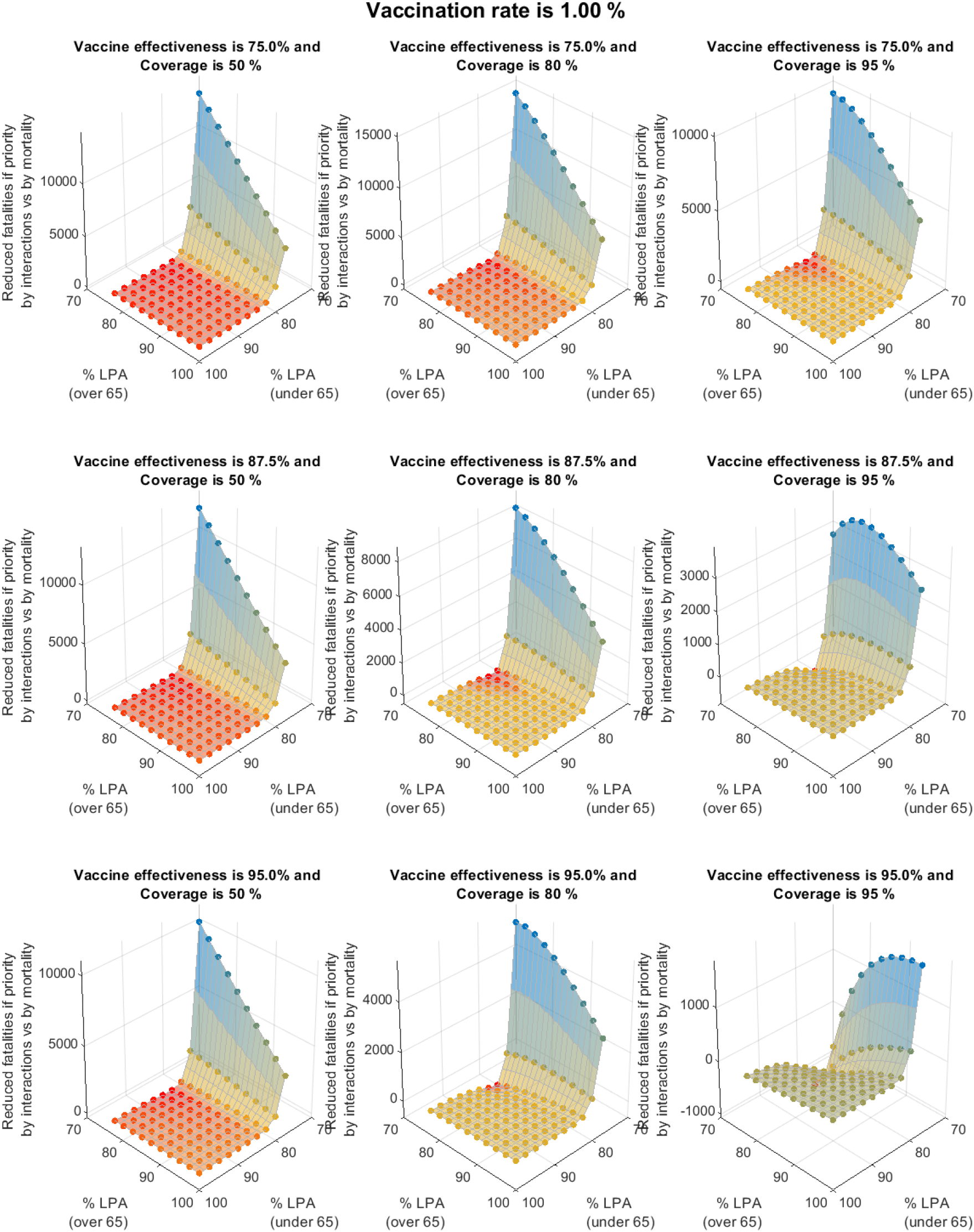
Impact of the different levels of self-protection and awareness (for those under 65 and for those over 65 years of age) on the number of fatalities that can be avoided if the vaccination is distributed with priority to groups from the highest to the lowest daily interactions vs. from the highest to the lowest mortality. The vaccination rate considered is 1% per day.

### B. Impact of the behavioural parameters of level of self-protection and awareness when children are not vaccinated

**Figure S8.5.**
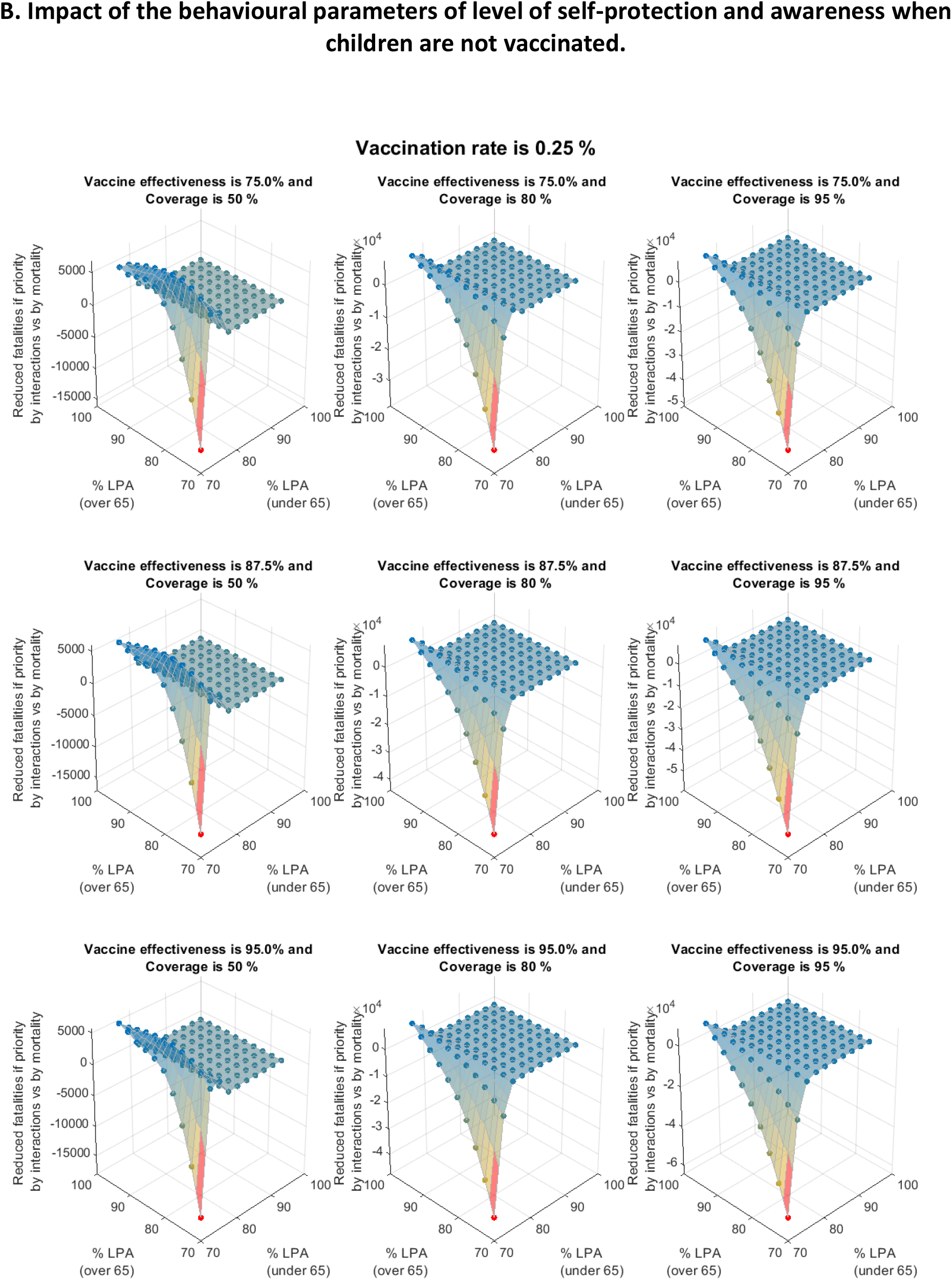
Impact of the different levels of self-protection and awareness (for those under 65 and for those over 65 years of age) on the number of fatalities that can be avoided if the vaccination is distributed with priority to groups from the highest to the lowest daily interactions vs. from the highest to the lowest mortality. The vaccination rate considered is 0.25% per day.

**Figure S8.6.**
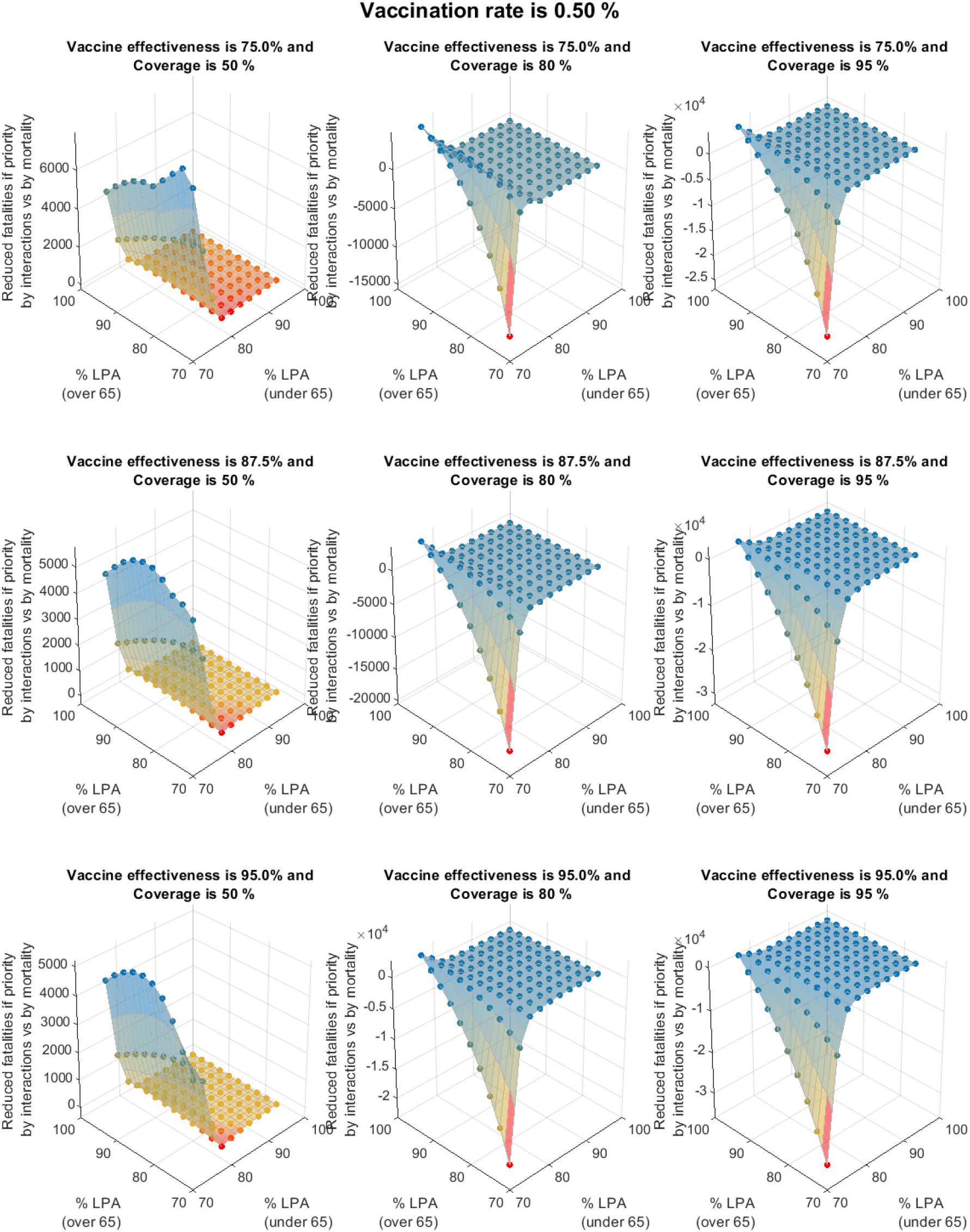
Impact of the different levels of self-protection and awareness (for those under 65 and for those over 65 years of age) on the number of fatalities that can be avoided if the vaccination is distributed with priority to groups from the highest to the lowest daily interactions vs. from the highest to the lowest mortality. The vaccination rate considered is 0.5% per day.

**Figure S8.7.**
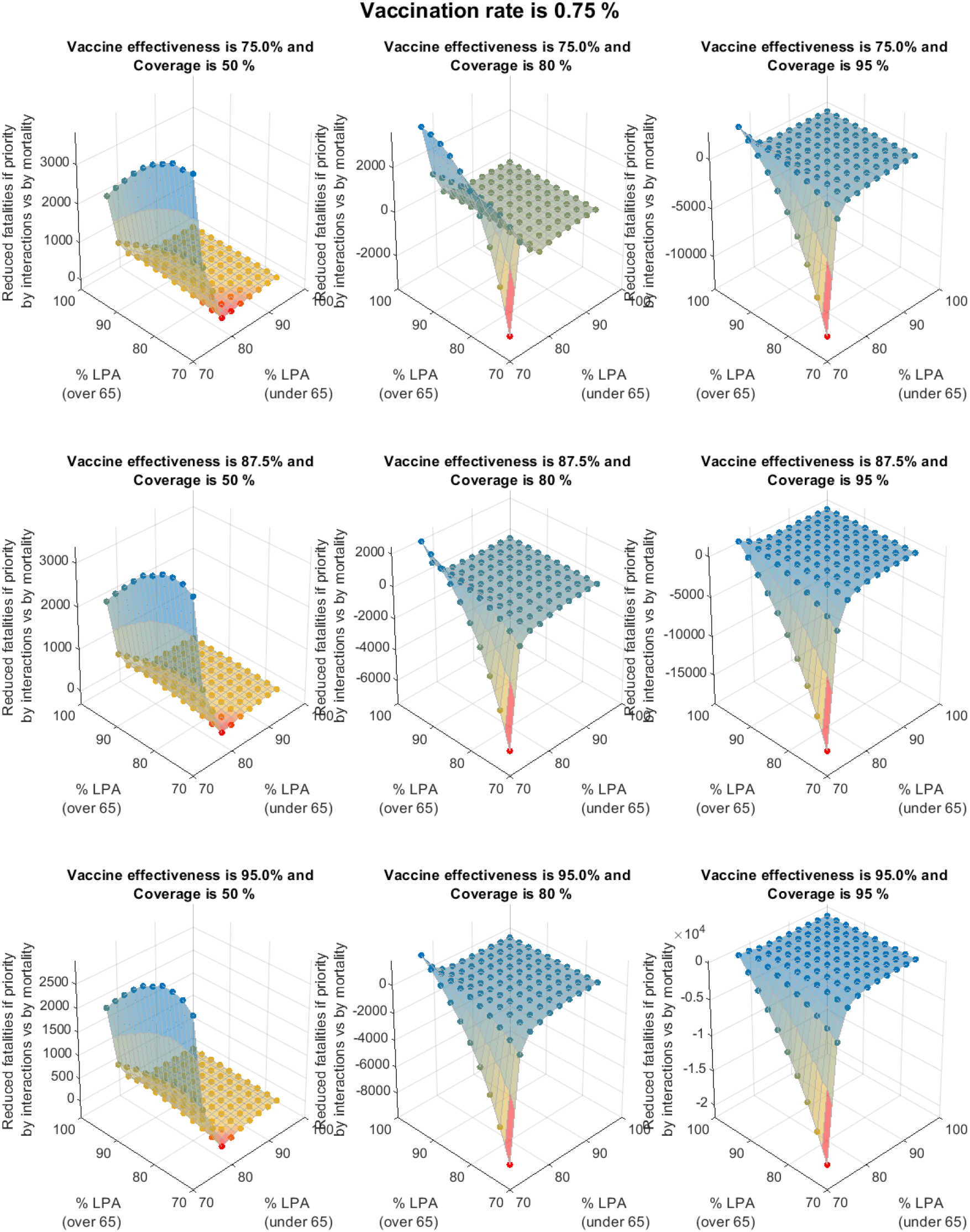
Impact of the different levels of self-protection and awareness (for those under 65 and for those over 65 years of age) on the number of fatalities that can be avoided if the vaccination is distributed with priority to groups from the highest to the lowest daily interactions vs. from the highest to the lowest mortality. The vaccination rate considered is 0.75% per day.

**Figure S8.8.**
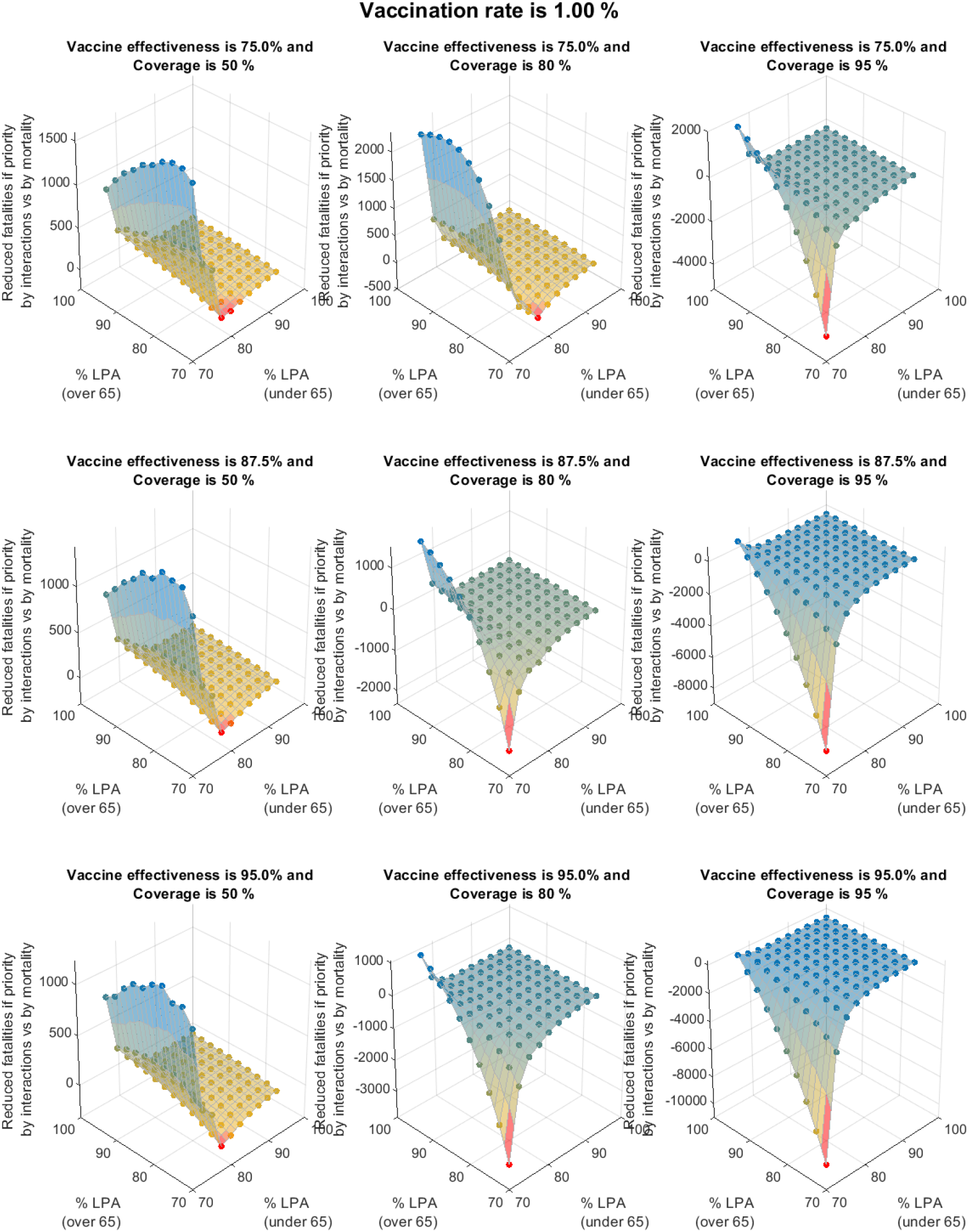
Impact of the different levels of self-protection and awareness (for those under 65 and for those over 65 years of age) on the number of fatalities that can be avoided if the vaccination is distributed with priority to groups from the highest to the lowest daily interactions vs. from the highest to the lowest mortality. The vaccination rate considered is 1% per day.

## Supplementary section IX. Impact of lpa parameters of R_t_ values

For the example case presented in the manuscript, the estimated behavioural parameters regarding levels of self-protection and awareness as shown in Table S5.2 were used. These values were arbitrarily selected and therefore remain of low confidence. An analysis was conducted of their magnitude such that they lead to typical R_t_ values observed. Figure S9.1 shows the results for the R_t_ are shown for different values of lpa for all age groups. Only values of lpa of 0.9-0.95 appear to yield expected R_t_ values throughout the simulation.

**Figure S9.1.**
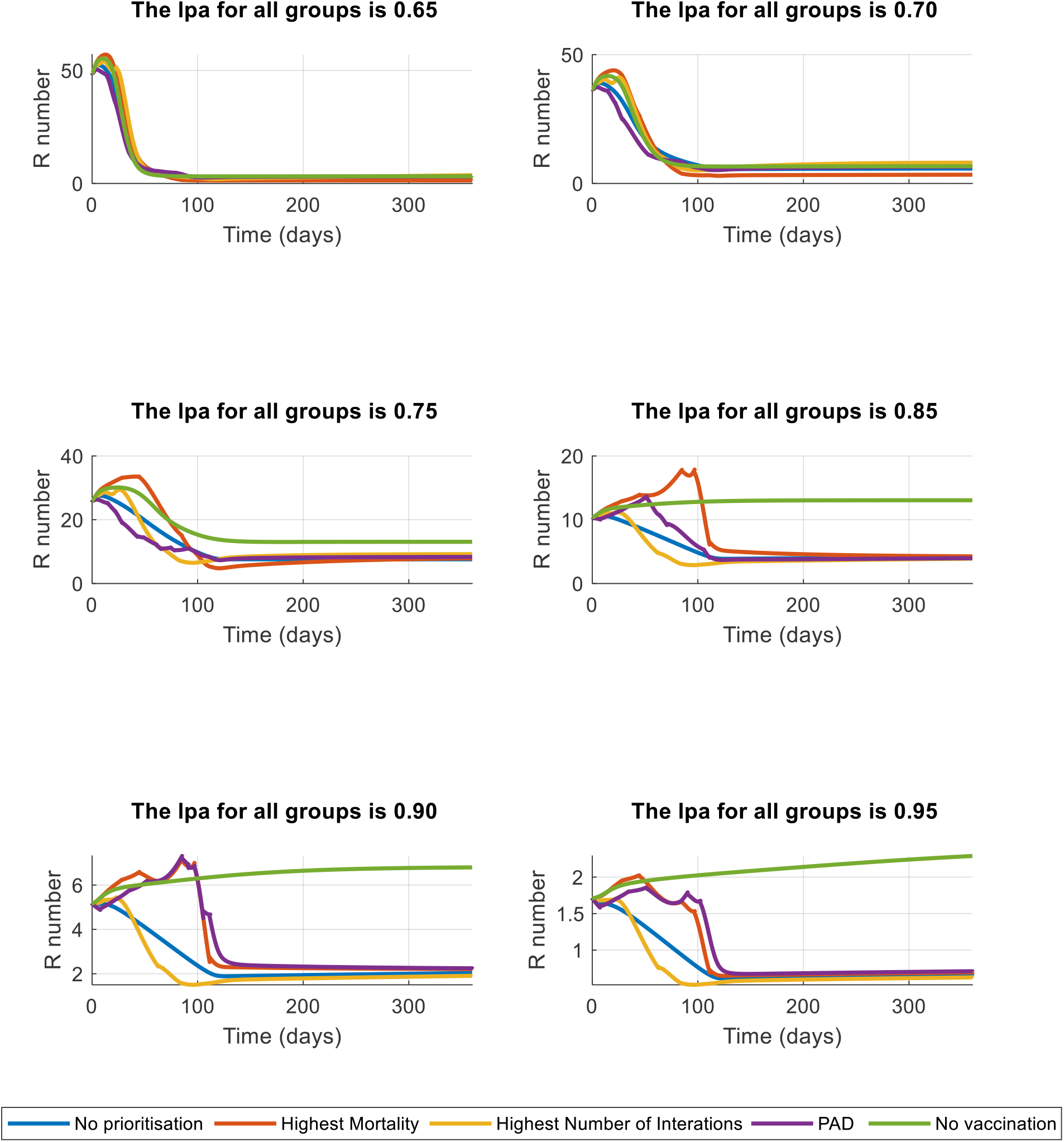
Magnitude estimation of behavioural level of protection and awareness parameters based on typically observed R_t_ values.

## Supplementary section X. Sensitivity of the daily number of interactions and the level of protection awareness

A sensitivity analysis on the number of interactions and the level of protection and awareness is shown in Figures S10.1-2. Different values of vaccine effectiveness for population coverage of 80% and a daily vaccination rate of 1% were evaluated. The number of interactions were multiplied then by a factor of 0.75, 1 and 1.25 to evaluate the difference between the strategy of prioritising by interaction vs. prioritising by mortality. As in previous sections, this was evaluated for an scenario in which children are vaccinated (Section X-A) and for an scenario in which children are not vaccinated (Section X-B).

Under a scenario in which all population is vaccinated, Figure S10.1 shows that the differences between strategies are fairly similar when interactions decrease 25%. Under this scenario however, it appears that a vaccination strategy that prioritises by interactions criteria rather than by mortality can decrease substantially the number of fatalities, as described in the main manuscript. The higher the number of interactions, the higher is the number of fatalities avoided by following this strategy. Under a scenario in which children are not vaccinated, Figure S10.2 shows that the differences between strategies are fairly similar when interactions decrease 25%. On the other hand, an increase of 25% in interactions increases substantially the fatalities, particularly if elders and youngers are not protected. Higher efficacy of the vaccine also appears to favour vaccination following a mortality criteria rather than an interactions-based one.

**Figure S10.1.**
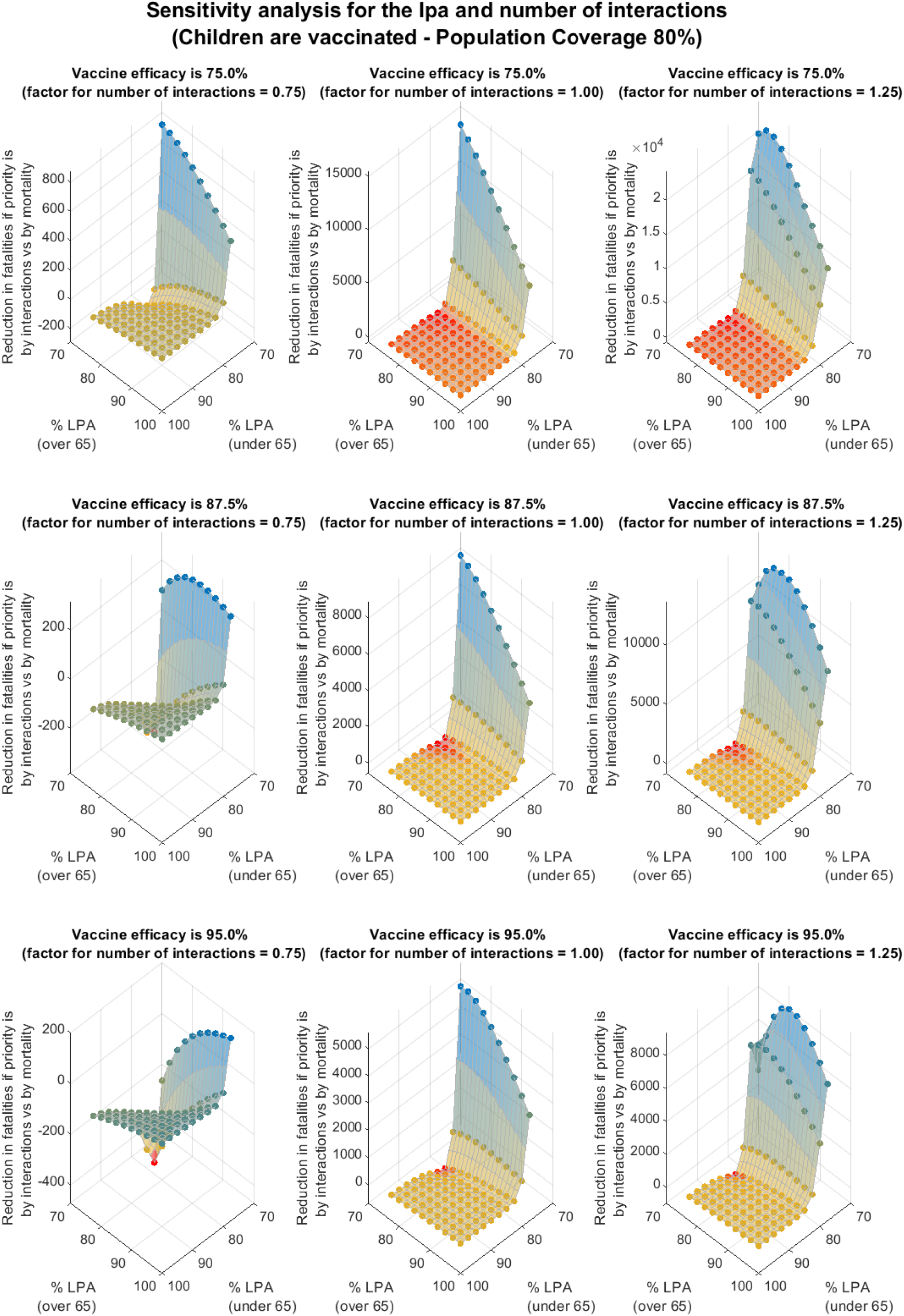
Sensitivity analysis for the number of interactions defined for the group age of 0-4 years old. Dark red bars indicate the group age in which their number of interactions is modified from the reference state.

**Figure S10.2.**
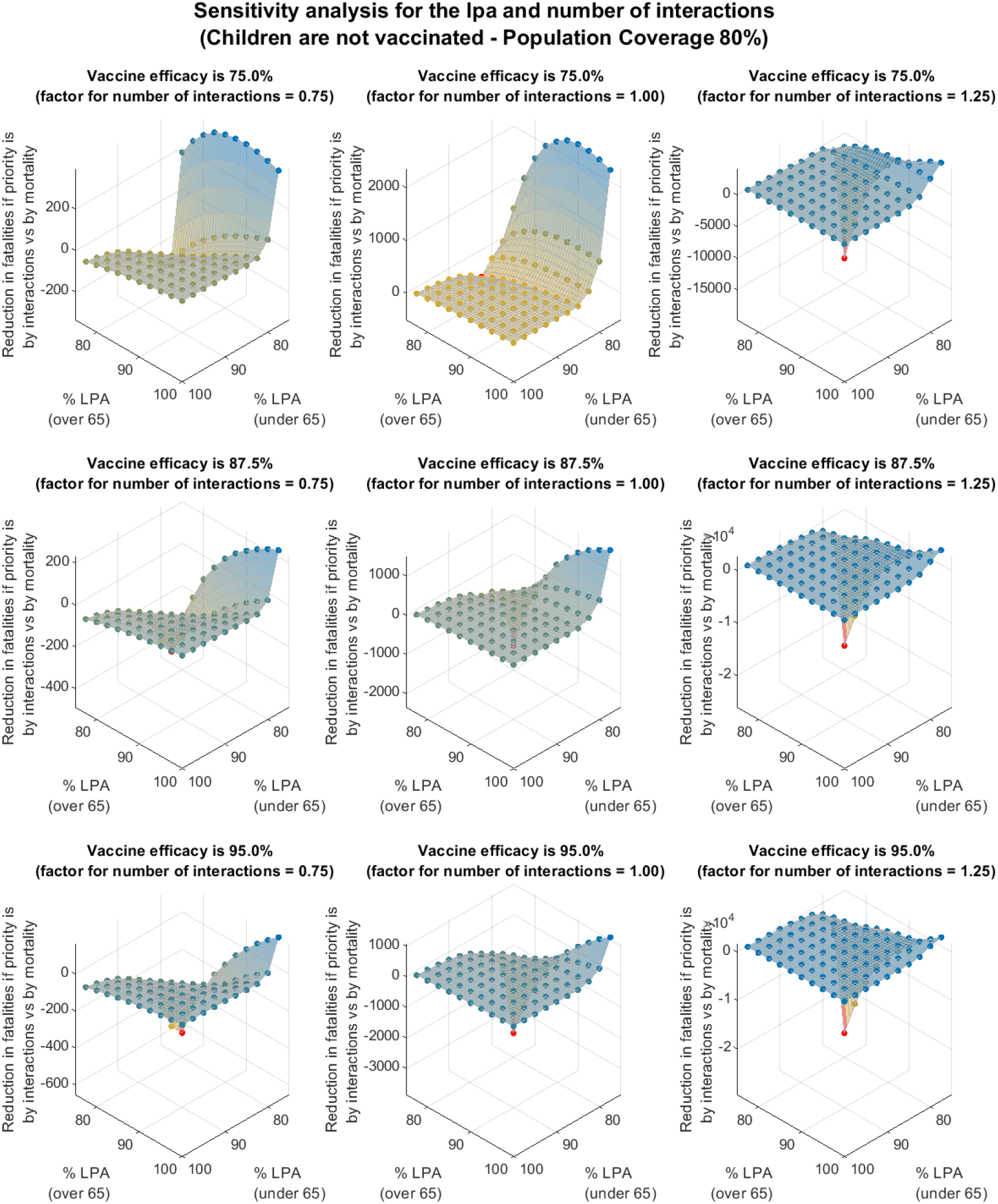
Sensitivity analysis for the number of interactions defined for the group age of 0-4 years old. Dark red bars indicate the group age in which their number of interactions is modified from the reference state.

